# Mechanistic Insights into Cancer Risk from the Circulating Proteome

**DOI:** 10.1101/2025.06.04.25328977

**Authors:** Valur Emilsson, Valborg Gudmundsdottir, Sean Bankier, Elisabet A. Frick, Thorarinn Jonmundsson, Kari Arnarson, Hulda K. Ingvarsdottir, Heida Bjarnadottir, Joseph Loureiro, Eva Jacobsen, Thor Aspelund, Eirikur Briem, Lenore J. Launer, Esther Bastiaannet, Tom Michoel, Sigurdis Haraldsdottir, Sigridur K. Bodvarsdottir, Thorarinn Gudjonsson, Nancy Finkel, Anthony P. Orth, Lori L. Jennings, John R. Lamb, Vilmundur Gudnason

## Abstract

Early, minimally invasive detection of cancer, ideally through biomarkers that also provide mechanistic insight, has the potential to substantially improve patient outcomes and reduce societal burden. To advance this goal, we examined associations between 7,523 circulating serum proteins and 13 cancer types in 5,376 older adults from the AGES cohort, including 1,235 individuals diagnosed with incident or prevalent cancers. The cancers analyzed spanned the digestive, genitourinary, respiratory, female reproductive systems, and skin. After adjusting for confounders and conducting sex-specific analyses, 526 proteins exhibited significant associations with cancer status. Additionally, 776 proteins were regulated by genetic cancer susceptibility loci, with 114 of these overlapping with cancer associations. Both sets were highly enriched for known cancer genes. Proteome-wide forward two-sample Mendelian randomization, leveraging 2,062 *cis*-acting pQTL instruments, identified 112 proteins with evidence of a causal influence on cancer risk. To place these findings in a broader biological context, we integrated them with genetic, tissue-specific expression, and tumor-specific datasets, thereby delineating the complex molecular landscape underlying cancer risk. This work illustrates the promise of serum proteomics for large-scale surveillance, early detection, and mechanistic insights into tumorigenesis.

## Introduction

Tumorigenesis is an evolutionary process, where cell populations gradually acquire autonomy from tissue control through cumulative genetic and epigenetic perturbations, fostering genomic instability and clonal expansion^1,2^. While environmental exposures and inherited susceptibility contribute^2–5^, the mechanisms underlying these transitions remain incompletely characterized, complicating efforts at early diagnosis and prevention. Among females, breast, lung, and colorectal cancers account for half of new diagnoses^6^, whereas prostate, lung, and colorectal cancers are predominant in men^6^. Despite declining mortality, attributable to improved detection, therapies, and lifestyle changes, cancer remains the second leading global cause of death^7^, fueled by aging demographics and increasing incidence among younger groups^6^.

Cancer risk reflects multifactorial influences, combining rare high-penetrance germline variants (such as BRCA1/2), common polymorphisms, somatic driver mutations, and environmental factors^8–14^. Genomic instability, chromosomal deletions or rearrangements, and epigenetic dysregulation accumulate, especially in advanced disease^14,15^. Evidence increasingly frames cancer as a systemic disorder, shaped by dynamic interactions between tumor, stroma, immune, and distal host tissues^8^. Current screening tools (e.g., mammography, Pap smear, chest imaging) for early detection encounter challenges of high false positive rates and low adherence^16^. Classic blood biomarkers such as PSA (KLK3), AFP, CEA and CA-125 (MUC16) remain limited in scope and specificity^17^. Emerging plasma-based assays targeting circulating cell-free DNA mutations and methylation offer hope for earlier detection^18,19^. However, large-scale, unbiased serum proteome profiling remains underexplored. Notably, recent proteomic analyses from the UK Biobank have revealed associations between plasma protein levels and cancer risk^17^, underscoring the need for further systematic investigation.

Comprehensive surveys of the circulating proteome in relation to cancer status, both before and after diagnosis, are warranted, given that protein variation reflects coordinated systemic regulation and that genetic factors broadly influence circulating protein levels, many of which are implicated in diverse complex diseases^20–31^. While many serum and plasma proteins are tightly regulated and linked to disease, their roles in cancer susceptibility, diagnosis, and progression are poorly mapped. Here, we leveraged highly sensitive aptamer-based profiling of 7,523 serum proteins in the prospective, population-based Age, Gene/Environment Susceptibility Reykjavik (AGES) cohort^32^, examining their associations with both prevalent cancer and incident cancer diagnoses during follow-up, with rigorous adjustment for risk factors and sex-specific stratification. The study additionally investigated the relationship between serum proteins and genetic cancer risk factors, applied forward Mendelian randomization (MR) analysis for causal inference, and contextualized cancer-associated proteins within a functional framework designed to elucidate the roles of distinct serum proteins in cancer risk.

## Results

### Study population and overview

This study leverages the population-based, prospective AGES cohort^32^ of older adults (N = 5,764; mean age 76.6 ± 5.6 years, range 66–98; 57% female), which is extensively annotated for disease risk factors, comorbidities, genotypes, and serum proteomics, with real-time follow-up. In total, 7,523 aptamers, corresponding to 6,586 unique proteins, were measured in 5,376 participants, who form the focus of the present analysis. Table 1 summarizes selected baseline characteristics, and Fig. 1 provides an overview of cancer patient counts and the 13 cancer types analyzed across multiple organ systems: digestive (esophagus, stomach, colon, rectum, pancreas), genitourinary (kidney, prostate, bladder), female reproductive (breast, ovary, corpus uteri), respiratory (lung, bronchus), and skin (melanoma).

**Fig. 1.**
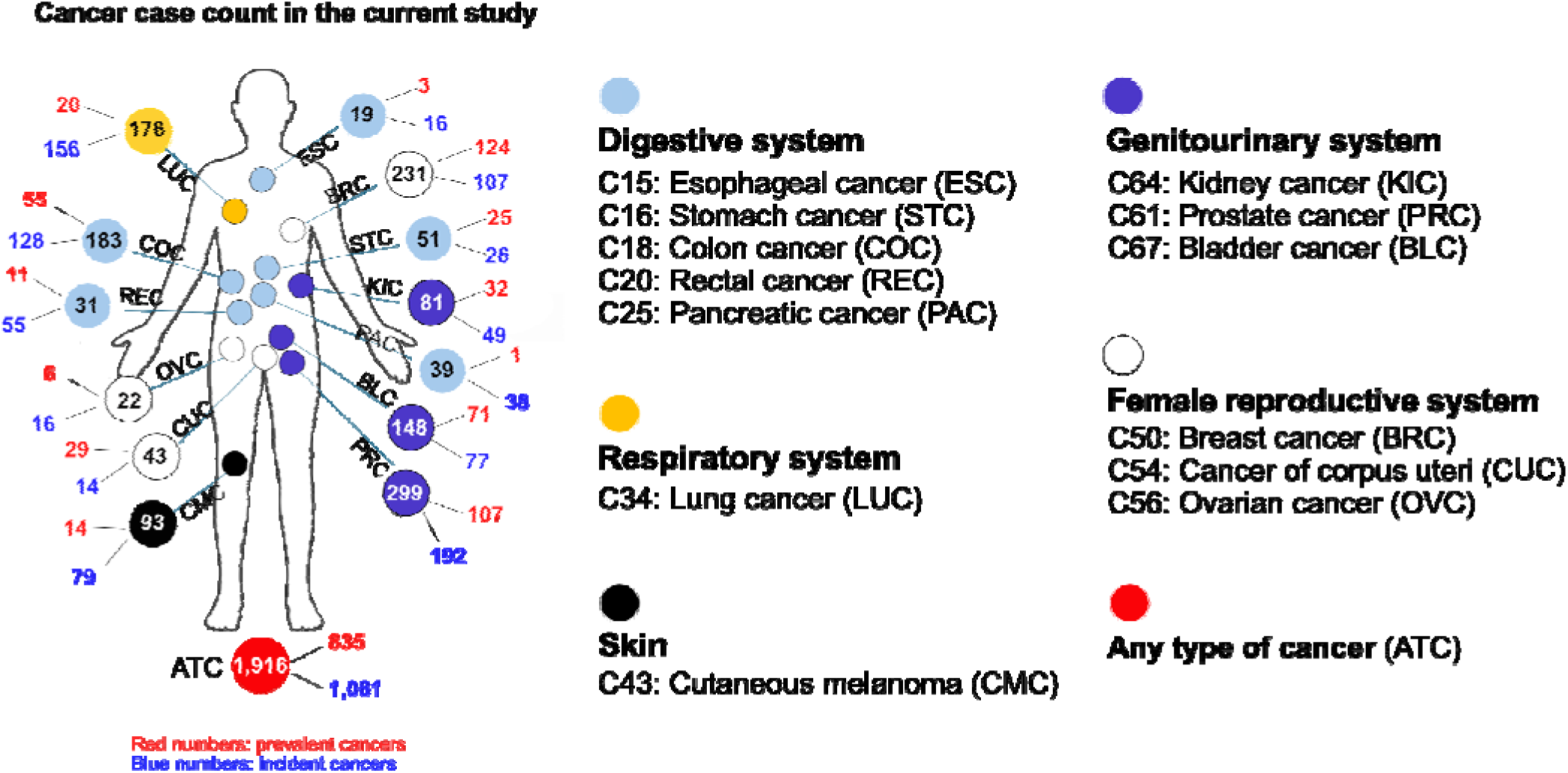
**Overview of cancer patient numbers and the types of cancer analyzed in this study**. The total number of individuals diagnosed with each of the 13 distinct cancer types, or with a history of any cancer (ATC), is shown within the circles. Cancer types are grouped by the anatomical organ systems in which they are diagnosed. Red numbers indicate prevalent cases; blue numbers indicate incident cases. Diagnoses were based on ICD-10 codes from the 10th revision of the WHO International Classification of Diseases, with specific codes listed for each cancer type. The cancer type abbreviations shown in this figure are used throughout the main text to refer to the respective cancer types.

**Table 1.**
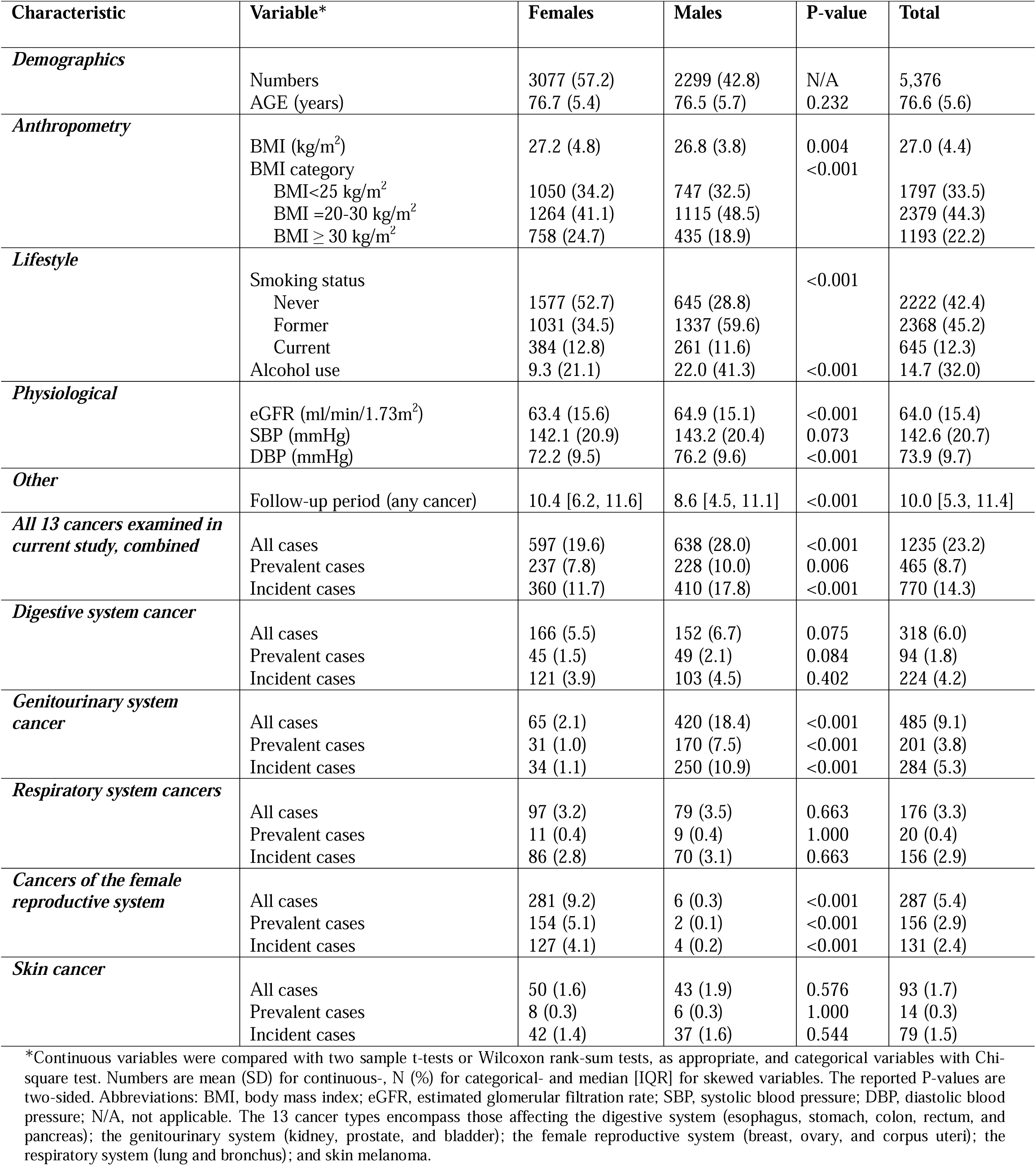
Baseline characteristics of AGES participants by sex, cancer group (body system), and disease status, with 7,523 serum proteins measured.

Incident cancers refer to new diagnoses occurring during up to 13.6 years of follow-up from baseline, whereas prevalent cancers were diagnosed prior to baseline. Across these 13 cancer types, 1,235 individuals were identified as either prevalent (38%, n = 465) or incident cases (62%, n = 770) (Table 1, Tables S1-S2). In total, there were 1,414 cancer diagnoses, including 179 individuals with more than one type of cancer; these individuals were included in analyses of each respective cancer rather than being excluded. Breast cancer was the only type with a higher proportion of prevalent cases at baseline (54%, n = 124) (Figure 1, Table S1). Supplementary Tables S2-S7 provide descriptive statistics for all 13 cancer types combined or grouped by the shared organ system, with incident and prevalent cases considered together.

Among modifiable risk factors, tobacco smoking, alcohol consumption, and obesity were strongly associated with cancer risk. Patients with digestive and female reproductive system cancers showed significantly different BMI distributions (Tables S3 and S6), and smoking status was associated with nearly all cancer types except skin cancer (Tables S2–S7). Individuals with genitourinary cancers reported higher alcohol consumption than those without these cancers (Table S4). Beyond the 13 primary cancer types, the AGES study included 684 additional cancer cases (Table S8), which showed no significant associations with common lifestyle risk factors (Table S9). Combining these with the 1,235 primary cases yielded a broad “any cancer” category (n = 1,916) for specific analyses. Given the study’s extensive scope, we highlight key findings in the main Results section, with Supplementary Note 1 detailing the study design and analytical framework, whereas Supplementary Notes 2-8 provide in-depth analyses by cancer type, grouped by share organ system for clarity.

### Observational study linking the serum proteome to incident cancers

To evaluate associations between serum proteins and incident cancers diagnosed after blood draw, a Cox proportional hazards model^33^ was applied to log2-transformed proteomic data. Unless stated otherwise, all models were adjusted for age, sex, and estimated glomerular filtration rate (eGFR) for kidney function (referred to as ’standard covariates’). Additional covariate adjustments, including BMI, alcohol consumption, smoking, height and blood pressure were used, along with sex-specific analyses for cancers not inherently sex-specific (Supplementary Note 1). Multiple testing was addressed using the Benjamini-Hochberg FDR method^34^. To enhance discovery and provide internal validation across cancer types within the AGES study, results are reported at FDR thresholds of <0.05 and <0.10. In the Cox regression analyses, 357 aptamers corresponding to 328 proteins were associated with at least one of 13 incident cancers at FDR < 0.05 (Table S10, Fig. 2A, Figs. S1A-S5A, Supplementary Notes 2-6), based on various covariate adjustments and sex-stratified analyses.

**Fig. 2.**
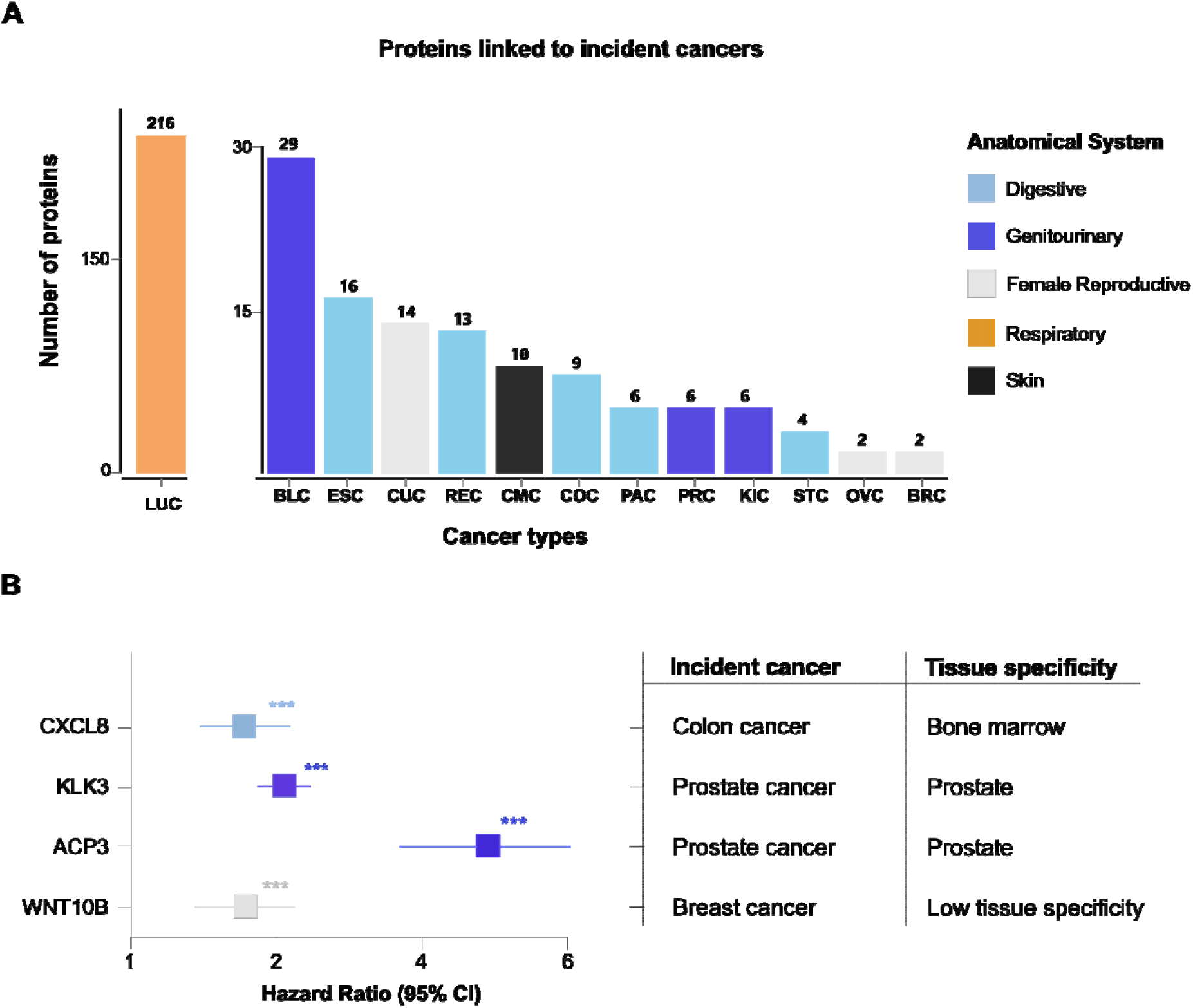
**The number of serum proteins associated with future risk of cancer**. (**A**) The number of serum proteins associated with each cancer type is shown using the abbreviations for different cancer types defined in Fig. 1. Cox regression analysis was performed with adjustment for age, sex, and eGFR, along with additional covariate and sex-specific adjustments tailored to each cancer type, as detailed in the Supplementary Note 1. Statistical significance was determined using Benjamini-Hochberg correction with a false discovery rate (FDR) threshold of < 0.05. Data columns are color-coded according to the anatomical system of tumor origin, with the number of significantly associated proteins indicated at the top of each column. Notably, a few proteins are linked to more than one cancer type. **(B**) Forest plot of log2-transformed protein values (hazard ratios) with 95% confidence intervals (CIs) for a selected subset of serum proteins associated with future cancer risk, stratified by tissue specificity. Data points are color-coded by the anatomical site of tumor origin and represent statistically significant findings (FDR < 0.05, ***). Horizontal lines denote 95% CIs.

Figure S6 shows the number of proteins identified using standard covariates compared with those identified using additional covariates across different incident cancer types based on epidemiological studies. Incident lung cancer (LUC) had the highest number of associated aptamers, or 216 (FDR < 0.05), while the remaining cancer types were associated with 2-29 aptamers each (Table S10, Fig. 2A). After adjustment for smoking status, 10 proteins remained significant for incident LUC (Fig. S6, Supplementary Note 7). More generally, while the number of proteins were reduced upon full adjustment for some incident cancer types the number increased for others (Fig. S6, Supplementary Note 7). This suggests that some associations observed with standard covariate adjustment may have been confounded or mediated by additional covariates, whereas others may have been masked by lifestyle-related factors or reflect biological pathways acting independently of them (Supplementary Note 7). Finally, the number of associated proteins appeared driven more by cancer type than by the number of incident cases; for example, bladder cancer (BLC), esophageal cancer (ESC), and rectal cancer (REC) had more associations despite fewer cases than prostate cancer PRC, BRC, and colon cancer (COC) (Figs. 1 and 2A, Fig. S6).

Only a few serum proteins were shared across cancer types, underscoring the origin-specific nature of these associations. For instance, at FDR < 0.05, SIGLEC6 was associated with incident BLC and PAC, while HAVCR1 was linked to incident KIC and LUC (Table S10). Additionally, 11 other proteins, including MPP2, TRAT1, TMEM106A, UBE2E3, TREM2, CREBBP, ITIH1, and RAB22A, were associated with multiple types of incident cancer at FDR < 0.05 or < 0.10 (Table S10). Interestingly, CREBBP is a tumor-specific protein encoded by well-established tumor suppressor gene^35,36^, with an enriched mutation frequency across several cancer types^37^. The other proteins may play diverse roles in tumor-host interactions, including immune modulation (SIGLEC6, MPP2, TRAT1)^38–40^, epithelial-mesenchymal transition (EMT) and/or metastasis (TMEM106A, RAB22A)^41,42^, tumor microenvironment (TME) remodeling (TREM2)^43^, and cellular senescence (UBE2E3)^44^. The kidney-specific expression of HAVCR1^45^, together with its association with incident KIC, suggests it may reflect tumor burden and could potentially contribute to organ-specific mechanisms of tumorigenesis.

While most circulating protein-cancer associations identified here are novel, several corresponding genes, expression traits, or proteins linked to future cancers in the AGES study have previously been implicated in their respective cancer types in other model systems. For instance, *CXCL8* (aka *IL8*), is a known pro-tumor effector gene in COC involved in resistance to immune checkpoint inhibitors and TME modulation^46,47^. Consistently, our study showed a positive association between CXCL8 and COC (Table S10, Fig. 2B). Additionally, KLK3 (aka PSA), a glycoprotein enzyme primarily produced by the prostate gland and one of the most well-known biomarkers for PRC, with well-established associations to multiple PRC-related clinical outcomes^48^, was positively associated with incident PRC (Table S10, Fig. 2B). Another example is acid phosphatase ACP3 (aka PAP), a protein produced exclusively by the prostate gland^45^, which was found to be strongly associated with incident PRC in the present study (Table S10, Fig. 2B). Elevated serum levels of ACP3 have been associated with accelerated progression of PRC^49^, and the protein is a key target for cellular immunotherapy, which has been shown to improve survival in men with metastatic PRC^50^. The observed associations between the prostate-specific proteins KLK3 and ACP3 and incident PRC align with their well-established roles as clinical biomarkers for prostate cancer^51^. WNT10B provides another example, showing an association with incident BRC in this study (Table S10, Fig. 2B) and having been previously linked to BRC^52,53^, particularly to aggressive subtypes such as triple-negative BRC. This association is thought to be driven by activation of the Wnt/β-catenin pathway, which is essential for development and tissue homeostasis but also contributes to poor prognosis and increased metastasis in BRC patients^52,53^. Additional examples include CTNNB1^54^, BCL2L14^55^, WNT7A^56–58^, PTPN6^59^, PRKCZ^60^, WFDC2 (aka HE4)^61–63^ and EPOR^64^, all of which were associated with one or more incident cancer type at an FDR < 0.05 in the current study (Table S10). While *CTNNB1* is a well-established canonical oncogene^35,36^, *EPOR* has more recently been implicated as an oncogene^35,36^, with truncated rearrangements identified in certain forms of leukemia^65,66^. Some proteins showed significant associations in one sex only (Table S10), including BCL2L14 with incident COC in males, CUL1 with incident STC in females, CSF1 with incident BLC in males, and ASMTL with incident CMC in females. Cancer-related roles of the remaining proteins are likely mediated through tumor-host interactions. These include inflammation and the mucin network (BCL2L14)^67^, the TME and metastasis (WNT7A)^68^, DNA damage response and apoptosis (CUL1)^69^, melatonin biosynthesis (ASMTL)^70^, as well as various immune-related functions (PTPN6, PRKCZ, CSF1 and WFDC2)^60,71–73^.

### Observational study linking the serum proteome to prevalent cancers

Associations between serum proteins and cancers diagnosed before blood draw were evaluated using logistic regression analysis, with various covariate adjustments and methods consistent with the incident cancer analysis (Methods, Supplementary Note 1). The analysis of prevalent cancers with ≥10 cases, identified 239 aptamers matching 223 proteins at FDR < 0.05 (Table S11, Fig. 3A, Figs. S1B-S5B, Supplementary Notes 2-6). Figure S7 compares the number of proteins identified using standard versus additional covariates across prevalent cancer types. Prevalent BRC had the highest number of associated proteins, or 144 (FDR < 0.05), while the remaining cancer types had 3-33 associated proteins each (Table S11, Fig. 3A). For example, in BRC, adjusting for additional risk factors (BMI, smoking, alcohol consumption) reduced the number to 100 proteins, while revealing 10 proteins not detected with standard covariate adjustment (Supplementary Note 5, Fig. S7). Like incident cancers, the number of associated proteins did not always correlate with the number of prevalent cases, but rather with the type of diagnosis. For example, prevalent STC and REC were linked to more proteins than PRC and BLC (Fig. 1, Fig. 3A), despite having fewer cases.

**Fig. 3.**
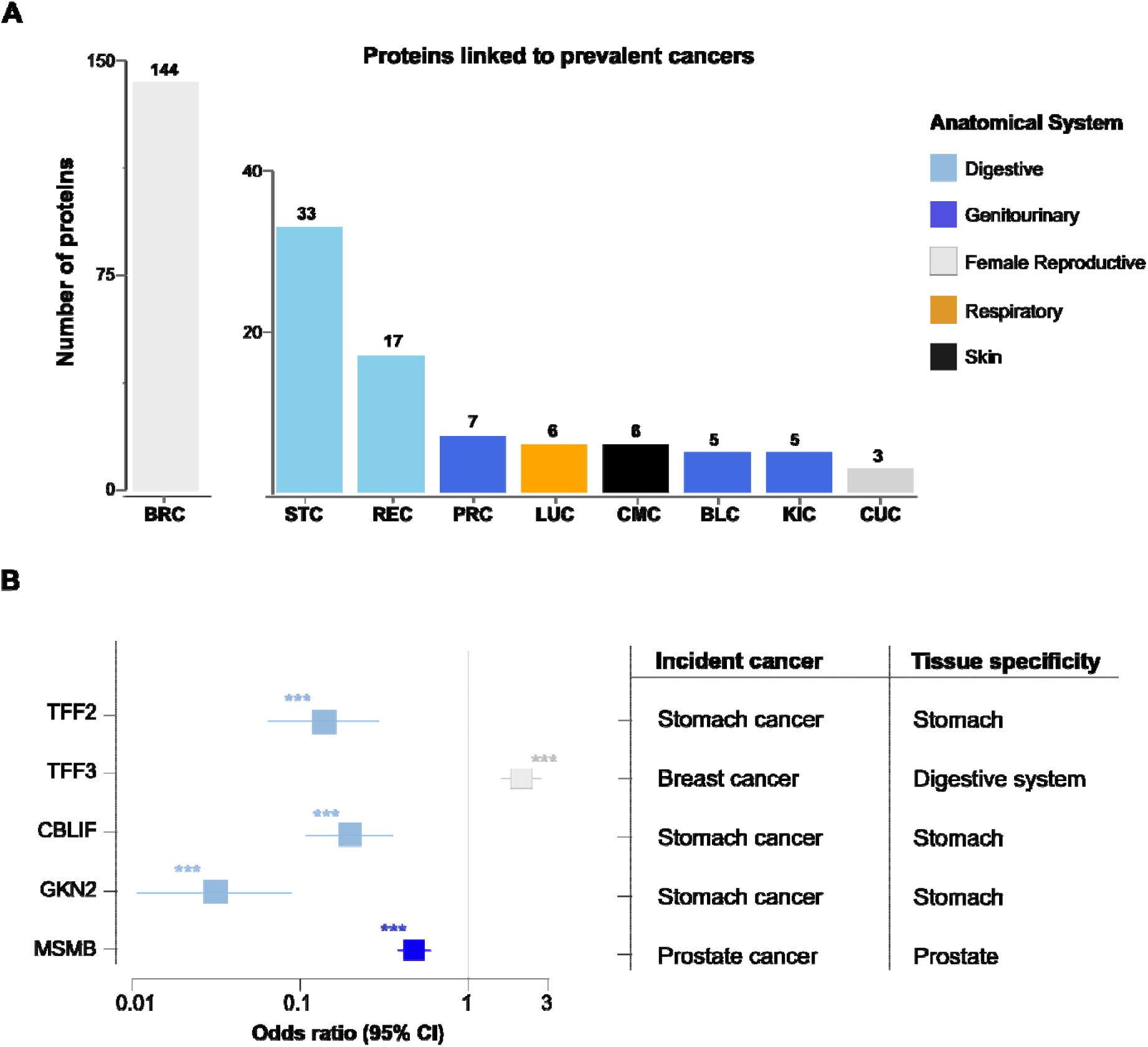
Serum protein associations with prevalent cancer. (**A**) Bar plots illustrating the number of serum proteins associated with different cancer types using abbreviations specified in Fig. 1. Prevalent cancers represent malignancies identified before participant recruitment into the AGES study. Statistical analysis used logistic regression adjusted for age, sex, eGFR, and established risk factors for each cancer type (Supplementary Note 1). Significance testing utilized the Benjamini-Hochberg method for multiple comparisons (FDR < 0.05). Column colors correspond to the anatomical origin system of tumors, with the number of significantly associated proteins displayed above each column. (**B**) Forest plot displaying log2-transformed protein levels (odds ratios) with 95% confidence intervals (CIs) for a selected subset of serum proteins associated with prevalent cancer diagnoses, highlighting tissue-specific patterns. Color-coded data points reflect tumor anatomical origin and indicate statistically significant findings (FDR < 0.05, ***). Horizontal bars represent 95% CIs.

Whereas 13 incident cancer types were analyzed, only nine prevalent cancers had sufficient case numbers or significant associations with serum proteins (Fig. 1, Fig. 3A). Like the findings from the incident cancer analysis, only a few proteins were associated with multiple prevalent cancer types, most notably the trefoil factors TFF2 and TFF3, linked to STC and BRC at FDR < 0.05 (Table S11, Fig. 3B). TFF2 and TFF3 are specifically expressed in mucin-secreting epithelial tissues of the gastric mucosa and gastrointestinal tract^74,75^, respectively, where they contribute to maintaining epithelial integrity^76^. While lower expression of TFF2 in STC suggests a local, tissue-specific role in tumor biology, the higher levels associated with increased risk of BRC (Table S11), may reflect systemic or ectopic effects. Additional four proteins, C7, OMD, LRRC15, and CLIC4, were associated with more than one prevalent cancer type at FDR < 0.05 or <0.10, primarily STC, BRC, PRC, and REC (Table S11). For instance, circulating levels of CLIC4 were positively associated with prevalent STC at FDR < 0.05 (Table S11), and with prevalent BRC and REC at FDR < 0.10 (Table S11). Notably, CLIC4, a protein involved in ion transport and present in both soluble and membrane-bound forms, has been linked to various cancer types, primarily due to its role in the TME and regulation of oxidative stress^77^, with elevated expression serving as a negative prognostic factor for patient survival^78^. The serum proteins C7, OMD, and LRRC15 have also been linked to pathways involved in tumor-host interactions^79–81^. Few proteins showed sex-specific associations with prevalent cancers, likely due to the smaller number of cases and cancer types. Notable exceptions are NR3C2 and IL1R2, linked to prevalent BLC in females and likely reflecting tumor–host interactions via inhibition of angiogenesis or a tumor promoting microenvironment^82,83^, respectively (Supplementary Notes 3 and 7).

Well-established cancer-associated genes encoding proteins like GKN2 and MSMB, which are known to be linked to stomach and prostate cancers^84,85^, respectively, were also associated with the prevalent forms of these cancer types in the current study, with no connection to other prevalent cancers (Table S11, Fig. 3B). Further, the stomach-specific protein CBLIF, whose role in cancer remains unclear, also showed specificity for prevalent STC in this analysis (Table S11, Fig. 3B). These proteins are primarily expressed in the organs where their respective tumors arise: TFF2, GKN2 and CBLIF in the stomach, and MSMB in the prostate. All four proteins exhibit a negative association with their prevalent cancer types in the AGES (Table S11, Fig. 3B). The inverse associations of these proteins with cancers diagnosed prior to the baseline visit and protein measurement may reflect that some participants had previously undergone partial or total gastrectomy or prostatectomy. Supporting the inverse relationship, however, both GKN2 and MSMB have previously been shown to be secreted and downregulated in their corresponding cancers^86,87^. Therefore, treatment alone is unlikely to fully explain the reduced protein levels observed in cancers with a prior diagnosis. As observed in many instances, proteins associated with cancer tend to be specifically expressed in the tissue of tumor origin. The proteins linked to prevalent STC are particularly notable due to their enrichment in the stomach and the broader gastrointestinal tract. More to the point, with 25 cases diagnosed with STC prior to study enrollment and protein measurements (Fig. S8A), a total of 33 serum proteins were significantly associated with prevalent STC at FDR < 0.05 (Fig. S8B). Many of these proteins were exclusively expressed in regions of the digestive system (Fig. S8C-D). Finally, Table S12 summarizes key characteristics of serum proteins associated with incident and/or prevalent cancers, including tissue specificity, tumor-related regulation, and prior genetic links to disease.

### Proteins associated with both incident and prevalent cancer states

A total of 25 protein associations overlapped between incident and prevalent cancer types at FDR < 0.05 (Fig. 4A-C). Among these, WFDC2, CLEC3B, CRTAC1, and KLK3, showed overlap for the same cancer type, whereas others, including trefoil factors (TFF1-3), MET (c-MET), GDF15, EGFR, and MSMB, were associated with different cancer types (Fig. 4C; Tables S10-S11). Interestingly, both MET and EGFR are considered canonical oncogenes^35,36^, playing pivotal roles in tumorigenesis. Both CRTAC1 and GDF15 are implicated in tumor-host interactions through the EMT process^88,89^, whereas the roles of the other overlapping proteins have been discussed above. While the direction of effect was consistent across the disease states (prevalent or incident) for most of these proteins, some proteins, including KLK3, MSMB and the trefoil factors TFF1 and TFF2, exhibited opposite directions of effect depending on the disease state and/or cancer type (Fig. 4B-C, Tables S10-S11). For example, KLK3, which was positively associated with future risk of PRC but showed a negative association in previously diagnosed PRC cases (Fig. 4B). The negative correlation between KLK3 and prevalent PRC may reflect the impact of common treatments including prostatectomy or androgen deprivation therapy, which are known to lower PSA levels^90^, the product of the *KLK3* gene.

**Fig. 4.**
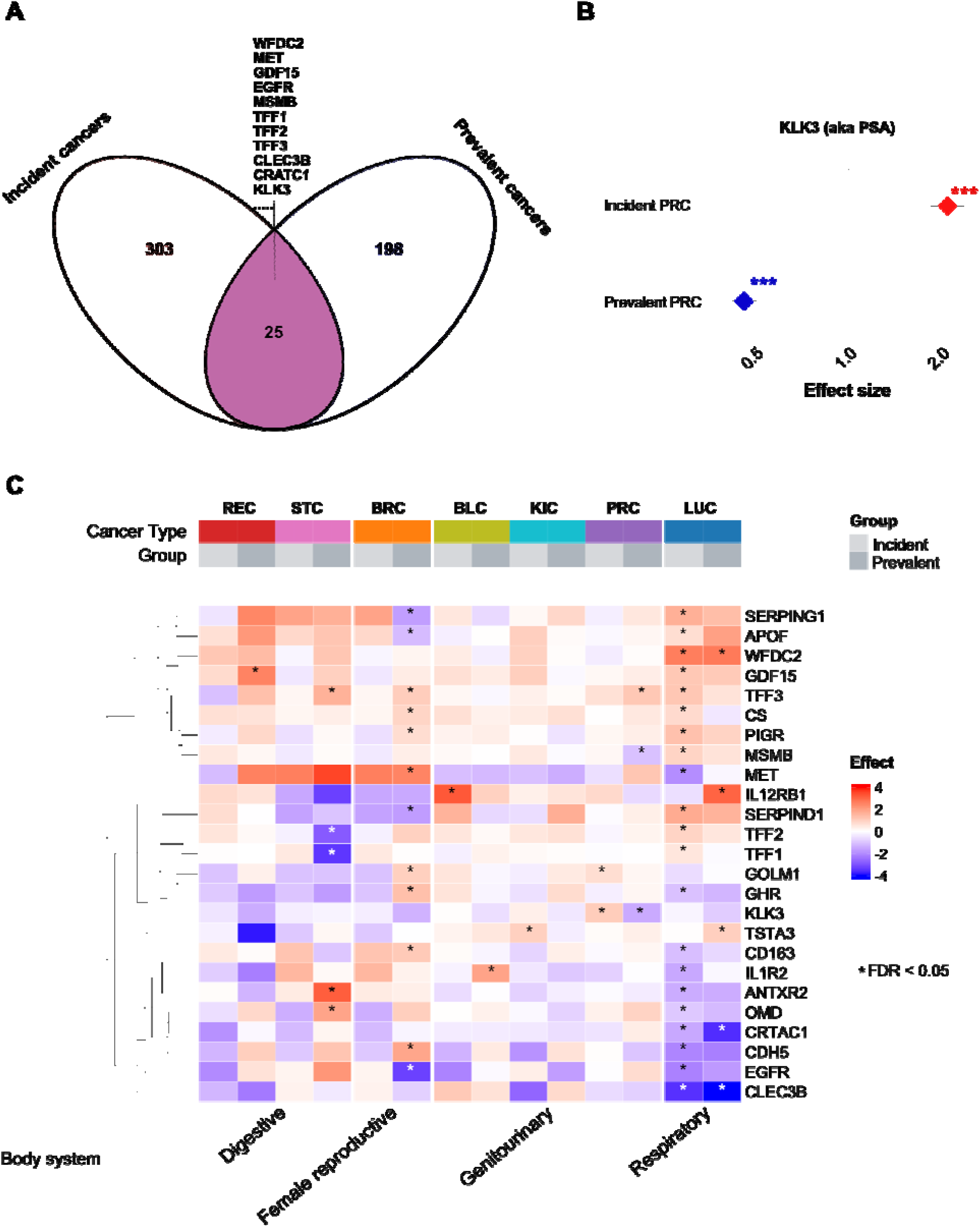
Comparison of serum protein associations between incident and prevalent cancers. (**A**) Venn diagram showing the overlap of proteins significantly associated with incident cancers and prevalent cancers, with 25 proteins shared between both groups. Representative examples of shared proteins are listed, including, for example, the oncogenes EGFR and MET. (**B**) Divergent effects of KLK3 (prostate-specific antigen, PSA) in prostate cancer (PRC), demonstrating contrasting associations between incident and prevalent disease. Effect sizes are shown as hazard ratios for incident PRC and odds ratios for prevalent PRC, with statistical significance indicated by asterisks (***FDR < 0.05). (**C**) Heatmap visualization of associations for 25 shared serum proteins with seven incident and prevalent cancer types: rectal (REC), stomach (STC), breast (BRC), bladder (BLC), kidney (KIC), prostate (PRC), and lung (LUC) cancers. Proteins are hierarchically clustered by regression estimates across body systems (digestive, female reproductive, genitourinary, and respiratory). Effect sizes are color-coded from blue (negative association, protective) to red (positive association, risk-promoting), with asterisks marking statistically significant associations (FDR < 0.05). Gray bars indicate incident versus prevalent cancer groups for each cancer type.

### Analysis of a combined group of patients of any cancer type

While many findings are cancer-type specific, previous research has revealed shared genetic associations across multiple cancer types^91,92^. Since some serum proteins may serve as pan-cancer markers reflecting fundamental cancer biology, and to enhance the power to detect new associations with the 7,523 serum proteins, we pooled cases with a diagnosis of any cancer type (ATC), including the 1,235 previously analyzed cases and an additional 684 cases with cancer diagnosis other than the above 13 types, resulting in 1,916 individuals with ATC (Table S8). Overall, the ATC study group included 835 cases of prevalent cancers (499 females) and 1,081 cases of incident cancers (535 females) (Supplementary Note 8).

Using multiple covariate adjustments as well as sex-specific analyses, a total of 291 proteins (320 aptamers) were associated with incident ATC at an FDR < 0.05 (Table S13, Fig. 5A), while 56 proteins (63 aptamers) were linked to prevalent ATC at the same threshold (Table S14, Fig. 5B). At FDR < 0.10, 449 proteins (489 aptamers) were associated with incident ATC (Table S13), whereas 99 proteins (112 aptamers) were associated with prevalent ATC (Table S14). While many novel protein associations were identified (209 for incident ATC and 32 for prevalent ATC) at FDR < 0.05, many proteins also appeared in analyses of individual incident or prevalent cancer types (Fig. 5C). Of the 291 proteins linked to incident ATC, 104 (36%) overlapped with those identified through analyses of individual cancer types (Fig. 5C). Notable serum proteins, such as WFDC2 and GDF15, which are components of the Cancer Seek blood test panel for early detection of various cancers^19^, were associated with incident ATC in our study as well as with incident LUC (Tables S10 and S13). Similarly, 25 (44%) of the proteins associated with prevalent ATC were also identified in individual cancer analyses (Fig. 5C). Two proteins, KLK3 and TFF3, were identified across all four study groups (Fig. 5C). Consistent with the differing effects observed in incident versus prevalent PRC (Fig. 4B), KLK3 serum quintiles showed opposite associations with incident and prevalent ATC (Fig. S9A), though effect sizes were reduced in the combined ATC group compared to individual cancer types (Fig. S9B). In contrast, TFF3 showed positive associations with multiple cancer types beyond incident and prevalent ATC, including incident LUC and prevalent BRC, STC, and PRC (Tables S10-S11, Fig.4C). These findings align with previous research that associated TFF3 with these and other cancer types^93^.

**Fig. 5.**
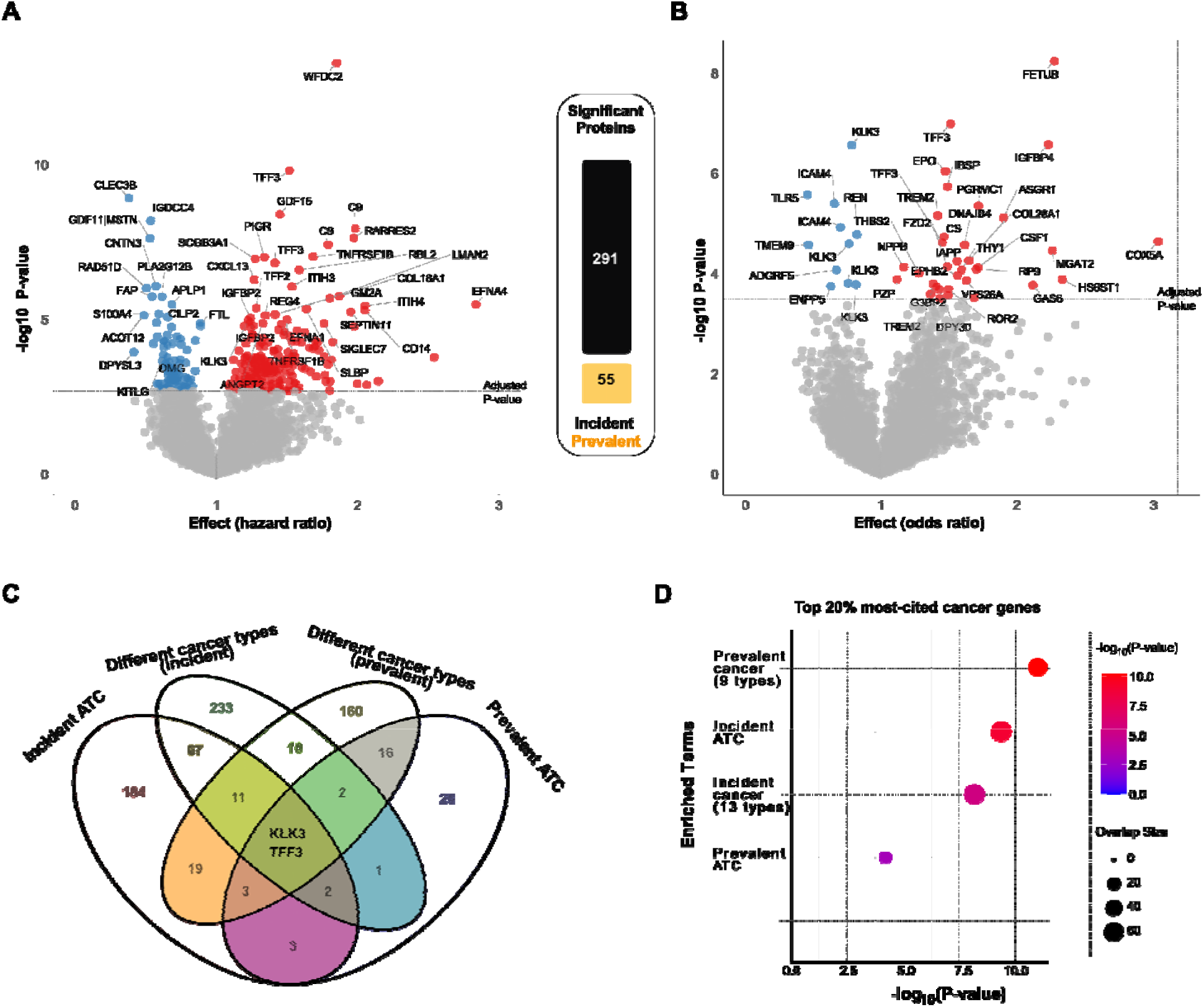
**The relationship between serum proteins and any cancer type**. Volcano plots showing serum protein associations with (**A**) incident (n = 1,081) and (**B**) prevalent (n = 835) cancers of any type (ATC), using standard covariate adjustment. The central panel shows the total number of significantly associated proteins across models with standard and full adjustments, including sex-specific analyses, with bar heights representing the respective counts. Incident cancers show 5.3× more significant protein associations than prevalent cancers. Proteins positively linked to ATC are highlighted in red, while downregulated proteins appear in blue. The plot emphasizes a selected group of proteins; however, any proteins positioned above the dotted horizontal line, representing the adjusted P-value threshold (FDR < 0.05), are considered statistically significant. (**C**) Venn diagram showing the overlap of proteins associated with different patient groups, including incident cancers (pooled from all 13 individual cancer types), incident ATC, prevalent cancers (pooled from the 13 prevalent cancer types), and prevalent ATC. (**D**) The plot illustrates the enrichment of cancer-associated proteins identified in this study among the 20% most highly cited cancer genes in the literature, as described in Methods.

### Integrative multilevel analysis of the serum proteome with cancer genetics, network biology, and existing cancer data

Given the lack of cancer studies with comparable proteomic and phenotype depth, we assessed the overlap between the genes encoding the cancer-associated proteins and top 20% ranked cancer-related genes from the literature (Methods). Our findings show a significant enrichment (hypergeometric test) of the most frequently cited cancer genes: for proteins associated with incident cancer types (*P* = 3×10 ), prevalent cancer (*P* = 4×10 ¹¹), incident ATC (*P* = 3×10 ¹¹), and prevalent ATC (*P* = 8×10 ^5^) (Fig. 5D). These overlaps included well-established cancer genes such as the oncogenes *FGFR1*, *MET*, *EGFR*, and *CTNNB1*, as well as *CXCL8* and *KLK3* highlighted above, and *NCAM1* and *CCK* which have been linked to tumor–host interactions within the TME^94,95^. The proteins encoded by these genes were associated with future cancer risk in the present study. Additionally, several highly cited genes including the oncogenes *RET* and *HSP90AA1*^35,36^, the tumor suppressor *TP53*^35,36^, and others such as *CDH17*^96^, *HSP90AB1*^97^, and *PIK3C3*^98^, which play diverse roles in tumor-host interactions, encode proteins that were associated with prevalent cancer in the current study. In contrast, a similar analysis of the bottom 20% of least-cited cancer-related papers revealed no evidence of enrichment (*P* = 0.967). The considerable overlap between our identified associations and previous gene- and protein-based cancer studies supports the robustness of our findings, while also revealing numerous novel links.

Germline genetic variation shapes circulating protein levels^20,24,31,99^, enabling systematic mapping of cancer risk loci to the serum proteome. By integrating GWAS data for multiple cancers with measurements of 7,523 proteins, we identified 300 variants influencing 737 proteins (Table S15, Supplementary Note 9). Of these, 188 showed proximal *cis*-acting associations across the cancer types examined (Fig. 6A), and demonstrating substantial cross-cancer overlap and revealing several circulating-proteome hotspot loci (Supplementary Note 9). Pan-cancer GWAS further expanded this to 776 genetically influenced proteins (Table S16, Supplementary Note 9), 149 of which are also associated with incident or prevalent cancer (OR = 2.19, *P* = 7×10 ¹ ). Fifteen proteins, including MET, CD163, GOLM1, CDH5, MSMB, SERPIND1,

**Fig. 6.**
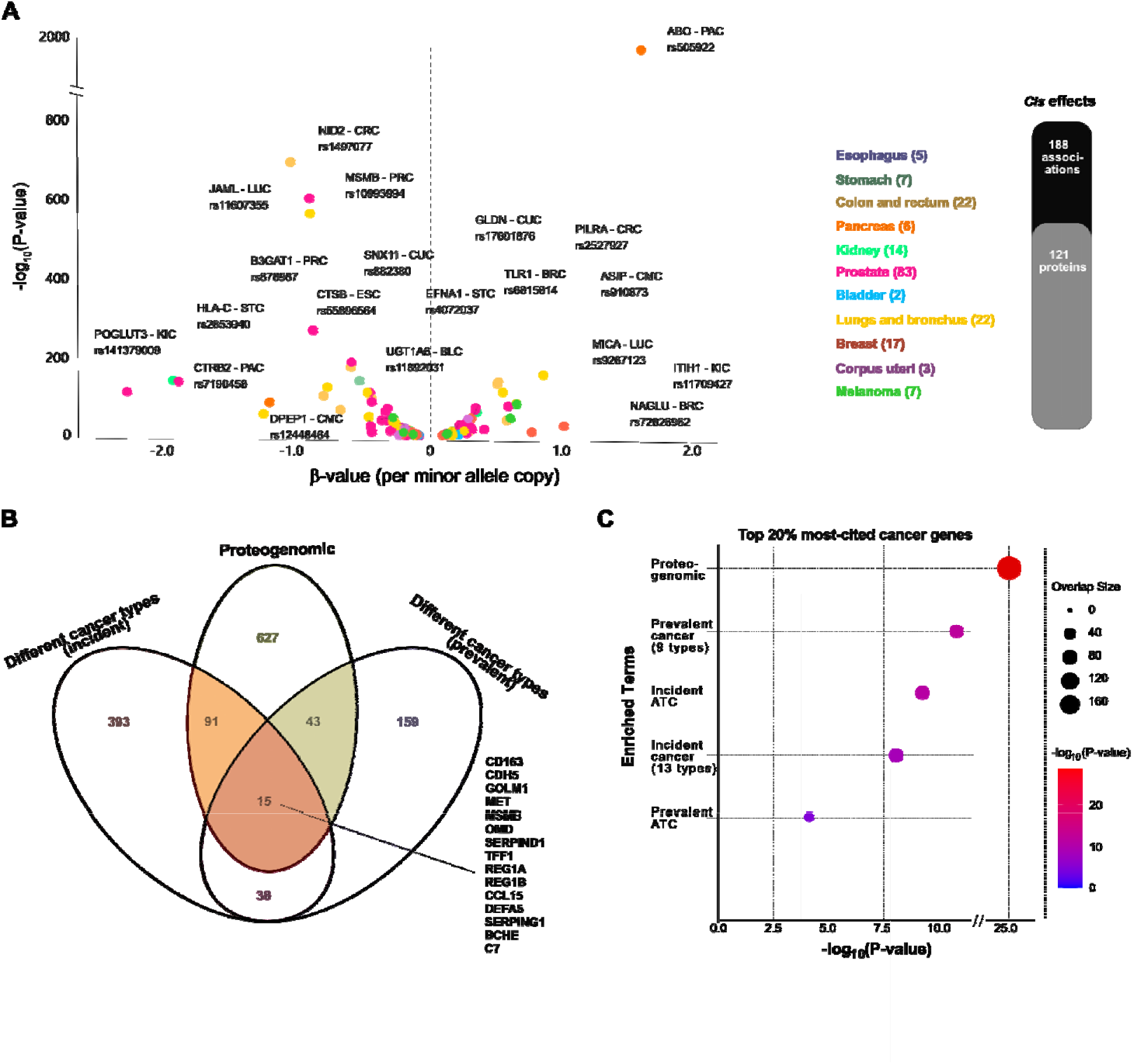
**Serum proteomic signatures of germline cancer risk loci**. (**A)** Volcano plot showing *cis*-regulatory effects of germline cancer risk loci on serum proteins across multiple cancer types (color-coded). The bar plot (right) depicts 188 proximal *cis* associations across cancers, with some variants shared between cancer types, involving 121 distinct proteins. (**B**) Overlap between proteins identified in the observational analyses (incident and prevalent cancers) and those linked to genetic cancer risk through proteogenomic analysis. (**C**) Enrichment analysis showing how proteins from both observational and genetic cancer risk analyses align with top-ranked cancer genes from literature-based gene citation data.

OMD, and TFF1, were consistently supported across observational, genetic, and proteomic analyses (Fig. 6B), several of which are regulated by *cis*-acting genetic variants (Fig. S10). While the roles of MET, MSMB, OMD, and TFF1 in cancer have been discussed above, the remaining proteins are implicated in tumor-host interactions, including immune modulation, EMT and TME, and metastatic processes^80,100–104^. Proteins regulated by cancer-risk loci were also highly enriched among well-established cancer genes (*P* = 9×10 ² ; Fig. 6C).

Finally, we found that proteins associated with individual cancer types, any cancer, or established germline genetic risk factors were significantly enriched within the co-regulatory and causal protein networks originally reconstructed in the AGES cohort^24,105^ (Tables S17-S18, Figs. S11-S12, Supplementary Note 10). Such enrichment indicates that these cancer-linked proteins are not acting in isolation but instead cluster within densely interconnected modules of the proteome, reflecting shared regulation, coordinated biological functions, or participation in common disease-relevant pathways. This network-level organization suggests that cancer risk may be influenced by perturbations to broader proteomic systems rather than single proteins acting alone.

### Causal inference through forward Mendelian randomization analysis

To explore potential causal relationships between circulating proteins and cancer risk, we conducted forward two-sample Mendelian randomization (MR) analyses^106,107^ (Methods). Here, we applied an unbiased, proteome-wide strategy by integrating comprehensive internal *cis*-acting pQTL data (2,062 instruments)^108^, with GWAS meta-analysis summary statistics for genetic liability across multiple cancer types (Table S19, Fig. 7A). This design enabled systematic evaluation of directional causal effects (protein → cancer) without restricting analyses to proteins identified in observational studies. We used the most up-to-date GWAS summary statistics to maximize statistical power (Table S19), except for PAC. To capture a broad set of potentially causal signals, results are reported at FDR < 0.10.

**Fig. 7.**
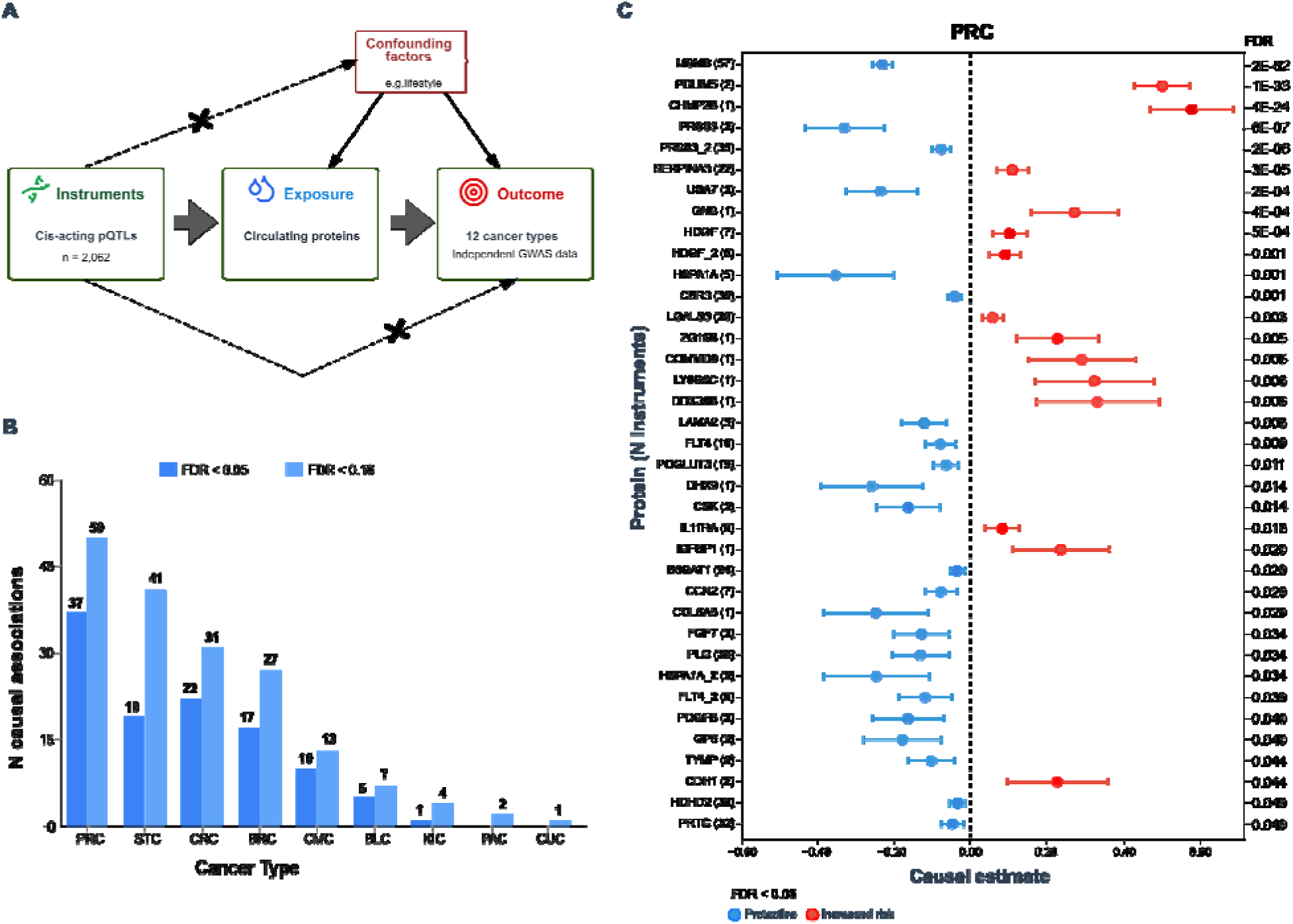
**Proteome-wide forward MR analysis establishing causal links between the circulating proteome and cancer**. (**A**) The diagram illustrates the forward two-sample MR framework used to assess causal effects of circulating proteins on cancer risk across 12 cancer types (CRC includes both colon and rectal cancers). Genetic variants (*cis*-acting pQTLs) from protein GWAS summary statistics served as instruments for 2,062 proteins. The approach relies on three MR assumptions. (1) relevance: genetic instruments are strongly associated with protein levels; (2) independence: variants are not associated with confounders (dotted line and X); and (3) exclusion restriction: variants influence cancer risk only via protein levels (dotted line and X). Although confounders may affect both protein levels and cancer risk, they should be independent of the instruments, allowing unbiased causal inference. Cancer risk estimates were derived from independent GWAS summary statistics for 12 cancer types (Table S19). (**B**) The column plot shows the number of associations with evidence for causal links to one or multiple cancer types at two false discovery rate (FDR) thresholds. Dark blue bars indicate FDR < 0.05, light blue bars indicate FDR < 0.10, and the numbers are displayed atop each bar. (**C**) Forest plot showing significant Mendelian randomization findings for prostate cancer (PRC) (FDR < 0.05), with proteins ordered by the significance of the association (FDR). Red data points denote causal associations with increased disease risk; blue points represent potentially protective effects. Protein annotations appear on the y-axis with the number of genetic instruments shown in parentheses. Repeated protein names indicate independent aptamer measurements with significant causal associations. FDR values for each test are listed on the right.

Across all cancers, we identified 112 putative causal associations at FDR < 0.05 and 176 at FDR < 0.10 (Table S20). The largest numbers of associations (FDR < 0.10) were observed for prostate (PRC, *n* = 50), stomach (STC, *n* = 41), colorectal (CRC, *n* = 31), breast (BRC, *n* = 27), and cutaneous melanoma (CMC, *n* = 13) (Table S20, Fig. 7B). No significant associations were detected for esophageal, ovarian, or lung cancer. Fifteen proteins demonstrated putative causal links across multiple cancer types, and 40 overlapped with proteins identified in observational analyses, 58% of which showed concordant effect directions (Table S20). Among causally implicated proteins, seven correspond to known oncogenes (e.g., *ERBB3*, *GREM1*, *RET*) or tumor suppressors (e.g., *CDH1*, *MST1*)^35,36^. In addition, 56 proteins overlapped with signals from the proteogenomic analysis of cancer risk loci (Table S20). Supplementary Note 11 highlights representative sets of causal candidate proteins across cancer types.

*Cancers of the digestive system*: For CRC, 22 proteins were identified at FDR < 0.05 and 31 at FDR < 0.10 (Table S20, Fig. 7B, Fig. S13). These included established oncogenes, tumor suppressors, and multifunctional molecules with context-dependent effects (Supplementary Note 11). The bone morphogenetic protein antagonist GREM1, which corresponding gene is classified as an oncogene^35,36^, showed the strongest positive causal association with CRC (β = 0.180, FDR = 1×10 ¹²). This finding aligns with previous MR studies suggesting circulating GREM1 mediates the link between adiposity and CRC risk^109^ and with reports implicating *GREM1* variants in the association between physical activity and CRC^110^. Several additional proteins, such as CHRDL2, DARS1, GSS, and GSR, demonstrated directionally consistent causal effects with previous evidence, while LGALS4 and HHIP displayed opposite directions compared with prior functional findings (Supplementary Note 11). For STC we identified 19 at FDR < 0.05 (41 at FDR < 0.10), respectively (Table S20, Fig. 7B, Fig. S14). Several STC-associated proteins had prior links to gastric cancer, including HSPA1A, RET, KLK10, IL6R, TLR3, and IL17RB (Supplementary Note 11).

*Cancers of the genitourinary system*: For PRC, 37 associations were supported at FDR < 0.05 and 50 at FDR < 0.10 (Table S20, Fig. 7B-C). We replicated previously reported MR findings^111^ linking circulating MSMB, PRSS3 (two aptamers), SERPINA3, POGLUT3, and PLG to aggressive or early-onset PRC, with consistent effect directions (Fig. 7C). Notably, POGLUT3 was also associated with KIC and CMC at FDR < 0.10 (Table S20), showing consistent directional effects to that in PRC, justifying the use of slightly relaxed FDR thresholds. Additional PRC-associated proteins, including CDH1, PDLIM5, HDGF, and FGF7, are described in Supplementary Note 11. Our forward MR analysis identified five circulating proteins associated with BLC at FDR < 0.05 and seven at FDR < 0.10 (Table S20, Fig. 7B), including inverse associations of GSTM1, GSTM3, and GSTM4 with BLC risk (Supplementary Note 11), suggesting potential protective effects. The findings for GSTM1 and GSTM4 align with recent GWAS and MR studies of BLC^112^, and in our analysis GSTM1 and GSTM3 were likewise negatively associated with BRC at FDR < 0.10 (Table S20). The *GSTM* genes, clustered on chromosome 1p, encode mu-class glutathione S-transferases that detoxify electrophilic and carcinogenic compounds, including tobacco-derived metabolites^113^. Epidemiological data link the *GSTM1*-null genotype and *GSTM3* intronic polymorphisms to increased BLC susceptibility^113,114^, particularly among smokers. As tobacco smoking remains the predominant BLC risk factor^115^, these findings provide genetic evidence that higher circulating GSTM levels may be protective, highlighting the role of detoxification pathways in bladder carcinogenesis.

*Female reproductive cancers*: For BRC, 17 serum proteins were potentially causally related at FDR < 0.05 and 27 at FDR < 0.10 (Table S20, Fig. 7B, Fig. S15). We replicated three of five plasma proteins previously reported as causally linked to BRC^116^, including a positive association for CTSF and potentially protective effects for SNUPN and PARK7 (Table S20, Fig. S15, Supplementary Note 11). Intriguingly, in that study^116^, CSK was reported to be negatively and causally associated with PRC, whereas in the current analysis it showed directionally consistent associations with both CRC and PRC (Table S20). Additional proteins with directionally consistent evidence from prior functional studies included ANXA4, TAGLN, and UBA7 (Supplementary Note 11).

*Skin cancer*: For cutaneous melanoma (CMC), our forward MR analysis identified 10 circulating proteins associated at FDR < 0.05 and 13 at FDR < 0.10 (Table S20, Fig. 7B, Fig. S16). The strongest signals were observed for ASIP, ERBB3, and ARG1, all of which have established links to melanoma biology (Supplementary Note 11). ASIP showed the most robust causal association, consistent with our previous proteogenomic findings implicating distant regulatory variants of *ASIP* in melanoma risk and its known role in pigmentation pathways and melanoma susceptibility^22^. ERBB3, frequently hyperactivated in melanoma^117,118^, also displayed a positive causal association. ARG1, a marker of immunosuppressive tumor-associated macrophages^119,120^, similarly showed a positive association with CMC. Additional proteins included RNASET2, which was positively associated with CMC despite its tumor-suppressive functions in other contexts, and IRF3, which demonstrated a negative, potentially protective association (Fig. S16–S17, Supplementary Note 11).

Given extensive pathway-level convergence across cancers¹² , we conducted STRING-based analyses^121^ of all proteins showing putative causal associations at FDR < 0.05. This revealed significant enrichment of functional and physical protein-protein interactions (PPI) among cancer-associated proteins (115 observed vs. 53 expected edges; *P* = 9 × 10 ¹ ; Fig. S17). K-means clustering revealed 11 PPI clusters, many of which were interconnected (Fig. S17), encompassing key biological processes, including extracellular matrix remodeling (laminins/nidogen/fibulins), constitutive PI3K/VEGF signaling in cancer, cytokine and ligand-receptor communication, glutathione-dependent redox regulation, and RNA helicase–mediated control of translation. Crosstalk among these modules likely contributes to tumor cell survival, angiogenesis, immune modulation, and metastatic progression (Supplementary Note 11). Further details on selected findings from the MR analysis of serum proteome associations across cancer types are provided in Supplementary Note 11. Finally, Table S21 summarizes key characteristics of the cancer-associated proteins highlighted in the main text above.

## Discussion

Cancer is a complex and heterogeneous disease marked by uncontrolled cell growth, invasion to surrounding tissues, and metastasis. Despite significant advances in treatment, cancer remains the second leading cause of death globally, highlighting the urgent need for better strategies in early detection and prevention. This study aimed to identify serum proteins associated with both previously diagnosed and subsequently diagnosed cancers across 13 distinct cancer types, with the purpose of facilitating early detection and to provide insights into the molecular mechanisms driving tumorigenesis. Among the many cancers examined, this study includes some of the most aggressive and treatment-resistant types, such as esophageal, gastric, pancreatic, and ovarian cancers, which often lack effective early diagnostic tools and are diagnosed at advanced stages, contributing to high mortality rates. In summary, this study identified hundreds of proteins associated with diverse cancer types, both incident and prevalent, as well as proteins linked to germline genetic risk factors and potential causal candidates.

### Serum proteins linked to past diagnoses and future cancer risk

In the prospective, population-based AGES study, serum levels of 7,523 proteins, covering more than 30% of all annotated human protein-coding genes, were quantified in 5,376 participants. More than half of all cancers are diagnosed in individuals aged 65 or older, the entry age for the AGES study, and this burden is projected to grow significantly as the global population ages^7,122^. The contributing factors to cancer in older adults may differ from those in younger individuals, due to multiple age-related changes, including impaired DNA repair, chronic inflammation, cellular senescence, and cumulative exposure to carcinogens^123^. The current study identified hundreds of protein-cancer associations, with most proteins showing specificity to individual cancer types and only limited overlap between those associated with prevalent and incident cancers. Nevertheless, several proteins were associated with two or more cancer types or timing, suggesting some shared molecular pathways across different cancers. It is important to note that the prevalent cancer cases in our study refer to individuals who had received a cancer diagnosis prior to enrollment in the AGES study. As such, this group likely includes a heterogeneous mix of clinical states, ranging from patients who may have undergone successful treatment with no current tumor burden, to those with metastatic disease or experiencing relapse. This diversity in disease stage, treatment history, and tumor activity at the time of sampling introduces biological variability that should be considered when interpreting the associations between protein levels and prevalent cancer.

### Proteins associated with genetic susceptibility loci

Large-scale GWAS meta-analyses have identified numerous genetic loci associated with elevated cancer risk that are considered causal drivers of cancer onset, though their individual effect sizes are typically weak or modest. In our dataset, 776 serum proteins were regulated in *cis* or *trans* by these susceptibility loci, including 149 that overlapped with proteins associated with prevalent or incident cancers, suggesting that some of these proteins could play a causal role in tumorigenesis. Attention should be given to proteins associated with future cancers and those already implicated as oncogenes or tumor suppressors in literature. Notably, several proteins influenced by susceptibility loci include well-known oncogenes such as *MET* and tumor suppressors like *TP53*. The finding that subtle impact of expression changes in oncogenes or tumor suppressors is associated with cancer risk supports their functional relevance. This contrasts with the more familiar model of tumorigenesis driven by somatic mutations with large functional impacts. In earlier work, we proposed that an individual’s non-cancer state can be represented as a point in high-dimensional gene expression space, with tumorigenesis conceptualized as movement through this space toward a tumor state^124^. In this framework, the Euclidean distance between normal and tumor states reflects the likelihood of tumor development, with shorter distances being more probable. The present findings, linking susceptibility loci to expression shifts in key proteins, support this model, suggesting these changes may collectively modulate the probability of cancer progression.

### Causal associations between circulating proteins and cancer risk

The forward MR analyses comprehensively and unbiasedly evaluated the potential causal effects of circulating proteins across multiple cancer types, utilizing proteome-wide *cis*-pQTL instruments alongside the most up-to-date GWAS summary statistics. Across cancer types, we identified numerous putative causal associations, both protective and risk-enhancing, with the largest numbers observed for prostate, stomach, colorectal, and breast cancers, as well as cutaneous melanoma. Many of our findings replicated recent reports for specific cancer types, corroborating emerging signals from proteomic studies conducted over the past year. Key oncogenes and tumor suppressors, such as ERBB3, GREM1, RET, CDH1, and MST1, were among the proteins implicated, highlighting known tumor biology. Network analysis revealed strong pathway-level convergence, with clusters related to extracellular matrix remodeling, PI3K/VEGF signaling, immune modulation, redox regulation, and translational control. These results illustrate the breadth and mechanistic plausibility of these circulating causal proteome cancer links. Overall, these results provide robust, systematic support for causal roles of circulating proteins in cancer etiology and reinforce the value of integrating proteomic and genetic evidence in uncovering novel cancer biomarkers and therapeutic targets.

### Toward an integrative functional framework of serum proteins in cancer biology

Our findings reveal key biological mechanisms underlying cancer etiology, reflected by the diverse set of serum proteins associated with incident and prevalent cancers (Table S21). To interpret these results, we propose a framework that groups cancer-associated proteins into four biologically informed categories: (1) tumor-specific proteins, including established oncogenes and tumor suppressors; (2) tissue-specific proteins, reflecting the tissue of origin; (3) proteins linked to germline genetic susceptibility, associated with common low-penetrance cancer risk variants; and (4) tumor-host interaction proteins, encompassing lifestyle- or systemic factors, TME, and processes such as EMT. This framework is illustrated in Fig. 8, with representative examples highlighting the breadth of cancer biology captured by serum proteomics. Notably, many proteins linked to cancer in our study fall into the tumor-specific category. The genes encoding these proteins include 24 oncogenes and 11 tumor suppressor genes^35,36^ among those associated with incident or prevalent cancers (Tables S10-S11 and S13-S14), such as *EGFR*, *MET*, *RET*, *FGFR1*, and *CTNNB1*, as well as *CREBBP*, *ARID1A*, *TP53*, and *CDH1*. Similarly, among genes encoding proteins linked to cancer susceptibility loci or putatively causal proteins, 41 and 10, respectively, are classified as oncogenes or tumor suppressors (Tables S15 and S20). These results demonstrate that serum proteomics not only capture well-established cancer pathways but also reveals previously unrecognized protein-cancer relationships.

**Fig. 8.**
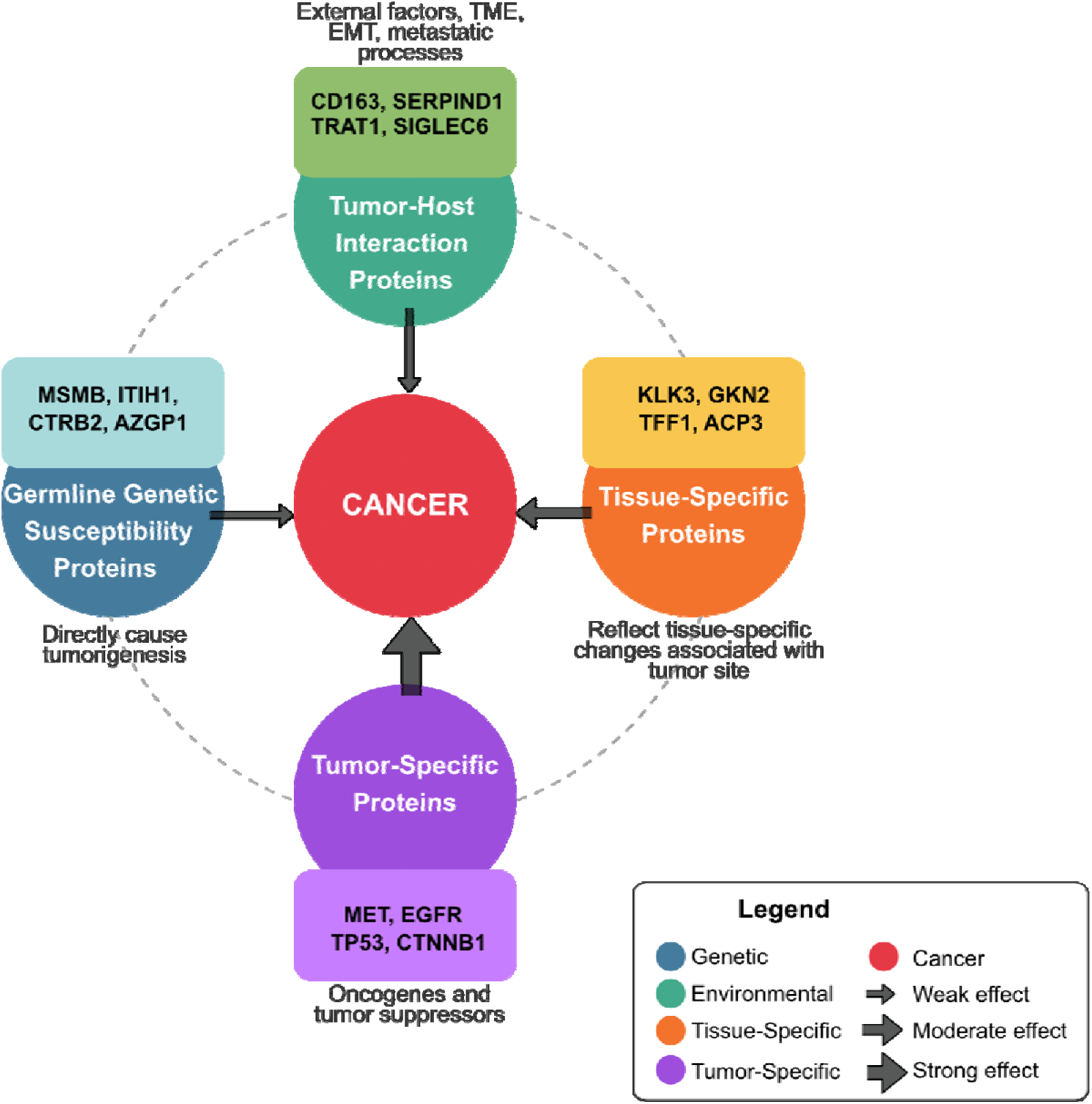
Classification of cancer-associated serum proteins identified in this study. The proteins are grouped into four main categories. 1. Tumor-specific proteins, encoded by oncogenes or tumor suppressor genes. 2. Tissue-specific proteins, reflecting alterations related to the tumor’s tissue of origin. 3. Genetic susceptibility proteins, associated with common low-penetrance germline variants. 4. Tumor-host interaction proteins, a broad category encompassing proteins that do not fit neatly into the other groups, representing the complex interplay between the tumor and its host environment. This includes proteins influenced by lifestyle factors (e.g., smoking) and systemic conditions (such as inflammation or metabolic dysfunction), as well as those involved in tumor-intrinsic processes like the tumor microenvironment (TME), which originates from stromal cells, immune cells, or the extracellular matrix, and epithelial-mesenchymal transition (EMT), linking tumor cell behavior to microenvironment remodeling. This category also includes proteins implicated in metastatic progression. Representative cancer-associated proteins are highlighted within each category. A comprehensive categorization of all cancer-associated proteins discussed in the main text is provided in Table S21.

The strong convergence between our proteomic findings and known oncogenic or tumor-suppressive processes provides a biologically coherent link between circulating proteins and intracellular tumor biology. Although genes such as *TP53*, *CDH1*, *CREBBP*, *MET*, *RET*, *EGFR*, and *CTNNB1* are well-characterized in tumor tissue, their detection as circulating proteins is far less explored and may reflect secretion, shedding, or broader tumor-host interactions. Importantly, several such proteins were identified with links to incident cancers, suggesting that systemic proteomic alterations preceded clinical detection and may offer mechanistically informed biomarkers for early surveillance. This supports the concept that serum proteins can report on coordinated regulatory processes across tissues, including inter-tissue signaling and systemic physiological responses^24,125^, providing a window into preclinical cancer biology. Our integrative framework therefore highlights how tumor-specific, tissue-specific, genetic susceptibility-linked, and tumor-host interaction proteins together map a multidimensional landscape of cancer biology that may help prioritize candidates for functional follow-up and translational development.

### Early detection and targeting of mechanisms driving pre-clinical tumorigenesis

The identification of dysregulated proteins years before the cancer diagnosis suggests that early detection is achievable. While current methods like circulating tumor DNA (ctDNA) analysis show promise, they are limited by low abundance in early-stage disease^126^. In contrast, proteomic biomarkers could potentially surpass existing techniques by providing deeper insights into early tumor biology. The presentation of cancer-associated proteins within key oncogenic pathways further indicates the potential for developing early intervention strategies.

Proteins associated with both current and future cancers not only inform us about the nature of these tumors but also suggest potential drivers of tumorigenesis, raising the possibility of intervention before clinical presentation. An intriguing question is whether these same proteins, linked to prevalent or incident cancers, could provide valuable information for patients’ post-surgery or treatment. For instance, could early indicators of relapses be detected? Although our current data does not address this, it presents an interesting avenue for future investigation. Many cancer-associated proteins are linked to germline risk variants and integrating genetic and proteomic profiling could enable long-term risk stratification. Combining genetic risk assessment with proteomic monitoring could help identify individuals who would benefit from more intensive surveillance or preventive therapies. A multi-omics approach that includes susceptibility markers, ctDNA, and serum proteins should be explored to enhance the identification of at-risk individuals before clinical presentation. This approach could open the door to more intensive monitoring and early interventions, such as surgery or therapy.

Circulating protein biomarkers represents a powerful tool for early cancer detection, risk prediction, and understanding the mechanisms underlying tumorigenesis. By leveraging susceptibility loci, environmental factors, and tissue-specific signatures, these biomarkers have the potential to revolutionize cancer screening and prevention. Future research should focus on validating these proteins in prospective cohorts, integrating intensive imaging and monitoring, and developing targeted interventions to intercept cancer at its earliest stages. In conclusion, our findings highlight the potential of circulating proteins as biomarkers of tumor dynamics, which may reflect changes in tumor growth, the microenvironment, and systemic mediators. These insights may lay the foundation for formulating hypotheses and directing future research, with significant implications for early cancer detection and future risk assessment.

## Methods

### Study population

Cohort participants aged 66 through 96 years at the time of blood collection were from the AGES study^32^, a single-center, prospective, population-based study of older adults (N = 5,764, mean age 76.6±6 years). The AGES study was formed between 2002 and 2006, and its participants were randomly selected from the surviving members of the established 40-year-long population-based prospective Reykjavik study^127,128^, with a 72% recruiting rate. The Reykjavik Study, a prospective cardiovascular survey, recruited a random sample of 30,795 adults born between 1907 and 1935 who lived in the greater Reykjavik area in 1967, and were examined in six phases from 1967 to 1996^127,128^. Measurements in the AGES study, including, for example, brain and vascular imaging, are designed to assess four biologic systems: vascular, neurocognitive (including sensory), musculoskeletal, and body composition/metabolism.^32^ All participants are of European ancestry, and a decade-long collaboration with large genetic and epidemiology consortia of multiple disease-related phenotypes revealed no discernible difference between the Icelandic population and other European ancestry cohorts^129–131^. This study was approved (approval number VSN-00-063) by the National Bioethics Committee in Iceland, which serves as the Icelandic Heart Association’s institutional review board in accordance with the Helsinki Declaration, and by the US National Institutes of Health, National Institute on Aging Intramural Institutional Review Board, with all participants providing informed consent.

Prevalent cancer cases were those with a history of cancer at the baseline visit, while the incident malignancies were diagnosed after the first visit with up to 13.6-year follow-up based on hospital records and cancer registries. Diagnoses were based on ICD-10 codes from the 10th revision of the WHO International Classification of Diseases: esophagus (C15), stomach (C16), colon (C18), rectum (C20), pancreas (C25), prostate (C61), kidney (C64), bladder (C67), breast (C50), corpus uteri (C54), ovaries (C56), lung and bronchus (C34), and malignant melanoma (C43). Systolic and diastolic blood pressure were measured twice with subjects in a supine position using a Mercury sphygmomanometer. We categorized smoking status as never smoked, former smoker, or current smoker, while alcohol consumption was determined as units per week. Height was measured in meters, while body mass index (BMI) was expressed in kg/m². The estimated glomerular filtration rate (eGFR) was estimated according to the Chronic Kidney Disease Epidemiology Collaboration equation^132^. Finally, the present study included only individuals who had their serum proteome measured (see below), which amounted to 5,376 AGES participants.

### Proteomics profiling assay

Blood samples were collected at the AGES baseline visit after an overnight fast, and serum samples prepared using a standardized protocol and stored in 0.5mL aliquots at -80°C. Serum samples collected from the inception period, i.e., from 2002 to 2006, were used to generate proteomics data in the current study. Before the protein measurements, all serum samples from this period went through their first freeze-thaw cycle. Serum protein levels from 5,376 AGES study participants were quantified using the multiplex SomaScan v4.1 proteomic platform^133^, which uses modified DNA aptamers designed to bind target proteins with high affinity and specificity. In total, 7,523 aptamers corresponding to 6,586 UniProt protein identifiers were measured across 8,592 samples (two time points). Thus, some proteins were targeted by more than one aptamer. In such cases, individual aptamers had distinct binding sites (epitopes) or binding affinity^24^. Examples include duplicate aptamers targeting single-pass transmembrane proteins (one binding the extracellular domain and another the intracellular loop), aptamers targeting multimers (e.g., interleukins), and duplicate aptamers produced in distinct expression systems. Of the 7,523 aptamers, 233 were derived from mouse-human chimeras, intended to target proteins from both species. The SOMAmer-based platform measures proteins with femtomole (fM) detection limits and a broad detection range (>8-log dynamic range) of concentration^134^. To avoid batch or time of processing biases, the order of sample collection and separate sample processing for protein measurements were randomized, and all samples run as a single set at SomaLogic Inc. (Boulder, CO, US). All aptamers that passed quality control exhibited mean and median intra-assay and inter-assay coefficients of variation (CV) below 4% at both time points, measured five years apart. Hybridization controls were used to correct systematic variability in detection and calibrator samples of three dilution sets (20% (1:5), 0.5% (1:200), and 0.005% (1:20,000)) were included so that the degree of fluorescence was a quantitative reflection of protein concentration. The adaptive normalization by maximum likelihood (ANML) method was employed to normalize QC replicates and samples using point and variance estimations from a normal U.S. population. The aptamers demonstrated consistent target specificity, as indicated by direct validation through mass spectrometry and/or indirect supporting evidence^24^.

### Genotype data and the identification of cis-acting pQTLs

The genotype data includes assayed and imputed genotype data for 5,661 AGES participants^20^. The genotyping arrays used were Illumina Hu370CNV and Illumina GSA BeadChip, which were quality controlled by eliminating variants with call rates <95% and HWE *P* < 1 × 10^−6^. Both arrays were imputed to the Trans-Omics for Precision Medicine (TOPMed r2) imputation panel^135^ and post-imputation quality control was performed separately for each platform. Variants with a call rate <95% or Hardy-Weinberg equilibrium (HWE) P-value <1×10 were removed prior to imputation. After imputation quality control (*r*^2^<0.7, minor allele frequency < 0.01, HWE P <1×10 ), 11,567,385 single nucleotide polymorphisms (SNPs) for 5,368 individuals were available for the analysis^108^. These variants were linked to each aptamer on the v4.1-7K serum protein panel to identify cis (proximal) pQTLs within a ±500 kb window, resulting in 2,062 proteins with at least one *cis*-acting instrument. These were then tested using a forward MR framework, as described in more detail previously^108^.

### Statistical analysis

Before the analyses, protein data were transformed using a log2 scale, and extreme outlier values excluded, defined as values above the 99.5th percentile of the distribution of 99th percentile cutoffs across all proteins. The relationship between serum protein levels and prevalent cancer was examined cross-sectionally using logistic regression analysis, while the associations of serum proteins with incident cancer were assessed longitudinally via the Cox proportional-hazards model.

A Geneshot^136^ search using the term ’cancer gene’ returned 9,952 entries (∼50% of all human protein-coding genes), each with at least one supporting publication and ranked by publication count. We performed a hypergeometric test to assess the enrichment of genes encoding proteins linked to incident and/or prevalent cancers, within the top 20% of the ranked cancer genes.

We performed forward two-sample Mendelian randomization (MR) analyses^106,107^, to investigate potential causal relationships between serum protein levels and a range of cancer phenotypes. This analysis was extended to include all serum proteins with well-defined *cis*-acting pQTL instruments, irrespective of their observational associations. Summary statistics for genetic liability to multiple cancer types were obtained from publicly available GWAS resources (Table S19). Forward MR analyses (protein → cancer) were conducted using significant *cis-*region SNPs within 500 kb of the start and stop positions for the cognate encoding gene as instruments, applying window-wide significance thresholds^21^, linkage disequilibrium (LD) clumping (r² < 0.2), and instrument strength filters (F-statistic > 10). All SNP sets were harmonized using the TwoSampleMR R package^42^. Multi-SNP causal estimates were derived using generalized weighted least squares (GWLS) to account for LD^21,137^, while Wald ratio estimators were applied for single-variant instruments. Results meeting an FDR threshold of <0.05 were considered putative causal associations.

## Data Availability

Data generated in this study can be obtained from the authors upon reasonable request.

## Acknowledgement

The authors acknowledge the contribution of the Icelandic Heart Association (IHA) staff to the AGES study, as well as the involvement of all study participants. The proteomics work was carried out in collaboration with Novartis Biomedical Research (NIBR). We thank the Research Fund of the Icelandic Cancer Society for supporting this study through funding for V.E. The National Institute on Aging (NIA) contracts N01-AG-12100 and HHSN271201200022C for V. G. financed the study. V. G. received funding from the NIA (1R01AG065596-01A1), and IHA received a grant from the Icelandic Parliament. T.M. acknowledges support from the Research Council of Norway (project number 312045), the European Union Horizon Europe (European Innovation Council) (grant agreement number 101115381), and the L. Meltzers Hoyskolefond.

## References

1. Tomasetti, C. & Vogelstein, B. Cancer etiology. Variation in cancer risk among tissues can be explained by the number of stem cell divisions. Science 347, 78–81 (2015).

2. Hanahan, D. & Weinberg, R.A. Hallmarks of cancer: the next generation. Cell 144, 646–674 (2011).

3. Frank, S.A. Genetic predisposition to cancer - insights from population genetics. Nat Rev Genet 5, 764–772 (2004).

4. Wu, S., Powers, S., Zhu, W. & Hannun, Y.A. Substantial contribution of extrinsic risk factors to cancer development. Nature 529, 43–47 (2016).

5. Hanahan, D. Hallmarks of Cancer: New Dimensions. Cancer Discov 12, 31–46 (2022).

6. Siegel, R.L., Miller, K.D., Fuchs, H.E. & Jemal, A. Cancer statistics, 2022. CA Cancer J Clin 72, 7–33 (2022).

7. Bray, F., et al. Global cancer statistics 2022: GLOBOCAN estimates of incidence and mortality worldwide for 36 cancers in 185 countries. CA Cancer J Clin 74, 229–263 (2024).

8. Swanton, C., et al. Embracing cancer complexity: Hallmarks of systemic disease. Cell 187, 1589–1616 (2024).

9. Qing, T., et al. Germline variant burden in cancer genes correlates with age at diagnosis and somatic mutation burden. Nat Commun 11, 2438 (2020).

10. Antoniou, A., et al. Average risks of breast and ovarian cancer associated with BRCA1 or BRCA2 mutations detected in case Series unselected for family history: a combined analysis of 22 studies. Am J Hum Genet 72, 1117–1130 (2003).

11. Gudmundsson, J., et al. Frequent occurrence of BRCA2 linkage in Icelandic breast cancer families and segregation of a common BRCA2 haplotype. Am J Hum Genet 58, 749–756 (1996).

12. Turnbull, C., Sud, A. & Houlston, R.S. Cancer genetics, precision prevention and a call to action. Nat Genet 50, 1212–1218 (2018).

13. Martincorena, I., et al. Universal Patterns of Selection in Cancer and Somatic Tissues. Cell 171, 1029–1041.e1021 (2017).

14. Vogelstein, B., et al. Cancer genome landscapes. Science 339, 1546–1558 (2013).

15. Baylin, S.B. & Jones, P.A. A decade of exploring the cancer epigenome - biological and translational implications. Nat Rev Cancer 11, 726–734 (2011).

16. Shieh, Y., et al. Population-based screening for cancer: hope and hype. Nature reviews. Clinical oncology 13, 550–565 (2016).

17. Duffy, M.J. Tumor markers in clinical practice: a review focusing on common solid cancers. *Medical principles and practice : international journal of the Kuwait University*, Health Science Centre 22, 4–11 (2013).

18. Chen, X., et al. Non-invasive early detection of cancer four years before conventional diagnosis using a blood test. Nat Commun 11, 3475 (2020).

19. Cohen, J.D., et al. Detection and localization of surgically resectable cancers with a multi-analyte blood test. Science 359, 926–930 (2018).

20. Gudjonsson, A., et al. A genome-wide association study of serum proteins reveals shared loci with common diseases. Nature Communications 13, 1–13 (2022).

21. Emilsson, V., et al. A proteogenomic signature of age-related macular degeneration in blood. Nat Commun 13, 3401 (2022).

22. Emilsson, V., et al. Coding and regulatory variants are associated with serum protein levels and disease. Nature Communications 13, 1–11 (2022).

23. Lamb, J.R., Jennings, L.L., Gudmundsdottir, V., Gudnason, V. & Emilsson, V. It’s in Our Blood: A Glimpse of Personalized Medicine. Trends Mol Med (2020).

24. Emilsson, V., et al. Co-regulatory networks of human serum proteins link genetics to disease. Science 361, 769–773 (2018).

25. Emilsson, V., Gudnason, V. & Jennings, L.L. Predicting health and life span with the deep plasma proteome. Nature medicine 25, 1815–1816 (2019).

26. Williams, S.A., et al. Plasma protein patterns as comprehensive indicators of health. Nat Med 25, 1851–1857 (2019).

27. Lehallier, B., et al. Undulating changes in human plasma proteome profiles across the lifespan. Nat Med 25, 1843–1850 (2019).

28. Axelsson, G.T., et al. The Proteomic Profile of Interstitial Lung Abnormalities. American journal of respiratory and critical care medicine 206, 337–346 (2022).

29. Gudmundsdottir, V., et al. Circulating protein signatures and causal candidates for type 2 diabetes. Diabetes 69, 1843–1853 (2020).

30. Emilsson, V., et al. Proteomic prediction of incident heart failure and its main subtypes. Eur J Heart Fail 26, 87–102 (2024).

31. Sun, B.B., et al. Plasma proteomic associations with genetics and health in the UK Biobank. Nature 622, 329–338 (2023).

32. Harris, T.B., et al. Age, Gene/Environment Susceptibility-Reykjavik Study: multidisciplinary applied phenomics. Am J Epidemiol 165, 1076–1087 (2007).

33. Cox, D.R. Regression Models and Life-Tables. 34, 187–202 (1972).

34. Benjamini, Y. & Hochberg, Y. Controlling the False Discovery Rate: A Practical and Powerful Approach to Multiple Testing. Journal of the Royal Statistical Society **Vol.**57, No. 1: 289–300 (1995).

35. Suehnholz, S.P., et al. Quantifying the Expanding Landscape of Clinical Actionability for Patients with Cancer. Cancer Discov 14, 49–65 (2024).

36. Chakravarty, D., et al. OncoKB: A Precision Oncology Knowledge Base. JCO Precis Oncol 17, 16 (2017).

37. Tsherniak, A., et al. Defining a Cancer Dependency Map. Cell 170, 564–576.e516 (2017).

38. Nunes, J., et al. Siglec-6 as a therapeutic target for cell migration and adhesion in chronic lymphocytic leukemia. Nat Commun 15, 5180 (2024).

39. Yang, Z., et al. Effect of MPP2 and its DNA methylation levels on prognosis of colorectal cancer patients. World journal of surgical oncology 22, 290 (2024).

40. Lu, C., et al. Analysis of Circulating Immune Subsets in Primary Colorectal Cancer. Cancers 14(2022).

41. Shi, S., et al. TMEM106A transcriptionally regulated by promoter methylation is involved in invasion and metastasis of hepatocellular carcinoma. Acta biochimica et biophysica Sinica 54, 1008–1020 (2022).

42. Fan, B., Wang, L. & Wang, J. RAB22A as a predictor of exosome secretion in the progression and relapse of multiple myeloma. Aging 16, 4169–4190 (2024).

43. Pierro, E.W., et al. Trem2 deficiency attenuates breast cancer tumor growth in lean, but not obese or weight loss, mice and is associated with alterations of clonal T cell populations. bioRxiv (2024).

44. Schroder, A.K., et al. miR-379-5p affects breast cancer cell behavior by targeting UBE2E3 ubiquitin conjugating enzyme. PLoS One 19, e0310315 (2024).

45. Schwenk, J.M., et al. The Human Plasma Proteome Draft of 2017: Building on the Human Plasma PeptideAtlas from Mass Spectrometry and Complementary Assays. J Proteome Res 16, 4299–4310 (2017).

46. Fisher, R.C., et al. Disrupting Inflammation-Associated CXCL8-CXCR1 Signaling Inhibits Tumorigenicity Initiated by Sporadic- and Colitis-Colon Cancer Stem Cells. *Neoplasia (New York*, N.Y.) 21, 269–281 (2019).

47. Fang, S., et al. CXCL8 Up-Regulated LSECtin through AKT Signal and Correlates with the Immune Microenvironment Modulation in Colon Cancer. Cancers 14(2022).

48. Srinivasan, S., et al. A PSA SNP associates with cellular function and clinical outcome in men with prostate cancer. Nat Commun 15, 9587 (2024).

49. Veeramani, S., et al. Cellular prostatic acid phosphatase: a protein tyrosine phosphatase involved in androgen-independent proliferation of prostate cancer. Endocrine-related cancer 12, 805–822 (2005).

50. Kantoff, P.W., et al. Sipuleucel-T immunotherapy for castration-resistant prostate cancer. N Engl J Med 363, 411–422 (2010).

51. Huben, R.P. Early detection of prostate cancer. Seminars in surgical oncology 5, 201–204 (1989).

52. El Ayachi, I., et al. The WNT10B Network Is Associated with Survival and Metastases in Chemoresistant Triple-Negative Breast Cancer. Cancer Res 79, 982–993 (2019).

53. Wend, P., et al. WNT10B/β-catenin signalling induces HMGA2 and proliferation in metastatic triple-negative breast cancer. EMBO Mol Med 5, 264–279 (2013).

54. Chan, J.J., et al. Pan-cancer pervasive upregulation of 3’ UTR splicing drives tumourigenesis. Nature cell biology 24, 928–939 (2022).

55. Hartman, M.L. & Czyz, M. BCL-G: 20 years of research on a non-typical protein from the BCL-2 family. Cell death and differentiation 30, 1437–1446 (2023).

56. Huang, X., et al. Wnt7a activates canonical Wnt signaling, promotes bladder cancer cell invasion, and is suppressed by miR-370-3p. J Biol Chem 293, 6693–6706 (2018).

57. MacLean, J.A., 2nd, King, M.L., Okuda, H. & Hayashi, K. WNT7A Regulation by miR-15b in Ovarian Cancer. PLoS One 11, e0156109 (2016).

58. Kondratov, A.G., et al. Alterations of the WNT7A gene in clear cell renal cell carcinomas. PLoS One 7, e47012 (2012).

59. Cui, P., et al. Pan-cancer analysis of the prognostic and immunological roles of SHP-1/ptpn6. Sci Rep 14, 23083 (2024).

60. Abdelatty, A., et al. Pan-Cancer Study on Protein Kinase C Family as a Potential Biomarker for the Tumors Immune Landscape and the Response to Immunotherapy. Frontiers in cell and developmental biology 9, 798319 (2021).

61. Xiong, Y., et al. WFDC2 suppresses prostate cancer metastasis by modulating EGFR signaling inactivation. Cell death & disease 11, 537 (2020).

62. Chen, Y., et al. WAP four-disulfide core domain protein 2 gene(WFDC2) is a target of estrogen in ovarian cancer cells. Journal of ovarian research 9, 10 (2016).

63. Min, B. & Wang, Y. WFDC2 is a potential prognostic and immunotherapy biomarker in lung adenocarcinoma. The Journal of international medical research 52, 3000605241258893 (2024).

64. Zhang, Y., Wang, S., Han, S. & Feng, Y. Pan-Cancer Analysis Based on EPOR Expression With Potential Value in Prognosis and Tumor Immunity in 33 Tumors. Frontiers in oncology 12, 844794 (2022).

65. Roberts, K.G., et al. Genetic alterations activating kinase and cytokine receptor signaling in high-risk acute lymphoblastic leukemia. Cancer cell 22, 153–166 (2012).

66. Iacobucci, I., et al. Truncating Erythropoietin Receptor Rearrangements in Acute Lymphoblastic Leukemia. Cancer cell 29, 186–200 (2016).

67. Nguyen, P.M., et al. Loss of Bcl-G, a Bcl-2 family member, augments the development of inflammation-associated colorectal cancer. Cell death and differentiation 27, 742–757 (2020).

68. Avgustinova, A., et al. Tumour cell-derived Wnt7a recruits and activates fibroblasts to promote tumour aggressiveness. Nat Commun 7, 10305 (2016).

69. Bai, J., et al. Overexpression of Cullin1 is associated with poor prognosis of patients with gastric cancer. Human pathology 42, 375–383 (2011).

70. Tan, D.X. & Hardeland, R. The Reserve/Maximum Capacity of Melatonin’s Synthetic Function for the Potential Dimorphism of Melatonin Production and Its Biological Significance in Mammals. *Molecules (Basel*, Switzerland) 26(2021).

71. Matous, J.G., et al. Shp-1 regulates the activity of low-affinity T cells specific to endogenous self-antigen during melanoma tumor growth and drives resistance to immune checkpoint inhibition. Journal for immunotherapy of cancer 13(2025).

72. Bingle, L., et al. WFDC2 (HE4): a potential role in the innate immunity of the oral cavity and respiratory tract and the development of adenocarcinomas of the lung. Respiratory research 7, 61 (2006).

73. Maldonado, M.D.M., Schlom, J. & Hamilton, D.H. Blockade of tumor-derived colony-stimulating factor 1 (CSF1) promotes an immune-permissive tumor microenvironment. Cancer immunology, immunotherapy : CII 72, 3349–3362 (2023).

74. Uhlén, M., et al. Proteomics. Tissue-based map of the human proteome. Science 347, 1260419 (2015).

75. Li, M., et al. Rediscovering publicly available single-cell data with the DISCO platform. Nucleic Acids Res 53, D932–d938 (2025).

76. Taupin, D. & Podolsky, D.K. Trefoil factors: initiators of mucosal healing. Nature reviews. Molecular cell biology 4, 721–732 (2003).

77. Suh, K.S., et al. Reciprocal modifications of CLIC4 in tumor epithelium and stroma mark malignant progression of multiple human cancers. Clin Cancer Res 13, 121–131 (2007).

78. Singha, B., et al. CLIC1 and CLIC4 complement CA125 as a diagnostic biomarker panel for all subtypes of epithelial ovarian cancer. Sci Rep 8, 14725 (2018).

79. Suryawanshi, S., et al. Complement pathway is frequently altered in endometriosis and endometriosis-associated ovarian cancer. Clin Cancer Res 20, 6163–6174 (2014).

80. Papadaki, V., et al. Two Secreted Proteoglycans, Activators of Urothelial Cell-Cell Adhesion, Negatively Contribute to Bladder Cancer Initiation and Progression. Cancers 12(2020).

81. Yang, Z., Yu, B., Hu, J., Jiang, L. & Jian, M. LRRC25 Is a Potential Biomarker for Predicting Immunotherapy Response in Patients with Gastric Cancer. Digestive diseases and sciences 70, 1395–1410 (2025).

82. Lang, Y., et al. IL-1R2 promotes tumorigenesis and modulates the tumor immune microenvironment in colorectal cancer. Cancer immunology, immunotherapy : CII 74, 284 (2025).

83. Li, J. & Xu, Z. NR3C2 suppresses the proliferation, migration, invasion and angiogenesis of colon cancer cells by inhibiting the AKT/ERK signaling pathway. Mol Med Rep 25(2022).

84. Liu, F. & Wu, H. Prognostic Value of Gastrokine-2 (GKN2) and Its Correlation with Tumor-Infiltrating Immune Cells in Lung Cancer and Gastric Cancers. Journal of inflammation research 13, 933–944 (2020).

85. Guy, M., et al. Identification of new genetic risk factors for prostate cancer. Asian journal of andrology 11, 49–55 (2009).

86. Chung Nien Chin, S., et al. Coordinate expression loss of GKN1 and GKN2 in gastric cancer via impairment of a glucocorticoid-responsive enhancer. American journal of physiology. Gastrointestinal and liver physiology 319, G175–g188 (2020).

87. Bergström, S.H., Järemo, H., Nilsson, M., Adamo, H.H. & Bergh, A. Prostate tumors downregulate microseminoprotein-beta (MSMB) in the surrounding benign prostate epithelium and this response is associated with tumor aggressiveness. Prostate 78, 257–265 (2018).

88. Yang, J., Fan, L., Liao, X., Cui, G. & Hu, H. CRTAC1 (Cartilage acidic protein 1) inhibits cell proliferation, migration, invasion and epithelial-mesenchymal transition (EMT) process in bladder cancer by downregulating Yin Yang 1 (YY1) to inactivate the TGF-β pathway. Bioengineered 12, 9377–9389 (2021).

89. Peake, B.F., Eze, S.M., Yang, L., Castellino, R.C. & Nahta, R. Growth differentiation factor 15 mediates epithelial mesenchymal transition and invasion of breast cancers through IGF-1R-FoxM1 signaling. Oncotarget 8, 94393–94406 (2017).

90. de Crevoisier, R., et al. Early PSA decrease is an independent predictive factor of clinical failure and specific survival in patients with localized prostate cancer treated by radiotherapy with or without androgen deprivation therapy. Annals of oncology : official journal of the European Society for Medical Oncology 21, 808–814 (2010).

91. Jiang, X., et al. Shared heritability and functional enrichment across six solid cancers. Nat Commun 10, 431 (2019).

92. Rashkin, S.R., et al. Pan-cancer study detects genetic risk variants and shared genetic basis in two large cohorts. Nat Commun 11, 4423 (2020).

93. Ishibashi, Y., et al. Serum TFF1 and TFF3 but not TFF2 are higher in women with breast cancer than in women without breast cancer. Sci Rep 7, 4846 (2017).

94. Thinyakul, C., et al. Hippo pathway in cancer cells induces NCAM1(+)αSMA(+) fibroblasts to modulate tumor microenvironment. Commun Biol 7, 1343 (2024).

95. Smith, J.P. & Solomon, T.E. Cholecystokinin and pancreatic cancer: the chicken or the egg? American journal of physiology. Gastrointestinal and liver physiology 306, G91–g101 (2014).

96. Jacobsen, F., et al. Cadherin-17 (CDH17) expression in human cancer: A tissue microarray study on 18,131 tumors. Pathology, research and practice 256, 155175 (2024).

97. Wang, D., et al. HSP90AB1 as the Druggable Target of Maggot Extract Reverses Cisplatin Resistance in Ovarian Cancer. Oxidative medicine and cellular longevity 2023, 9335440 (2023).

98. Chu, C.A., et al. The Role of Phosphatidylinositol 3-Kinase Catalytic Subunit Type 3 in the Pathogenesis of Human Cancer. International journal of molecular sciences 22(2021).

99. Sun, B.B., et al. Genomic atlas of the human plasma proteome. Nature 558, 73–79 (2018).

100. Lauwers, Y., et al. Imaging of tumor-associated macrophage dynamics during immunotherapy using a CD163-specific nanobody-based immunotracer. Proc Natl Acad Sci U S A 121, e2409668121 (2024).

101. Li, Y., Wu, Q., Lv, J. & Gu, J. A comprehensive pan-cancer analysis of CDH5 in immunological response. Front Immunol 14, 1239875 (2023).

102. Feng, P., et al. Golgi protein 73: the driver of inflammation in the immune and tumor microenvironment. Front Immunol 15, 1508034 (2024).

103. Guo, Q., et al. SERPIND1 Affects the Malignant Biological Behavior of Epithelial Ovarian Cancer via the PI3K/AKT Pathway: A Mechanistic Study. Frontiers in oncology 9, 954 (2019).

104. Inamoto, S., et al. Loss of SMAD4 Promotes Colorectal Cancer Progression by Accumulation of Myeloid-Derived Suppressor Cells through the CCL15-CCR1 Chemokine Axis. Clin Cancer Res 22, 492–501 (2016).

105. Bankier, S., et al. Circulating causal protein networks linked to future risk of myocardial infarction. medRxiv (2025).

106. Davey Smith, G. & Hemani, G. Mendelian randomization: genetic anchors for causal inference in epidemiological studies. Hum Mol Genet 23, R89–98 (2014).

107. Pierce, B.L. & Burgess, S. Efficient design for Mendelian randomization studies: subsample and 2-sample instrumental variable estimators. Am J Epidemiol 178, 1177–1184 (2013).

108. Jonmundsson, T., et al. Systematic comparison of observational and Mendelian Randomization estimates for cardiometabolic proteomic signatures. 2025.2012.2002.25341031 (2025).

109. Lee, M.A., et al. A proteogenomic analysis of the adiposity colorectal cancer relationship identifies GREM1 as a probable mediator. Int J Epidemiol 54(2024).

110. Peoples, A.R., et al. Genetic risk factors modulate the association between physical activity and colorectal cancer. Research square (2025).

111. Desai, T.A., et al. Identifying proteomic risk factors for overall, aggressive, and early onset prostate cancer using Mendelian Randomisation and tumour spatial transcriptomics. EBioMedicine 105, 105168 (2024).

112. Larsson, S.C., Chen, J., Ruan, X., Li, X. & Yuan, S. Genome-wide association study and Mendelian randomization analyses reveal insights into bladder cancer etiology. JNCI cancer spectrum 9(2025).

113. Schnakenberg, E., Breuer, R., Werdin, R., Dreikorn, K. & Schloot, W. Susceptibility genes: GSTM1 and GSTM3 as genetic risk factors in bladder cancer. Cytogenetics and cell genetics 91, 234–238 (2000).

114. Zhou, T., et al. Association of Glutathione S-transferase gene polymorphism with bladder Cancer susceptibility. BMC cancer 18, 1088 (2018).

115. Burger, M., et al. Epidemiology and risk factors of urothelial bladder cancer. European urology 63, 234–241 (2013).

116. Wang, Y., Yi, K., Chen, B., Zhang, B. & Jidong, G. Elucidating the susceptibility to breast cancer: an in-depth proteomic and transcriptomic investigation into novel potential plasma protein biomarkers. Frontiers in molecular biosciences 10, 1340917 (2023).

117. Tiwary, S., et al. ERBB3 is required for metastasis formation of melanoma cells. Oncogenesis 3, e110 (2014).

118. Buac, K., et al. NRG1 / ERBB3 signaling in melanocyte development and melanoma: inhibition of differentiation and promotion of proliferation. Pigment cell & melanoma research 22, 773–784 (2009).

119. Grzywa, T.M., et al. Myeloid Cell-Derived Arginase in Cancer Immune Response. Front Immunol 11, 938 (2020).

120. Arlauckas, S.P., et al. Arg1 expression defines immunosuppressive subsets of tumor-associated macrophages. Theranostics 8, 5842–5854 (2018).

121. Szklarczyk, D., et al. The STRING database in 2021: customizable protein-protein networks, and functional characterization of user-uploaded gene/measurement sets. Nucleic Acids Res 49, D605–d612 (2021).

122. Pal, S.K., Katheria, V. & Hurria, A. Evaluating the older patient with cancer: understanding frailty and the geriatric assessment. CA Cancer J Clin 60, 120–132 (2010).

123. Laconi, E., Marongiu, F. & DeGregori, J. Cancer as a disease of old age: changing mutational and microenvironmental landscapes. British journal of cancer 122, 943–952 (2020).

124. Lamb, J.R., et al. Predictive genes in adjacent normal tissue are preferentially altered by sCNV during tumorigenesis in liver cancer and may rate limiting. PLoS One 6, e20090 (2011).

125. Lamb, J.R., Jennings, L.L., Gudmundsdottir, V., Gudnason, V. & Emilsson, V. It’s in our blood: a glimpse of personalized medicine. Trends in molecular medicine 27, 20–30 (2021).

126. Tan, A.C., et al. Detection of circulating tumor DNA with ultradeep sequencing of plasma cell-free DNA for monitoring minimal residual disease and early detection of recurrence in early-stage lung cancer. Cancer 130, 1758–1765 (2024).

127. Sigurdsson, E., Thorgeirsson, G., Sigvaldason, H. & Sigfusson, N. Unrecognized myocardial infarction: epidemiology, clinical characteristics, and the prognostic role of angina pectoris. The Reykjavik Study. Ann Intern Med 122, 96–102 (1995).

128. Merkler, A.E., et al. Association Between Unrecognized Myocardial Infarction and Cerebral Infarction on Magnetic Resonance Imaging. JAMA Neurol 76, 956–961 (2019).

129. Psaty, B.M., et al. Cohorts for Heart and Aging Research in Genomic Epidemiology (CHARGE) Consortium: Design of prospective meta-analyses of genome-wide association studies from 5 cohorts. Circ Cardiovasc Genet 2, 73–80 (2009).

130. Igap & Schellenberg, G.D. International Genomics of Alzheimer’s Disease Project (IGAP) genome-wide association study. Alzheimer’s & Dementia: The Journal of the Alzheimer’s Association 8, P101 (2012).

131. Emerging Risk Factors, C., et al. The Emerging Risk Factors Collaboration: analysis of individual data on lipid, inflammatory and other markers in over 1.1 million participants in 104 prospective studies of cardiovascular diseases. Eur J Epidemiol 22, 839–869 (2007).

132. Inker, L.A., et al. Estimating glomerular filtration rate from serum creatinine and cystatin C. N Engl J Med 367, 20–29 (2012).

133. Cespedes Feliciano, E.M., et al. Association of Prediagnostic Frailty, Change in Frailty Status, and Mortality After Cancer Diagnosis in the Women’s Health Initiative. JAMA network open 3, e2016747 (2020).

134. Corey, K.E., et al. ADAMTSL2 protein and a soluble biomarker signature identify at-risk non-alcoholic steatohepatitis and fibrosis in adults with NAFLD. Journal of hepatology 76, 25–33 (2022).

135. Taliun, D., et al. Sequencing of 53,831 diverse genomes from the NHLBI TOPMed Program. Nature 590, 290–299 (2021).

136. Lachmann, A., et al. Geneshot: search engine for ranking genes from arbitrary text queries. Nucleic Acids Res 47, W571–w577 (2019).

137. Gkatzionis, A., Burgess, S. & Newcombe, P.J. Statistical methods for cis-Mendelian randomization with two-sample summary-level data. Genet Epidemiol 47, 3–25 (2023).

