## Supplementary Material for "Mechanistic Insights into Cancer Risk from the Circulating Proteome"

[1]Icelandic Heart Association, Holtasmari 1, IS-201 Kopavogur, Iceland.
[2]Faculty of Medicine, University of Iceland, 101 Reykjavik, Iceland.
[3]Computational Biology Unit, Department of Informatics, University of Bergen, 5020 Bergen, Norway
[4]Faculty of Pharmaceutical Sciences, University of Iceland, Reykjavik, Iceland
[5]Novartis Biomedical Research, 22 Windsor Street, Cambridge, MA 02139, USA.
[6]Department of Genetics and Molecular Medicine, University Hospital, Reykjavík, Iceland
[7]Laboratory of Epidemiology and Population Sciences, National Institute on Aging, MD, USA
[8]Cancer Epidemiology, University of Zurich, Epidemiology, Biostatistics and Prevention Institute, CH-8001 Zurich, Switzerland
[9]Department of Medical Oncology, Landspitali University Hospital, Reykjavik, Iceland
[10]Biomedical Center, School of Health Sciences, University of Iceland, Reykjavík, Iceland
[11]Biomedical Center, School of Health Sciences, University of Iceland and Department of Hematology, University Hospital, Reykjavík, Iceland
[12]Novartis Biomedical Research, 10675 John Jay Hopkins Drive, San Diego, CA 92121, USA
[13]Monoceros Biosystems, 12636 High Bluff Drive, Suite 400, San Diego, CA. 92130, USA

†Shared first authors

### Supplementary Note 1

#### *Overview of the research approach*

This study systematically assesses associations between 7,523 serum proteins and 13 distinct cancer types, grouped by shared body system, in the population-based Age, Gene/Environment Susceptibility Reykjavik Study (AGES) cohort of older adults[1]. Cancer types span the digestive, genitourinary, respiratory, female reproductive systems, and skin. Associations with "any cancer" are also analyzed. Cancer type abbreviations follow main-text Fig. 1.

Incident cancers (diagnosed post-baseline) were modeled with Cox proportional hazards[2], using $log_2$-transformed proteomics; prevalent cancers (pre-baseline diagnoses) used logistic regression. Outliers (exceeding the 99.5th percentile) were excluded. Given that declining kidney function may bias serum protein levels in the elderly, all analyses adjust for age, sex, and estimated glomerular filtration rate (eGFR), referred to as "standard covariates" from now on. Additional corrections include body mass index (BMI, kg/m$^2$), tobacco smoking (categorized as never, former, or current smoker), and alcohol consumption (measured in units per week), tailored to each cancer type. Specific variables like body height for prostate and blood pressure for kidney cancers[3,4], were also considered. These adjustments were made to improve estimate precision[5], and may identify circulating proteins and biological processes that are independent of these recognized risk variables. To account for multiple hypothesis testing, the false discovery rate (FDR) was adjusted using the Benjamini-Hochberg method[6], and we used an arbitrary minimum of 10 cases per study.

In this study, follow-up for incident cancer cases lasted up to 13.6 years from baseline, with person-years calculated from the participant’s first (baseline) visit until cancer diagnosis, death, or end of follow-up. Prevalent cancer cases at the baseline visit refer to patients who had already been diagnosed with cancer before enrollment in the study. All proteins associated with prevalent or incident cancer types with an FDR adjusted P-value < 0.05 are considered significant, but we additionally report on a more lenient FDR adjusted P < 0.10 to allow for maximum discovery across the sample. Table S1 details the number of patients for each of the 13 cancer types examined in the present study. Tables S2-S7 present descriptive statistics, with no missing values, stratified by all 13 cancer cases combined or different cancer types combined if originating from the same body system. Excel Table S8 lists the ICD-10 diagnosis codes for additional cancer types beyond the 13 specifically analyzed in this study, while Table S9 summarizes descriptive statistics for the 684 participants (of 1,916 total cancer cases) diagnosed with these cancers. In total, 772 diagnoses were recorded, as some individuals were diagnosed with more than one cancer type (Table S8).

Associations between 7,523 serum proteins and multiple cancer types from Cox and logistic regression analyses are provided in Tables S10 and S11 (excel files), respectively; see also Supplementary Notes 2–7 below. Table S12 (excel file) summarizes the key characteristics of proteins significantly associated (FDR < 0.05) with cancer in the observational analysis. The results of the observational analyses for the 1,916 individuals diagnosed with any type of incident or prevalent cancer are provided in Table S13 and Table S14 (excel files), respectively; see also related Supplementary Note 8 below. We acknowledge that both the aptamer and the corresponding protein annotation are included in data tables; however, most of the discussion centers on the unique gene symbol for proteins identified by a single or multiple aptamers.

Several characteristics were analyzed for serum proteins significantly associated with cancers in the observational analyses (FDR < 0.05) listed in Table S12. These included for instance the identification of proximal (*cis*-acting) protein quantitative trait loci (pQTLs) in the AGES study, differential regulation at the corresponding tumor site or related body system using Gene Expression Profiling Interactive Analysis (GEPIA)[7], a web-based tool that provides interactive and customizable functionalities to analyze tumor-specific RNA sequencing expression data from The

Cancer Genome Atlas (TCGA) (https://www.cancer.gov/tcga) and Genotype-Tissue Expression (GTEx)[8] databases. We also included genetic associations from the GWAS Catalog[9] and the FinnGen study[10], as well as *cis*-acting expression and splice QTLs (eQTL and sQTL, respectively) from the GTEx portal. Only expression or splice SNPs reported in the GTEx portal that directly match the *cis*-acting protein SNP in Table S12 were considered, i.e., proxy SNPs at a certain correlation threshold were not included. Additionally, since some of the protein SNPs we report are relatively rare, they may be absent from the GTEx portal. We note in passing that significant findings from the GWAS and FinnGen databases do not necessarily imply that the protein-coding gene is causal for the phenotype in question; rather, they highlight the GWAS-associated SNP nearest to the protein encoding gene. To evaluate protein panel enrichment across tissues, we utilized the Human Protein Atlas (HPA) portal[11].

Finally, Supplementary Notes 9–11 (below) presents a series of crosscutting, integrative analyses that extend the originality of our findings by linking the circulating proteome to both external and internal datasets. These analyses span germline genetic susceptibility for specific cancers and pan-cancer genetic risk (Supplementary Note 9), the incorporation of cancer-associated proteins into biological networks reconstructed within the AGES cohort (Supplementary Note 10), and a causal inference framework based on forward two-sample Mendelian randomization (MR) using proteome-wide 2,062 *cis*-acting pQTLs identified in AGES (Supplementary Note 11).

**Reference tables and figures**:

- Table S1: Case counts by cancer type.
- Tables S2–S7: Clinical characteristics of cancer types grouped by body system.
- Table S8 (excel)–S9: Coverage for other (non-primary) cancer diagnoses.

- Fig. 1 (main text): Summary of the number of cancer patients and cancer types included in this study
- Tables S10 (excel) – S11 (excel): Complete association results for each protein at FDR thresholds of 0.05 and 0.10, for incident and prevalent cancers, respectively.
- Table S12 (excel): Summary characteristics of proteins with significant associations (FDR < 0.05) in the observational analysis.
- Figs. S1-S5: Volcano plots showing serum proteins associated with different types of cancer, organized by the shared body system of respective cancer diagnosis.
- Figs. S6-S7: The effect of covariate adjustment on proteins associated with incident and/or prevalent cancers, respectively.
- Fig. S8: Tissue specificity of serum proteins associated with prevalent stomach cancer.
- Tables S13–S14 (excel): Associations with any cancer for incident and prevalent diagnoses, respectively.

### Supplementary Note 2

#### *Digestive system cancers*

Cancers of the digestive system are among the most common cancers and the primary cause of cancer deaths globally[12], accounting for 26% of all new cancer diagnosis and 35% of all cancer-related deaths. In the current study, cancers of the digestive system comprised esophageal cancer, stomach cancer, colon cancer, rectal cancer, and pancreatic cancer. Table S1 shows the number of individuals diagnosed with either prevalent or incident cancers of the digestive system in the AGES cohort, while Table S3 provides descriptive statistics stratified by all cancers of the digestive system combined. Figure S1 presents a multi-cancer volcano plot of serum proteins associated with incident or prevalent cancers of the digestive system, with additional details on the findings provided below

##### Esophageal cancer (ESC)

The cancer of the esophagus (ESC, ICD-10 code C15) is an aggressive gastrointestinal cancer that is usually diagnosed at an advanced stage, with a 5-year survival rate of only 15-25% once diagnosed[13]. It is noteworthy that the incidence of ESC is higher in men than in women[14]. In the current study, 16 individuals were diagnosed with incident ESC, three of whom were women. Additionally, only three participants, including two women, had been diagnosed with ESC before the baseline visit. We identified 13 serum proteins associated with all incident ESC cases (FDR $< 0.05$) after adjusting for the standard covariates: age, sex, and eGFR (Table S10). Obesity, smoking, and alcohol intake are the most well-known non-genetic risk factors for ESC[13]. When the three females were excluded from the analysis, i.e., males only analysis, three additional proteins were found to be linked with incident ESC at FDR $< 0.05$, i.e. nuclear receptor subfamily 5 group A member 2 (NR5A2), signal sequence receptor subunit 2 (SSR2) and transmembrane serine protease 11A (TMPRSS11A) (Table S10). After adjusting for BMI, smoking, and alcohol use in addition to standard covariates, six serum proteins remained associated with incident ESC at FDR < 0.05 (Table S10). Thus, at FDR < 0.05, a total of 16 proteins were associated with new-onset ESC whereas at FDR < 0.10 a total of 30 proteins were associated with ESC (Table S10).

***Key observations and functional highlights**:* The different characteristics of each serum protein that were associated with incident ESC at FDR < 0.05 are summarized in Table S12. Examples of proteins associated with all incident ESC cases include phospholipase A and acyltransferase 4 (PLAAT4), transcription factor MafG (MAFG) and WW domain containing oxidoreductase (WWOX) (Table S10). For instance, the retinoic acid early transcript 1L protein (RAET1L), which was associated with incident ESC (Table S10), is expressed exclusively in the esophagus and its cognate mRNA is expressed differentially in esophageal tumors (Table S12). Furthermore, RAET1L shows strong *cis*-acting regulatory control in serum (Table S12), and a corresponding *cis*-eQTL for RAET1L expression in esophageal tissue is reported in the GTEx portal (Table S12). Another notable protein associated with incident ESC is WWOX, whose encoding gene contains a so-called fragile site[15], characterized by high mutation rates in cancer cells, likely due to rearrangements resulting from defects in the DNA repair process. Some of the 16 proteins associated with incident ESC (FDR < 0.05) were specifically expressed in the esophagus or other parts of the digestive system (Table S12).

#### Stomach cancer (STC)

Stomach cancer (STC, aka gastric cancer, ICD-10 code C16) is one of the leading causes of cancer-related deaths worldwide[16], and since it is often diagnosed at an advanced stage the prognosis is generally poor. Tobacco use, obesity, alcohol consumption, and infection with Helicobacter pylori (H pylori) are all well-known risk factors for STC[17]. In the AGES cohort, 51 individuals were diagnosed with STC: 25 participants (12 females) had the diagnosis before the baseline blood collection, while 26 participants (11 females) developed the disease during the follow-up period. At FDR < 0.05, and controlling for standard covariates, no protein was associated with all cases of incident STC, whereas two proteins, cullin 1 (CUL1) and leukocyte cell derived chemotaxin 2 (LECT2), were significantly associated with incident STC in a sex-specific manner (Table S10). After controlling for risk variables such as BMI, smoking status, and alcohol intake, two additional proteins, TNFSF4 and SPATA5, were found to be associated with incident STC in a sex-specific manner at FDR < 0.05 (Table S10). Overall, four serum proteins were associated with incident STC at FDR < 0.05, and six serum proteins at FDR < 0.10 (Table S10). In terms of prevalent STC, we found that 30 serum proteins were linked to all prevalent STC at FDR < 0.05 after adjusting for standard covariates (Table S11). Using additional available covariate data (data on H pylori infections not available) including BMI, smoking status, and alcohol consumption, 14 proteins remained associated with prevalent STC at FDR < 0.05 (Table S11). Additionally, three more proteins were found to be associated with prevalent STC in males only at an FDR < 0.05 (Table S11), but these associations were no longer significant after adjusting for all covariates. Overall, 33 serum proteins were significantly associated with prevalent STC at FDR < 0.05 (Table S11), while a total of 62 proteins were at FDR < 0.10 (Table S11).

***Notable findings and functional highlights****:* Serum proteins associated with prevalent STC included for example the trefoil factors 1-3 (TFF1-3) and the anti-inflammatory protein gastrokine 2 (GKN2), a previously proposed prognostic biomarker for STC[18]. Additionally, the regenerating islet-derived (Reg) proteins REG1A and REG1B, which are specifically expressed in the pancreas, were positively associated with the risk of prevalent STC (Table S11). Both proteins have also been previously linked to gastric cancer[19]. Furthermore, three serum proteins, MANSC domain containing 4 (MANSC4), cytokine receptor like factor 3 (CRLF3), and phosphotyrosine interaction domain containing 1 (PID1), were associated with prevalent STC in males alone (Table S11). Serum levels of TFF1, TFF2 and GKN2 were found to be inversely related to prevalent STC, whereas TFF3 was directly related to prevalent STC (Table S11). The relationship between the gene expression levels of the genes encoding these proteins and STC in solid tissues has previously been reported and is consistent with the current findings[20-22]. Notably, however, proteins inversely associated with prevalent STC, including TFF1, GKN2, cobalamin-binding intrinsic factor (CBLIF), and TFF2, are all specifically expressed in the stomach (Table S12). Although these associations may reflect genuine disease-related mechanisms (e.g., genetic and rodent studies reporting an inverse association of GKN2 with STC; see Supplementary Note 9 below), the observed inverse relationship with STC diagnosed prior to protein measurements may partly reflect patients in the cohort who had undergone partial or total gastrectomy.

Many of the genes encoding serum proteins associated with prevalent STC are expressed exclusively in digestive system regions and differentially expressed in gastric tumors (Table S12, Fig. S8). For example, GKN2 is specifically expressed in stomach and negatively associated with prevalent STC and downregulated in stomach tumors (Table S12). Furthermore, several of these proteins, such as GKN2, are regulated in a *cis*-acting manner (Table S12). Furthermore, the genetic variant rs62133344 regulates GKN2 in *cis* and TFF1 in *trans* (Table S12, and more details in Supplementary Note 9), another protein linked to prevalent STC in our study (Table S11). Interestingly, studies have shown that GKN2 and TFF1 are co-regulated and form heterodimers in mucus secreting cells of the stomach[23], hence affecting mucous homeostasis. Importantly, this co-regulatory behavior of GKN2 and TFF1 is frequently lacking in stomach cancers[24]. Finally, GKN2 and TFF1 were recently found to synergistically mediate antiproliferative effects on stomach cancer[25]. The protein TNFSF4 (aka OX40L), associated with incident STC after adjusting for standard covariates plus well-established STC risk factors (Table S10), serves yet another example. Immunotherapy can elevate OX40L levels, which function as a stimulatory checkpoint on dendritic cells while simultaneously reducing the expression of the inhibitory checkpoint PD-L1 on tumor cells[26]. This dual effect enhances the effectiveness of antitumor immunotherapy in gastrointestinal cancers[26]. Conversely, TNFSF4 was associated with an increased risk of STC in our study (Table S10), potentially indicating elevated TNFSF4 levels as a response to the advancing prodromal stage of STC.

#### Colon and rectal cancers (COC, REC)

Colon cancer (COC, ICD-10 code C18) and rectal cancer (REC, ICD-10 code C20) are frequently referred to as colorectal cancer (CRC) and are often studied as a single disease[27]. Both colon and rectal cancers occur in the large intestine, which is the lowest part of the digestive system. Despite their anatomical proximity, there are significant pathophysiological differences between them. For example, while physical activity reduces the risk of colon cancer (COC), it does not affect the risk of developing rectal cancer (REC)[28]. In fact, it has been suggested that these two types of cancer should be studied separately[28]. In this study, we analyzed colon and rectal cancers separately. Colon cancer is the world's second leading cause of death among all cancers and the third most common form of cancer[29]. In addition to a strong genetic component underlying COC[30], diet (e.g. red meat), tobacco use, alcohol intake, and obesity are frequent modifiable lifestyle factors that increase the risk of COC[31]. Overall, 183 individuals were diagnosed with COC in the AGES (Table S1). Prior to the protein measurements in AGES, 55 individuals (28 females) had been diagnosed with COC, while 128 individuals (75 females) developed the disease during the follow-up period. No proteins were significantly associated with prevalent COC, whereas eight proteins were associated with incident COC at FDR < 0.05, six of which were sex-specific, using the standard covariate adjustment (Table S10). These include for instance the serum proteins ASH1 like histone lysine methyltransferase (ASH1L) and high density lipoprotein binding protein (HDLBP) in females and BCL2 like 14 (BCL2L14) in males (Table S10). An additional protein, PIH1 domain containing 2 (PIH1D2), was identified exclusively in females after further adjustments for BMI, smoking status, and alcohol consumption (Table S10). In summary, nine proteins were associated with incident COC at an FDR < 0.05, and 15 at an FDR < 0.10 (Table S10).

Rectal cancer is less prevalent than colon cancer, yet it is still one of the most common malignancies worldwide[29]. In the current study, 31 individuals were diagnosed with REC: at the baseline visit, 11 participants (3 females) had a prior diagnosis of REC, while 20 individuals (8 females) were diagnosed during the follow-up, after the protein measurements. Using standard covariate adjustment, three proteins were significantly (FDR < 0.05) linked to all incident REC (Table S10), which remained significantly associated with REC following further adjustment for BMI, smoking status, and alcohol intake (Table S10). Additional 10 serum proteins were associated with incident REC in males only at FDR < 0.05 including for instance defensin alpha 1, defensin alpha 1B (DEFA1), RAB22A member of RAS oncogene family (RAB22A) and thioesterase superfamily member 4 (THEM4) (Table S10). In summary, there were 13 proteins linked to incident REC at FDR < 0.05, with overall 17 proteins associated with REC at FDR < 0.10 (Table S11). There were 17 proteins associated with prevalent REC at FDR < 0.05 adjusting for the standard covariates (Table S11), but with further adjustments nine proteins remained associated with REC (Table S11).

***Functional highlights***: Serum proteins significantly linked to incident COC included for instance C-X-C motif chemokine ligand 8 (CXCL8), myosin light chain 6 (MYL6) and dual specificity phosphatase 16 (DUSP16), all previously linked to cancer development or treatment[32-34]. For instance, elevated expression of the chemokine CXCL8 in the tumor microenvironment (TME) is associated with advanced stage, right-sided tumors and poor survival in colorectal cancer[35]. Moreover, studies have demonstrated that CXCL8 plays a key role in the onset and progression of COC by promoting tumor growth, facilitating immune evasion, and modulating a stem cell-like phenotype in cancer cells[36,37]. Proteins associated with incident REC included the proteins heterogeneous nuclear ribonucleoprotein A1 (HNRNPA1), interleukin 17F (IL17F), and Wnt family member 7A (WNT7A) (Table S10). These three proteins have been linked to or proposed to play significant roles in the development of CRC. More to the point, HNRNPA1 is involved in RNA processing and splicing, which may affect cancer cell proliferation and survival[38]. IL17F contributes to inflammation, which promotes tumorigenesis[39], while WNT7A, a tumor suppressor, is part of the Wnt signaling pathway, known for its critical role in cell proliferation and cancer progression[40]. Various proteins associated with COC or REC are differently regulated in the relevant tumor (Table S12).

#### Pancreatic cancer (PAC)

Pancreatic cancer (PAC, ICD-10 code C25) has one of the worst cancer prognoses, and despite being less common than many other cancers, it is expected to become the second leading cause of cancer deaths by 2030 in the US[41]. Only one patient was diagnosed with PAC at the baseline visit, whereas 38 subjects (25 females) developed the disease during the follow-up period. As with many other cancers, tobacco use, obesity, and alcohol consumption are the most common modifiable risk factors[41]. Four proteins, including for instance CD300 molecule like family member g (CD300LG) and protein tyrosine phosphatase non-receptor type 6 (PTPN6), were linked to incident PAC in a sex-specific manner at FDR < 0.05 using standard covariate adjustment (Table S10). Additionally, two proteins, RNA polymerase I and III subunit C (POLR1C) and carbohydrate sulfotransferase 12 (CHST12), were linked to incident PAC at FDR < 0.05 with further adjustment including BMI, smoking status, and alcohol intake, likewise in a sex-specific manner (Table S10). In total, six proteins were linked to an incident PAC at FDR < 0.05, while nine proteins were associated at an FDR < 0.10 (see Table S10).

***Notable findings***: Different characteristics of the serum proteins associated with incident PAC at FDR < 0.05 are highlighted in Table S12. In this study, three of the six proteins associated with incident PAC, i.e., sialic acid binding Ig like lectin 6 (SIGLEC6), CD300LG and CHST12 were strongly linked to *cis*-acting pQTLs. Three proteins associated with incident PAC, i.e., PTPN6, POLR1C, and CHST12, are upregulated in pancreatic tumors (Table S12). According to a recent study, PTPN6 predicts pancreatic cancer well in a panel with three other immune-related genes[42].

Finally, SIGLEC6, a member of the sialic acid-binding immunoglobulin-like lectin family, may play a role in modulating tumor cell phenotypes by interacting with sialoglycans on cancer cells[43]. These interactions could contribute to immune suppression within the TME, aiding tumor evasion of immune surveillance[43].

**Reference tables and figures**:

- Tables S10–S11 (excel): Complete association results for each protein at FDR thresholds of 0.05 and 0.10, for incident and prevalent cancers, respectively.
- Table S12 (excel): Summary characteristics of proteins with significant associations (FDR < 0.05).
- Fig. S1: A multi-cancer volcano plot summarizes the associations between serum proteins and both incident and prevalent digestive system cancers examined in this study, with selected protein annotations highlighted
- Fig. S8: Tissue specificity of serum proteins associated with prevalent stomach cancer.

### Supplementary Note 3

#### *Genitourinary system cancers*

Cancers of the genitourinary system, those originating in any of the genitourinary organs, are the most common tumors affecting men[44]. In the current study, cancers of the genitourinary system comprise kidney cancer, prostate cancer, and bladder cancer. Table S1 shows the number of individuals diagnosed with either prevalent or incident cancers of the genitourinary system in the AGES cohort, while Table S4 provides descriptive statistics stratified by all cancers of the genitourinary system combined. Figure S2 presents a multi-cancer volcano plot of serum proteins associated with incident or prevalent genitourinary cancers, with additional details on the findings provided below.

##### **Kidney cancer (except renal pelvis) (KIC)**

Kidney cancer (KIC, ICD-10 code C64), is the 9$^{th}$ most common cancer in men and the 14$^{th}$ most common cancer in women worldwide[45]. Men are more likely than women to be diagnosed with KIC, and they have a more aggressive histology, larger tumors, and poorer oncological prognosis[46]. Indeed, of the 81 individuals diagnosed with KIC in the AGES study, 52 were males: 32 individuals (11 females) were diagnosed with KIC prior to the first visit, while 49 individuals (18 females) developed KIC after the first visit, or approximately three times more males than females. High blood pressure, obesity, and tobacco use are all common risk factors for KIC[4]. Four proteins were associated with incident KIC at FDR < 0.05 after adjusting for the standard covariates (including eGFR): kidney injury molecule-1 (KIM-1, aka HAVCR1), plexin B3 (PLXNB3), small nuclear ribonucleoprotein polypeptide C (SNRPC), and GTPase, IMAP family member 4 (GIMAP4) (Table S10). When correcting for additional covariates like BMI, smoking status, and systolic and diastolic blood pressure, and analyzing the sexes separately, these proteins remained associated with all incident KIC, with GIMAP4 in males alone, and an additional protein,

GDP-L-fucose synthase (GFUS, aka TSTA3), in females only (Table S10). Overall, five serum proteins were associated with incident KIC at FDR < 0.05, and 23 proteins at FDR < 0.10 (Table S10). Three proteins, protein phosphatase 1 regulatory subunit 10 (PPP1R10), EPH receptor B2 (EPHB2), and gastrin releasing peptide (GRP), were found to be associated with prevalent KIC at FDR < 0.05 (Table S11). These associations remained significant after adjusting for BMI, blood pressure and smoking status (Table S11). Two other proteins, butyrophilin 8 (BTNL8) and RNA transcription, translation, and transport factor (RTRAF), were found to be associated with prevalent KIC in females alone (Table S11). Overall, five proteins were significantly associated with prevalent KIC at FDR < 0.05, with no additional proteins at FDR < 0.10.

***Notable findings and functional highlights**:* Different characteristics of the serum proteins associated with prevalent or incident KIC at FDR < 0.05 are listed in Table S12. For example, the protein HAVCR1 (as well as GIMAP4), which is directly related to incident KIC (Table S10), is upregulated in renal tumors according to The Cancer Genome Atlas Kidney samples (TCGA-KIRC)[7] (Table S12). Intriguingly, HAVCR1 has previously been found to be over-expressed in urine of patients suffering from KIC[47], which is directionally consistent with our findings. In line with these findings, we find that increased HAVCR1 serum levels are associated with shorter overall survival (all-cause mortality) in the AGES ($P = 2 \times 10^{-10}$). Interestingly, and somewhat counterintuitive to the data from TCGA-KIRC[7], low levels of HAVCR1 and GIMAP4 in kidney tumors are predictors of poor prognosis in TCGA-KIRC[7], highlighting potential differences between tissue/tumor-specific and circulating biology. HAVCR1, a member of the T-cell immunoglobulin and mucin domain gene family, is, according to the Human Protein Atlas[48], expressed specifically in kidney and intestine. The EPHB2 protein, which is directly associated to prevalent KIC in our study and under *cis*-acting control (Tables S11-S12), has been linked to kidney injury in mice and is over-expressed and activated in fibrotic kidney tissues[49]. EPHB2, however, does not exhibit differential expression in kidney tumors nor has an impact on survival, according to GEPIA[7]. HAVCR1, GIMAP4, and EPHB2, are all subject to *cis*-acting genetic control in the AGES (Table S12).

A decline in eGFR (one of the standard covariates used in this study; see Supplementary Note 1) is associated with renal dysfunction and is expected to be present in individuals with a history of KIC prior to study enrollment. When adjusting for age and sex alone, 624 serum proteins were associated with prevalent KIC at an FDR < 0.05, and these proteins were also independently linked to eGFR (data not shown). When eGFR was included as a covariate, three proteins were significantly associated with prevalent KIC, while five proteins showed significant associations with prevalent KIC after full covariate adjustment (see above). When eGFR was excluded from the adjustment for incident KIC, the effect differed from that observed for prevalent KIC, with three proteins remaining associated including HAVCR1, GIMAP4, and GFUS, which were also among the four proteins linked to incident KIC when eGFR was included in the model. This suggests that renal dysfunction was likely present in KIC patients at the baseline visit but had not yet manifested during the prodromal phase of the disease.

**<u>Prostate cancer (PRC)</u>**

Prostate cancer (PRC, ICD-10 code C61) is the most common type of cancer in men and the fifth leading cause of death from cancer worldwide[50]. Many risk factors have been linked to PRC, but the evidence varies due to the disease's heterogeneity, with obesity, tobacco smoking, and height all potential risk factors[3]. In this study, 299 males were diagnosed with PRC: 107 males were identified with PRC at the baseline visit, and 192 males developed PRC throughout the follow-up period. The Cox regression analysis identified five serum proteins associated with incident PRC including the well-established biomarker prostate-specific antigen (PSA, aka KLK3)[51] and prostatic acid phosphatase PAP (aka ACP3) (Table S10). Using additional covariates including height, BMI and smoking status, all, including KLK3 and ACP3, remained significantly associated with incident PRC (Table S10), but an additional protein, apolipoprotein M (APOM), appeared

(Table S10). In summary six proteins were linked to incident PRC at FDR < 0.05, while 9 were at FDR <0.10 (Table S10). Seven proteins were associated with prevalent PRC at FDR < 0.05, including KLK3 and microseminoprotein beta (MSMB) (Table S11), but variants across the *MSMB* gene have been linked to genetic risk of PRC[52]. Adding BMI, height and smoking status as additional covariates in the regression analyses did not affect the outcome compared to using the standard covariates only (Table S11). Thus seven proteins were associated with prevalent KIC at FDR < 0.05, while 13 proteins were at FDR < 0.10 (Table S11).

***Functional highlights***: Table S12 delineates the characteristics of serum proteins significantly (FDR < 0.05) associated with prevalent and/or incident PRC. Notably, KLK3 was associated with both prevalent and incident PRC, but in opposite directions (Tables S10-S11). The inverse relationship between KLK3 and prevalent PRC may indicate that some patients with prevalent PRC had low baseline KLK3 serum levels because of prostatectomy. Another notable finding is the serum protein MSMB, which is associated with prevalent PRC in the current study and with malignant neoplasm of the prostate in both the FinnGen and GWAS Catalog databases (Table S12). Intriguingly, in a MR analysis[53], men with higher plasma levels of MSMB were causally linked to a reduced risk of PRC, supporting the inverse correlation between MSMB and PRC as observed in our observational analysis (Table S11); see also the causal inference analysis in Supplementary Note 11 below. Consistent with this finding, the minor allele of the causal variant rs10993994, which increases the risk of PRC[53], is also associated with decreased MSMB levels in serum (Table S12). However, since MSMB is predominantly expressed in the prostate gland, the lower serum levels of the protein observed in PRC patients may partially result from some patients having undergone prostatectomies.

**<u>Bladder cancer (BLC)</u>**

Urothelial bladder cancer (BLC, ICD-10 code C67) is most common in developed countries and is now the 9th most common type of cancer worldwide[54], with males being more frequently affected than females[54]. In fact, males outnumbered females by more than three to one among BLC patients in this study, with 71 (21 females) having prevalent BLC and 77 (17 females) diagnose with incident BLC. Tobacco smoking is the single most frequent risk factor for BLC[55]. Six proteins were significantly associated with all incident BLC cases at FDR < 0.05 using standard covariates, with 19 additional proteins associated with incident BLC in a sex-specific manner at the same FDR threshold (Table S10). Four additional proteins showed associations with incident BLC after adjustment for smoking status (Table S10), including kallikrein-1 (KLK1), a member of the same gene family as KLK3 (aka PSA). Overall, 29 proteins were linked to incident BLC at FDR < 0.05 and 82 proteins at FDR < 0.10 (Table S10). At FDR < 0.05, three proteins were associated with prevalent BLC, with an additional two proteins in females alone including nuclear receptor subfamily 3 group C member 2 (NR3C2) and interleukin 1 receptor type 2 (IL1R2) (Table S11). Adjusting for smoking status had no effect on the regression analysis results for prevalent BLC (Table S11).

***Highlights***: Table S12 provides an overview of the serum protein characteristics associated with prevalent or incident BLC at an FDR < 0.05. Most of the proteins associated with BLC do not have a well-established role in cancer. However, several show differential expression in BLC and/or other genitourinary cancers (Table S12). Interestingly, the gene encoding KLK1 associated with incident BLC, is located proximal to variants linked to PRC and PSA levels in different GWAS studies (Table S12). The gene encoding NR3C2 (Table S11), which encodes the mineralocorticoid receptor, functions as a potent tumor suppressor in urothelial/bladder carcinoma[56]. In muscle-invasive bladder cancer, NR3C2 expression is markedly lower than in adjacent normal urothelium, and high expression independently predicts improved cancer-specific survival[56]. Moreover, NR3C2 overexpression inhibits angiogenesis and suppresses AKT/ERK signaling in colon cancer cells[57]. While NR3C2 is established as a tumor suppressor in bladder carcinogenesis, our observation of a positive association in prevalent BLC cases (Table S11), suggests that its role may shift in established tumors, potentially contributing to disease resistance. The other protein associated with BLC in a sex-specific manner was IL1R2, a decoy receptor previously shown to promote progression and metastasis in clear cell renal carcinoma[58]. IL1R2 primarily functions by sequestering pro-inflammatory IL-1 ligands, such as IL-1β, thereby dampening anti-tumor immune responses and fostering a tumor-promoting microenvironment[59]. Accordingly, the positive association of IL1R2 with prevalent bladder cancer observed in our study (Table S11) aligns with these prior findings.

**Reference tables and figures**:

- Tables S10 (excel) – S11 (excel): Complete association results for each protein at FDR thresholds of 0.05 and 0.10, for incident and prevalent cancers, respectively.
- Table S12 (excel): Summary characteristics of proteins with significant associations (FDR < 0.05).
- Fig. S2: A multi-cancer volcano plot summarizes the associations between serum proteins and both incident and prevalent genitourinary cancers analyzed in this study, with selected proteins highlighted for illustration.

### Supplementary Note 4

#### *Respiratory system cancer*

The most common cancers of the respiratory system occur in the trachea, lungs, bronchus, and larynx, with lung cancer being the most prevalent[60]. These cancers represent a significant public health burden, with smoking and environmental exposure being the most common risk factors[60]. In the present study, we report on the association of 7,523 serum proteins with cancers of the lungs and bronchus (hereafter lung cancer). Table S1 displays the number of individuals diagnosed with either prevalent or incident lung cancer in the current study, while Table S5 provides descriptive statistics stratified by combined prevalent and incident lung cancers. Figure S3 shows a volcano plot of serum proteins associated with incident or prevalent lung cancer, with additional details on the findings provided below.

**<u>Cancer of lung and bronchus (LUC)</u>**

Cancer of the lung (LUC, ICD-10 code C34), is the most common type of cancer and a leading cause of cancer-related deaths in men and the second leading cause of cancer death in women[61]. The most significant risk factor for LUC is tobacco smoking[61], which increases the risk by a factor of 20. In the AGES study, 176 individuals were diagnosed with LUC: 20 participants (11 females) had a history of LUC at the baseline visit, while 156 participants (86 females) were diagnosed with LUC during the follow-up period. For incident LUC, 188 proteins were significantly associated at FDR < 0.05 when correcting for the standard covariates (Table S10). After adjusting for smoking status, eight proteins remained significant, including WAP four-disulfide core domain protein 2 (WFDC2, aka HE4), secretoglobin family 3A member 1 (SCGB3A1), C-type lectin family 3 member B (CLEC3B), contactin 3 (CNTN3), and growth differentiation factor 11 (GDF11, aka myostatin) (Table S10). Additional 28 proteins were associated with incidence LUC in a sex specific manner at FDR < 0.05 of which 2 remained stignificant after adjusting for smoking status, i.e. proprotein convertase subtilisin/kexin type 2 (PCSK2) in males only, and glutamate decarboxylase 1 (GAD1) in females only (Table S10). In total, 216 serum proteins were significantly associated with incident LUC at FDR < 0.05, when using the standard covariates (Table S10). Ten serum proteins remained significantly associated with incident LUC at FDR < 0.05 after accounting for sex, age, eGFR and smoking status including including for instance WFDC2, SCGB3A1 and CLEC3B (Table S10). Overall, 315 serum proteins were significantly associated with incident LUC at FDR < 0.10 (Table S10). Logistic regression analysis of prevalent LUC revealed six proteins that were associated with the disease at FDR < 0.05, using standard covariate adjustment, including for instance WFDC2 and CLEC3B, also associated with incident LUC (Table S11), and cellular tumor antigen p53 (TP53) (Table S11). After adjusting for smoking status, these proteins were no longer significantly linked to prevalent LUC at FDR < 0.05 (Table S11).

***Notable findings and functional highlights***: Due to the strong risk associated with tobacco smoking, we concentrated primarily on proteins that remained significantly associated with LUC (FDR < 0.05) after full adjustments, including age, sex, eGFR and smoking status (Table S12). This decision was influenced by the large number of proteins associated with LUC when adjusting only for age and sex. These proteins may be linked to LUC independently of the influence of smoking. Here, many of these proteins were differentially expressed in lung tumors and influenced by *cis*-acting pQTLs in our study (Table S12). The proteins WFDC2, SCGB3A1, and CLEC3B have compelling prior links to lung carcinogenesis, which underscores the biological plausibility of their associations with LUC in the current study. WFDC2 (aka HE4) is a well-established biomarker and functional oncogene in lung adenocarcinoma, where it has been shown to promote tumor cell proliferation and invasion[62,63]. In contrast, SCGB3A1 appears to act as a potent tumor suppressor: it is frequently silenced by promoter hypermethylation in early lung tumors, resulting in loss of its growth-inhibitory functions[64,65]. Finally, CLEC3B is implicated as a putative tumor suppressor: its down-regulation correlates with advanced stage and poorer prognosis in non-small cell lung cancer, potentially via modulation of tissue-remodeling and metastasis pathways[66,67]. Taken together, these markers map onto key molecular pathways in lung cancer development from loss of tumor suppression (SCGB3A1, CLEC3B) to active oncogenic promotion (WFDC2).

**Reference tables and figures**:

- Tables S10–S11 (excel): Complete association results for each protein at FDR thresholds of 0.05 and 0.10, for incident and prevalent cancers, respectively.
- Table S12 (excel): Summary characteristics of proteins with significant associations (FDR < 0.05).
- Fig. S3: Volcano plot showing the associations of serum proteins with both incident and prevalent lung cancer, with select proteins highlighted for illustration.

### Supplementary Note 5

#### *Female reproductive system cancers*

Cancers of the female reproductive system, such as those originating in the ovaries and uterus, typically begin in the pelvic region. Despite being anatomically distant, breast cancer is frequently included in discussions of the female reproductive system, and we will follow this convention here. In this study, we investigated the association of 7,523 serum proteins with cancers of the breast, ovaries, and corpus uteri. It is noteworthy that estrogen and other sex-specific hormones play a crucial role in the development of all these common cancers within the female reproductive system. Table S1 lists the number of individuals diagnosed with either prevalent or incident cancers of the female reproductive system in this study, while Table S6 offers descriptive statistics stratified by all diagnosed cancers of the female reproductive system. Figure S4 presents a multi-cancer volcano plot of serum proteins associated with incident or prevalent cancers of the female reproductive system, with additional details on the findings provided below.

**Breast cancer (BRC)**

Breast cancer (BRC, ICD-10 code C50) is the most common type of cancer and cancer-related deaths in women, with reported 25% new incidence of cancer in women worldwide[61]. Family history and environmental risk factors, as well as several modifiable risk factors connected to hormone usage, obesity, alcohol consumption and tobacco smoking, all have a significant role in the risk of BRC[68]. Rare coding variants in genes such as breast cancer 1 and 2 (*BRCA1* and *BRCA2*) affect 3% of all BRC cases, and genetic tests for these mutations are routinely utilized in BRC (as well as ovarian cancer, see below) risk screening[69]. In the AGES cohort, 231 individuals had BRC diagnosis: 124 individuals (122 females) had a BRC diagnosis before the first blood draw, while 107 individuals (103 females) were diagnosed with BRC during the follow-up period. After removing the four males from the analysis, two proteins, taxilin alpha (TXLNA) and Wnt family member 10B (WNT10B), were associated with incident BRC at FDR < 0.05 and remained significantly associated after adjusting for all covariates, including BMI, smoking status, and alcohol consumption (Table S10). Four proteins were associated with incident BRC at FDR < 0.10 (Table S10). After adjusting for standard covariates, 134 serum proteins were linked to prevalent BRC at FDR < 0.05, with 90 remaining significant after controlling for BRC risk variables such as BMI, smoking status, and alcohol consumption (Table S11). Additionally, after full adjustment, 10 more proteins were identified as significant at FDR < 0.05. In total, 144 serum proteins were associated with prevalent BRC at FDR < 0.05 (Table S11), while 197 serum proteins were associated with prevalent BRC at FDR < 0.10 (Table S11).

***Notable findings***: The serum protein WNT10B was positively associated with future breast cancer risk (Table S10), and its related network has previously been linked to survival and metastasis in triple-negative breast cancer[70]. Another Wnt family member, WNT5B, was associated with prevalent BRC (Table S11), highlighting the potential impact of Wnt signaling in BRC development. Indeed, endogenous Wnt pathway inhibitors are frequently downregulated during breast cancer development[71]. Other proteins of interest include fetuin B (FETUB), whose expression is stimulated by estrogen[72], suggesting a potential role in the development and progression of BRC under estrogenic influence. Several proteins associated with BRC in the observational analysis are influenced by *cis*-acting pQTLs (Table S12). Many of these proteins are differentially expressed in BRC-related tumors and/or tumors associated with other cancers of the female reproductive system (Table S12).

#### Cancer of the corpus uteri (CUC)

Cancer of the corpus uteri (CUC, ICD-10 code C54) is commonly known as endometrial cancer, which originates from the epithelial lining of the uterine cavity. Corpus uteri cancer (CUC) is the most common gynecological cancer in developed countries, with estimated 2.8% lifetime risk for females in the US[73]. Risk factors for CUC include the age at menopause onset, the balance between estrogen and progesterone levels, and obesity[74,75]. However, data on plasma levels of estrogen and progesterone are not yet available in the AGES study. In the AGES, 43 females were diagnosed with CUC: 29 were diagnosed before the blood draw, and 14 were diagnosed during the follow-up period. Eleven serum proteins were associated with incident CUC at an FDR of < 0.05 after standard covariate adjustment, with eight remaining significant following additional adjustment for BMI (Table S10). Additionally, three more proteins were significantly associated with incident CUC after adjusting for BMI, including cutA divalent cation tolerance homolog (CUTA), PAXIP1 associated glutamate rich protein 1 (PAGR1), and ubiquitin conjugating enzyme E2 E3 (UBE2E3) (Table S10). In total, 14 proteins were associated with incident CUC at an FDR of < 0.05, while 16 proteins were associated at an FDR of < 0.10 (Table S10). Three proteins, including the osteoclast-associated Ig-like receptor (OSCAR), NudC domain containing 2 (NUDCD2) and decorin (DCN), were linked to prevalent CUC at an FDR < 0.05 using standard covariate adjustments (Table S11). Among these, only OSCAR remained significant after further adjusting for BMI (Table S11). In total, nine serum proteins were associated with prevalent CUC at an FDR < 0.10.

***Notable findings***: Table S12 presents key characteristics of proteins linked to incident or prevalent CUC with FDR < 0.05 in the AGES study. Many of these proteins exhibit differential expressions in tumors of the corpus uteri, breast, and/or ovaries, and many are influenced by *cis*-acting pQTLs identified in the AGES (see Table S12). Emerging evidence suggests that PAGR1, UBE2S, and OSCAR may contribute to oncogenesis in hormone-driven female cancers, though the literature remains limited. For instance, *PAGR1* (aka PA1), a component of the TRiC/CCT chaperonin complex, has been shown to be expressed in breast tumors, and its levels associate with clinical outcomes in a cohort of invasive breast cancer patients[76]. Meanwhile, the ubiquitin–conjugating enzyme UBE2S (notably, more so than UBE2E3) is overexpressed in ovarian carcinoma and appears to promote tumor proliferation and therapy resistance via activation of

Wnt/β-catenin and PI3K/AKT/mTOR signaling[77,78]. Additionally, OSCAR, an innate immune receptor, has increased mRNA expression in BRC transcriptomic datasets and may facilitate malignant phenotypes[79], although functional and clinical data in OVC and CUC are currently scarce. In contrast, for CUTA, which is involved in metal-ion (e.g., copper) homeostasis, there is no clear evidence for a direct role in breast, ovarian, or endometrial cancers, despite the broader relevance of copper biology in cancer[80]. Taken together, these findings highlight diverse mechanisms including protein folding, ubiquitin-mediated regulation, inflammation, and ion homeostasis.

**Ovarian cancer (OVC)**

Ovarian cancer (OVC, ICD-10 code C56) is usually diagnosed at an advanced stage and is associated with high mortality rate[81]. In fact, among gynaecological malignancies, OVC, once diagnosed, has the greatest mortality rate[81]. Certain hormones, both endogenous and administered as medicine, enhance the risk of OVC, and some studies have linked obesity and smoking to an increased risk[82]. Furthermore, high-penetrance rare mutations in *BRCA1* and *BRAC2* cause OVC clustering in families[83,84]. A total of 22 individuals were diagnosed with OVC in the AGES study: six females had OVC at baseline, while 16 females developed OVC during the follow-up period. Two proteins were significantly linked to incident OVC at FDR < 0.05, i.e. erythropoietin receptor (EPOR) and eva-1 homolog B (EVA1B) (Table S10), but when BMI and smoking status were added to the adjustment model, only EVA1B remained significantly associated with incident OVC at FDR < 0.05 (Table S10). Fourteen serum proteins in total were linked to incident OVC at FDR < 0.10 (Table S10).

***Notable findings***: In the AGES study, only two proteins were associated with incident OVC, both of which are downregulated in either breast or endometrial tumors (Table S12). Prior studies have identified non-canonical roles for both EPOR and EVA1A in cancer, including potential relevance to ovarian (and other) tumors. In OVC cell lines such as A2780 and SKOV-3, EPOR is expressed and stimulation with erythropoietin (EPO) under hypoxic conditions promotes angiogenic factor release and enhances angiogenesis in endothelial cells, potentially contributing to tumor progression and chemoresistance[85]. Moreover, RNA interference-mediated knockdown of EPOR in A2780 cells reduces their invasiveness and *in vivo* tumor growth, suggesting a role for EPOR in supporting ovarian carcinoma growth[86]. On the other hand, EVA1A, a membrane-associated protein that regulates both autophagy and apoptosis, has been characterized as a tumor suppressor in other cancer contexts. It interacts with autophagy machinery (e.g., ATG16L1) to promote autophagosome formation and can induce cell death through autophagic and apoptotic mechanisms[87]. Although direct studies in ovarian cancer are limited, downregulation of EVA1A has been observed in multiple other tumor types[88], indicating a broader tumor-suppressive potential. Together, these findings imply a complex, context-dependent role for EPOR and EVA1A in tumor biology: EPOR may promote tumor survival and vascularization in the ovarian TME, while EVA1A appears to counterbalance tumorigenesis by activating autophagy-mediated cell death.

**Reference tables and figures**:

- Tables S10–S11 (excel): Complete association results for each protein at FDR thresholds of 0.05 and 0.10, for incident and prevalent cancers, respectively.
- Table S12 (excel): Summary characteristics of proteins with significant associations (FDR < 0.05).
- Fig. S4: A multi-cancer volcano plot summarizes the associations between serum proteins and both incident and prevalent cancers of the female reproductive system analyzed in this study, with selected proteins highlighted for illustration.

### Supplementary Note 6

#### *Skin cancer*

Cutaneous melanoma is the most prevalent and deadliest form of skin cancer, with a significant incidence rate of 3-6% in the United States[89]. Although sun exposure is the primary risk factor for melanoma, there are other types of melanomas that are not related to sun exposure[90]. In the present study, we investigated the association of 7,523 serum proteins with cutaneous melanoma. Table S1 lists the number of individuals diagnosed with either prevalent or incident melanoma in the AGES cohort, while Table S7 offers descriptive statistics stratified by all diagnosed melanoma cases. Unlike many of the cancer types mentioned above, patients with melanoma did not show a significant differential distribution of well-established cancer risk factors, such as smoking status, alcohol consumption, and obesity (Table S7). Figure S5 presents a volcano plot of serum proteins associated with incident or prevalent cutaneous melanoma, with additional details on the findings provided below.

##### Cutaneous melanoma (CMC)

The incidence of cutaneous malignant melanoma (CMC, ICD-10 code C43) is highest among fair-skinned populations due to sun exposure, with men experiencing higher mortality rates than women[91]. The risk of CMC is greatest among individuals of European ancestry compared with other population[92]. Globally, the incidence of CMC is rising due to population aging, while it is decreasing among younger individuals[92]. While sunlight exposure is the primary risk factor for melanoma, recent evidence suggests a positive correlation with obesity, alcohol intake, and tobacco smoking[93]. In the current study, 93 individuals were diagnosed with CMC: 14 prevalent cases (8 females) at the baseline visit, and 79 cases (42 females) that developed CMC over the follow-up period. Three serum proteins were linked to all incident cases of CMC with an FDR < 0.05 when standard covariate adjustments were applied including cholecystokinin (CCK), transmembrane protein 41B (TMEM41B) and sialic acid acetylesterase (SIAE) (Table S10). Additionally, seven more serum proteins were associated with incident CMC in a sex-specific manner, also at an FDR < 0.05, using standard covariate adjustments (Table S10). After additional adjustment for BMI, smoking status, and alcohol intake, nine proteins remained significantly (FDR < 0.05) associated with incident CMC (Table S10). In total, 10 proteins were associated with incident CMC at FDR of < 0.05, and 12 were associated at FDR < 0.10 (Table S10). Six serum proteins were associated with prevalent CMC at FDR < 0.05 using standard covariate adjustment (Table S11). Proteins such as fragile histidine triad diadenosine triphosphatase (FHIT) and chorionic somatomammotropin hormones (CSH1|CSH2) are examples of these associations. Two of these proteins, including FHIT, remain significantly associated with prevalent CMC after further adjusting for BMI, smoking status, and alcohol intake (see Table S11).

***Functional highlights***: Table S12 outlines key characteristics of proteins linked to CMC in the AGES study. Notably, two proteins, i.e., proprotein convertase subtilisin/kexin type 1 inhibitor (PCSK1N) and TMEM41B are downregulated in CMC tumors (Table S12). Like the gene encoding WWOX, which is associated with incident ESC (see above), the FHIT gene contains fragile sites or genomic regions prone to frequent chromosome breakage[15], often manifesting as rearrangements and deletions in cancer cells. Research on the roles of FHIT, SIAE, TMEM41B, and PCSK1N in skin cancer (especially melanoma) provides intriguing but incomplete insights. The *FHIT* gene, a well-known tumor suppressor, is notably reduced in melanoma: studies show diminished FHIT expression in melanoma cells, and restoration of FHIT can limit melanoma growth, suggesting its loss contributes to tumorigenesis[94]. Meanwhile, SIAE (sialic acid acetylesterase) is emerging as an immune-regulatory enzyme: in cancer cell lines, loss of SIAE increases acetylated sialic acid, which dampens NK cell–mediated cytotoxicity via the sialic-acid/Siglec pathway, suggesting that defective SIAE function may help tumor cells evade immune surveillance[95]. The TMEM41B protein, a transmembrane autophagy regulator, has been identified as essential for autophagosome formation and lipid mobilization[96], and recent data in breast cancer also link its regulation by miRNAs, implying a broader role in tumour cell survival under stress[97]. Together, these proteins reflect a spectrum of tumor-relevant mechanisms, from genomic instability and immune evasion to autophagy and protease regulation, but more targeted research is needed to establish their precise roles in melanoma onset and progression.

**Reference tables and figures**:

- Tables S10–S11 (excel): Complete association results for each protein at FDR thresholds of 0.05 and 0.10, for incident and prevalent cancers, respectively.
- Table S12 (excel): Summary characteristics of proteins with significant associations (FDR < 0.05).
- Fig. S5: A volcano plot summarizing the associations between serum proteins and both incident and prevalent malignant melanoma analyzed in this study, with selected proteins highlighted for illustration.

### Supplementary Note 7

#### *The impact of covariate adjustments and sex-specific analyses*

The most common modifiable risk factors for cancer include tobacco use, alcohol consumption, and excess body weight (overweight and obesity)[98,99]. In addition to adjusting for age, sex and eGFR, we accounted for the common risk factors when supported by epidemiological evidence linking them to specific types of cancer. Considering the established epidemiological associations between body height and the risk of PRC[100], and blood pressure and KIC[101], we included adjustments for these factors in our analysis (Tables S10-S11). Some cancer types were more influenced than others by the inclusion of additional adjustments for well-established risk factors specific to each cancer type, and in some cases additional proteins were detected (Figs. S6-S7).

The cancer type most affected by the inclusion of additional covariate adjustment was incident and prevalent LUC (Figs. S6I and S7G). This is not surprising, considering the high proportion of current smokers in the combined LUC patient group (38.8% vs. 11.4%) and the low proportion of never smokers (7.6% vs. 43.6%) (Table S5). In this study, 216 serum proteins were significantly associated with incident LUC (Table S10), and six proteins with prevalent LUC (Table S11), at FDR of $< 0.05$ when using standard covariates only. After adjustment for smoking, 10 proteins remained significant for the incident LUC, while no proteins remained significant for prevalent LUC. The proteins still significantly associated with incident LUC included WFDC2, SCGB3A1, CLEC3B, CNTN3, and GDF11 (Table S10). Notably, WFDC2, SCGB3A1, and CLEC3B are differentially regulated in tumors of the lung (Table S12). For instance, SCGB3A1 is specifically expressed in the lung and salivary gland (Table S12), and lower levels of this protein have been reported in non-small cell lung cancer, suggesting a role in modulating tumor progression[102,103]. Moreover, SCGB3A1 and WFDC2 are markers of early secretory cells in the lung conducting airway epithelium and may thus help identify distinct cell populations within tumors[104,105]. The serum proteins CLEC3B and GDF11 have been linked to tumor-host interactions, through their involvement in the epithelial–mesenchymal transition (EMT)[106] and the TME[107], respectively. Further, GDF11, a negative regulator of muscle growth and a member of the transforming growth factor-beta (TGF-β) superfamily, has been associated with muscle wasting in lung cancer-related cachexia in mouse models[108]. Given the strong influence of smoking on lung cancer, many proteins associated with the disease may reflect biological responses to tobacco exposure.

In certain types of cancer, including incident ESC, prevalent STC, prevalent REC, prevalent BRC, incident and prevalent CUC, incident OVC, and prevalent CMC, full adjustment led to a reduction in the total number of protein associations (Figs. S6-S7), though many associations remain. In contrast, for other cancers, full adjustment had little effect or led to the identification of additional protein associations, as seen for incident STC, incident and prevalent KIC and PRC, incident and prevalent BLC (Figs. S6-S7). Among proteins highlighted above, several remained robust to additional covariate adjustments beyond the standard set, including KLK3 and ACP3 for incident PRC, MSMB for prevalent PRC, and WFDC2 and SCGB3A1 for incident LUC, among others (Tables S10-S11). Proteins that became significant only after full adjustment included, for example, APOM for incident PRC, TNFSF4 (aka OX40L) for incident STC, POLR1C and CHST12 for incident PAC, TSTA3 for incident KIC, and PAGR1 and UBE2E3 for incident CUC (Table S10). Although the roles of POLR1C and TSTA3 in cancer remain unclear, the other proteins have been implicated in various aspects of tumor-host interactions[109-111]. For example, immunotherapy may increase OX40L levels, a stimulatory checkpoint on dendritic cells, while concurrently decreasing the expression of the inhibitory checkpoint PD-L1 on tumor cells[26]. This dual effect enhances the effectiveness of antitumor immunotherapy in gastrointestinal cancer[26]. Both POLR1C and CHST12 were differentially regulated in pancreatic tumors compared to normal samples (Table S12). For instance, high expression levels of TSTA3 in kidney tumors were associated with poor survival in KIC patients[112]. In summary, presenting results with both standard and full adjustment

(Figs. S6-S7) offers a more refined and comprehensive understanding of the findings, ensuring that potential confounding factors are adequately controlled.

Analyzing the sexes separately is important for fully understanding the distinct biological and molecular characteristics of cancers that are not inherently linked to a single sex. In sex-specific analyses, new protein associations were identified for COC, STC, REC, BLC, KIC, and CMC (Tables S10-S11, Supplementary Notes 2-6 above). These included proteins such as BCL2L14, noted above, which was positively associated with incident COC in males only and has been previously linked to colon tumorigenesis[113]. In females only, CUL1 was positively linked to incident STC, but elevated CUL1 levels have also been associated with poor prognosis in gastric cancer patients[114]. Other examples include CSF1, which was positively correlated with incident BLC in males only, and high CSF1 levels have been linked to poorer overall survival in BLC patients[115]. The proteins CUL1 and CSF1 may influence tumor-host interactions through distinct mechanisms, CUL1 by modulating the DNA damage response and apoptosis[114], and CSF1 through immune regulation of the TME[116]. ASMTL is another protein positively associated with incident CMC in females only (Table S10) and may contribute to melatonin biosynthesis in a sex-specific manner[117], a pathway linked to circadian regulation and tumorigenesis[118]. In the male-specific analysis of ESC, additional protein associations emerged, suggesting a male-dominant effect in protein associations within ESC in the AGES (Supplementary Note 2). Notably, significant protein associations were only observed for incident PAC when the sexes were analyzed separately, with no overlap between the findings in males and females (Table S10). Sex-specific associations between proteins and prevalent cancers were relatively rare, likely reflecting both the smaller number of prevalent versus incident cases and the fewer number of cancer types analyzed (Supplementary Note 1). Nonetheless, notable examples were observed, including NR3C2 and IL1R2, which were linked to prevalent BLC exclusively in females (Table S11, Supplementary Note 3). In summary, sex-specific biological differences such as hormonal, immune, or genetic factors, can uncover distinct molecular signatures, even when statistical power is reduced due to smaller sample sizes.

**Reference tables and figures**:

- Tables S10 (excel) – S11 (excel): Complete association results for each protein at FDR thresholds of 0.05 and 0.10, for incident and prevalent cancers, respectively.
- Fig. S6 illustrates how different covariate adjustments influence the number of proteins associated with future cancer risk.
- Fig. S7 depicts the effect of covariate adjustment on protein associations with prevalent cancers.

### Supplementary Note 8

#### *Cancers of any type*

We examined whether the 7,523 serum proteins were associated with any type of cancer (ATC), including malignancies not specified above, such as cancers of the larynx, brain, testis, and small intestine, as well as Hodgkin lymphoma and multiple myeloma, to name a few. A total of 1,916 individuals were diagnosed with some form of cancer, including 835 cases of prevalent cancers (n = 499 females) and 1,081 cases (n = 535 females) of incident cancers in the AGES cohort. This means that 684 patients had different cancer diagnoses compared to 1,235 individuals affected by the 13 cancer types discussed above. Table S8 (excel) lists ICD-10 codes for cancer types beyond the 13 primary analyses, and Table S9 summarizes characteristics of the 684 participants (1,916 total cases) with these cancers, comprising 772 diagnoses due to multiple cancers diagnosed in some individuals. In contrast to the strong associations commonly seen between the 13 prevalent cancers and lifestyle factors such as smoking, obesity, and alcohol consumption, the additional 684 cancer patients showed no such associations with these common risk factors (Table S9).

At an FDR < 0.05, and after adjusting for standard covariates, 269 serum proteins were associated with incident ATC (Table S13), while 38 proteins were linked to prevalent ATC (Table S14). After adjusting for the common covariates across different cancer types, including age, sex, eGFR, BMI, alcohol use, and smoking status, we found that 147 proteins remained significantly associated with incident ATC (Table S13), with additional 13 more serum proteins at FDR < 0.05 (Table S13). Further, 35 proteins remained significantly linked to prevalent ATC after full adjustment, with an additional 10 proteins identified at FDR < 0.05 (Table S14). The sex-specific analysis revealed nine additional proteins associated with incident ATC (Table S13), while another seven were associated with prevalent ATC in a sex-specific manner at an FDR < 0.05

(Table S14). A total of 291 proteins (320 aptamers) were associated with incident ATC at an FDR < 0.05 (Table S13), while 56 proteins (63 aptamers) were linked to prevalent ATC at FDR < 0.05 (Table S14). At FDR < 0.10, 449 proteins (489 aptamers) were associated with incident ATC (Table S13), whereas 99 proteins (112 aptamers) were associated with prevalent ATC (Table S14). Figure 5A–B in the main text presents volcano plots depicting the associations between serum proteins and incident versus prevalent ATC.

Several of the top proteins associated with prevalent and/or incident ATC, including TFF3 and KLK3, were also identified in the cancer-type specific analysis (see above). Notably, KLK3, which was linked to both prevalent and incident PRC (Tables S10-S11), was associated with incident and prevalent ATC only in males (Tables S13-S14), consistent with its specific expression in the prostate and its well-established role as a biomarker for PRC[51]. TFF3 has been proposed as a biomarker for various cancers, including breast, stomach, lung, prostate, and pancreatic cancer[119,120]. We found TFF3 to be associated with incident LUC and prevalent STC, PRC and BRC (Tables S10-11). Other notable serum proteins including WFDC2 and growth differentiation factor 15 (GDF15), which are a part of the Cancer Seek blood test panel for multi-cancer early detection[121], are associated with incident ATC (Table S13). In our cancer-type specific analysis, WFDC2 was linked to both incident and prevalent LUC (Tables S10-S11), while GDF15 was associated with incident LUC and prevalent REC (Tables S10-S11). Similarly, HAVCR1, associated with incident KIC (Table S10), was also linked to incident ATC, while FETUB, linked to prevalent BRC (Table S11), was found to be associated with prevalent ATC (Table S14). Interestingly, 43.2% of all proteins associated with incident ATC are predicted to be secreted, based on the HPA database[11], and compared to 19.1% of secreted targets identified across the entire aptamer-based array (Fisher's exact test OR = 4.9, P = $2\times10^{-16}$). Similarly, 40% of proteins associated with prevalent ATC are also predicted to be secreted (Fisher's exact test OR = 4.2, P = $2\times10^{-6}$). This is noteworthy because proteins secreted or shed into the interstitial and other biological fluids are increasingly studied as markers of primary tumors, metastasis, and the TME in both advanced and asymptomatic cancer stages[122,123].

**Reference tables and figures**:

- Table S8 (excel): Contains ICD-10 codes for cancer types not included in the 13 primary analyses.
- Table S9: Summarizes the characteristics of the additional 684 cancer patients.
- Tables S13 (excel) – S14 (excel): Present the associations between 7,523 serum proteins and incident and prevalent cancers of any type, respectively.
- Fig. 5A-B in the main text shows volcano plots depicting the associations between serum proteins and incident versus prevalent any type of cancer.
- Fig. 5C-D in the main text illustrates the overlap between results for any type of cancer and each of the 13 specific cancer types and shows that the cancer-associated proteins identified in this study are highly enriched among the most highly cited cancer genes in the literature.

### Supplementary Note 9

#### *Serum proteins linked to germline genetic susceptibility to cancer*

In this section, we have linked the lead SNPs from multiple meta-analyses of GWAS for each of the 13 cancer types mentioned above, as well as a pan-cancer study encompassing multiple cancers. Germline variation regulates circulating protein levels[124-127], enabling the identification of proteins with potential causal roles in specific cancer types. To explore these links, we integrated data on 7,523 serum proteins, with the latest meta-analyzed genome-wide association studies (GWAS) results across several cancer types. This included both pan-cancer GWAS spanning a broad range of cancer types and cancer-specific GWAS corresponding to the cancers analyzed in our primary observational study (see below). In total, we examined 980 lead SNPs across 13 cancer-specific GWAS, identifying 300 independent SNPs significantly associated ($P < 5\times10^{-5}$), acting in *cis* and/or *trans*, with 737 proteins, represented by 800 aptamers (Table S15). Unlike the cancer-type-specific patterns seen in the observational analyses, these genetically linked proteins often overlapped across cancer types: 210 proteins (28.5%) were linked to more than one cancer (Table S15), and 23 proteins were associated with four or more distinct cancer types (Table S15).

Several "protein hotspots" accounted for multiple genetic-protein associations, including loci at chr. 3p21.1 (*ITIH1*, *ITIH3*), 6p21.3 (*MHC* region), 9q34.2 (*ABO*), 12q24.1 (*SH2B3*), 14q32.1 (*SERPINA1*), 14q32.3 (*AKT1*), and 19q13.3 (*FUT2*) (Table S15). These loci suggest pleiotropic molecular effects across multiple cancer types, as well as associations with other diseases[124,126,127]. For example, *AKT1*, a canonical oncogene[128,129], and *SH2B3*, a tumor suppressor[128-130], lie proximal to the corresponding risk variants; however, their protein levels were unaffected in the present study. Interestingly, the GWAS risk variants rs2498797 and rs2498796 at the 14q32.3 hotspot, which influence KIC and CUC respectively, as well as numerous serum proteins (apart from AKT1) (Table S15), act as expression QTL (eQTL) across multiple tissues[8].

Similarly, the GWAS risk variant rs3184504 at 12q24.1, which influences CUC and numerous serum proteins (Table S15), is an eQTL for *SH2B3* in whole blood[8]. Proteins regulated in *cis* by known cancer risk loci include for example *CTRB2* for pancreatic cancer and *AZGP1* for colorectal cancer (Table S15). *CTRB2* is pancreas-specific[11], downregulated in tumors[7], and low circulating levels were associated with increased PAC risk (Table S15). *AZGP1* is a negative regulator of angiogenesis and may influence both the TME and EMT[131,132]. Among the 737 genetically influenced proteins (Table S15), we found 188 proximal *cis*-acting associations across the different cancer types (Fig. 6A, main text), involving 121 unique serum proteins, 29 of which were also identified in the observational analyses.

Pan-cancer genomic analyses further reinforce molecular overlaps across cancers[133,134]. In one GWAS of 18 cancers from over 400,000 individuals[134], 136 independent SNPs across 25 loci were associated with at least two cancer types. In a separate analysis, we found that 34 of these SNPs regulated 158 serum proteins (Table S16), including 39 additional proteogenomic associations, increasing the number of proteins regulated by cancer-associated loci to 776. Notably, several independent variants within the MHC region influenced the same proteins in opposite directions, consistent with findings from Rashkin et al.[134] and supporting both distinct and a shared molecular basis among cancers. The collective evidence presented here points to a substantial shared molecular basis across diverse cancer types.

*Proteogenomic profiling across various cancer types*

A GWAS of over 4,000 individuals with ESC of European ancestry identified 13 distinct genetic risk loci for ESC[135], seven of which influenced serum levels of 25 proteins in the AGES (Table S15). This included, for example, *cis* acting, i.e. genetic variants proximal to the protein´s cognate gene, effect on cathepsin B (CTSB) at 8p23.1 (rs10108511), which has previously been implicated in the genetic risk of ESC[100]. More specifically, a recent study found a putative functional variant at the 8p23.1 locus, i.e. rs55896564 ($r^2$ correlation with rs10108511 = 0.899), with allele-specific enhancer activity in an esophageal adenocarcinoma cell line[100], whereas CRISPR-mediated deletion of the enhancer region resulted in reduced expression of three colocalized genes, notably B lymphocyte kinase (BLK), nei like DNA glycosylase 2 (NEIL2), and CTSB[100]. Intriguingly, in our observational analysis, NEIL2 was among the serum proteins associated with incident ESC in the AGES (Table S10). The variant rs55896564 is found in the enhancer region at the 8p23.1 locus, and its G allele was linked to lower CTSB mRNA expression[100]. We found that the G allele for rs55896564 was linked to lower levels of CTSB in the AGES, as well as lower serum levels of three other proteins acting in *trans* (Table S15), while NEIL2, however, was not included. CTSB is specifically expressed in the esophagus, according to the HPA database[11]. Our proteomics platform also detects BLK, however neither CTSB nor BLK were associated with ESC in our study. Finally, a variant at another enhancer region harboring the ESC GWAS locus rs10423674 at chr19p13.11[100], was associated with serum levels of the cytokine receptor like factor 1 (CRLF1) (Table S15).

Whole genome association studies on European and Asian populations have identified 12 genome-wide significant risk loci to date for STC[136-138]. Of these, four variants influenced serum levels of 24 proteins in the AGES including several *cis*-acting pQTLs (Table S15). GKN2, which was significantly associated with prevalent STC in our study (Table S11), was linked to two independent SNPs, including rs760077, both acting in *trans* at GWAS risk loci for STC (Table S15). In this case, the allele (T-allele for rs760077) linked with low blood GKN2 levels is also associated with an increased risk of STC[10,136-138]. In the current study, serum levels of GKN2 were lower in prevalent STC (Table S15), implying that GKN2 deficiency may be causally related to STC. This is consistent with mouse knock-out and over-expression analyses relating GKN2 deficiency to STC pathogenesis[139]. However, the strong inverse relationship involving GKN2, which is specifically expressed in the stomach (Table S12), may be in part due to reduced serum levels of the protein in some STC patients who had undergone partial or total gastrectomy before protein measurements. The SNP influencing GKN2 serum levels also affected other proteins acting in *cis* including glucosylceramidase beta (GBA) and ephrin A1 (EFNA1) (Table S15). Interestingly, a recent multiplex mass spectrometry analysis of urine samples from patients with and without gastric cancer revealed elevated levels of EFNA1 in those with the disease[140]. However, the direction of effect between the allele associated with increased risk of STC and EFNA1 levels differs between that study and ours (Supplementary Table S15).

To date, the majority of GWAS studies have been conducted on colorectal cancer (CRC) rather than colon and rectal cancers separately. A GWAS of European and Asian ancestry subjects revealed numerous genetic risk loci for CRC[30,141]. In the AGES cohort, 31 of these genetic variants affected the levels of 171 proteins in the serum (Table S15). These included several proteins influenced by previously identified serum proteogenomic hotspots[127,142], such as those on chromosomes 6 (MCH locus) and chr. 19 (FUT2 locus) (Table S15). Several proteins from related families were identified, including members of the butyrophilin (BTN) and butyrophilin-like (BTNL) families (including BTN2A2, BTN3A2, BTNL8, BTNL9), which have been linked to colorectal and other cancers. Notably, BTN2A2 and BTNL9 exhibit altered expression or frameshift mutations in colon tumors[143], indicating potential roles in tumor immune regulation, whereas evidence for BTNL8 and BTN3A2 remains limited. These proteins may thus represent novel candidates for further study in cancer-associated immune pathways. Several other proteins with roles in colorectal cancer are highlighted: CDH1, a tumor suppressor[128,129], encoding E-cadherin, shows altered expression in colon tumors, implicating it in tumor progression and epithelial integrity[144]. CDH17, a gastrointestinal cadherin, is upregulated in CRC and linked to poor prognosis and Wnt/β-catenin signaling[145]. CSF1R, a receptor regulating tumor-associated macrophages[146], exhibits context-dependent effects, with expression in macrophages often correlating with worse outcomes. FGF19, an endocrine growth factor and a potential oncogene[128,129], is overexpressed in a subset of CRC and promotes tumor proliferation and survival, representing a potential therapeutic target. Together, these proteins reflect diverse mechanisms, including tumor suppression, cell adhesion, immune modulation, and growth factor signaling, relevant to colorectal cancer biology.

To date, 18 genome-wide significant risk loci for PAC (mainly pancreatic ductal adenocarcinoma) have been reported[147]. The GWAS findings, however, account for only 4% of the overall heritability of PAC[148]. We found that six distinct genetic risk loci for PAC influenced 116 serum proteins in total, with the ABO hotspot locus at chr. 9 influencing 106 of them (Table S15). Interestingly, two of the PAC risk loci, at chromosomes 13 and 16, were associated with varying serum levels of the protein pancreatic lipase related protein 1 (PNLIPRP1) (Table S15). Here, the two effect alleles raising the risk of PAC[147], were associated with lower levels of PNLIPRP1 in serum acting in *trans* (Table S15). This is intriguing because, in a study that identified a free fatty acid network linked to PAC, PNLIPRP1 was found to be downregulated in pancreatic tumors[149].

A recent large-scale multi-ancestry GWAS meta-analysis of 29,020 cases with kidney cancer, the majority of whom were of European ancestry, found 108 independent SNPs at 63 unique genomic regions[150], with some additional risk loci primarily associated with clear cell or papillary renal cell carcinoma.  We found 28 distinct genetic risk loci for KIC affecting 73 different serum proteins, which included several *cis* acting pQTL (Table S15). However, there was no overlap between the proteins affected by KIC associated GWAS risk loci and those identified through the observational study. Prostate cancer is among the most heritable cancers, with genetics accounting for 57% of the variability in risk[151]. A multi-ancestry meta-analysis of 156,319 PRC cases has yielded a total of 451 distinct genetic risk loci for PRC. We found that 142 distinct PRC genetic risk loci impacted the expression of 261 serum proteins (Table S15). For example, MSMB, which was associated with the genetic risk variant rs10993994 at chr. 10 in a *cis* acting manner (Table S15), was also one of the top proteins associated with prevalent PRC (Table S11). To present, 24 independent risk loci for BLC have been identified in a large-scale meta-analysis covering 32 independent studies that included 13,790 BLC cases[152,153]. Five of these risk loci were found to influence the serum levels of 31 proteins (Table S15), including REG1A and REG1B, which are also influenced by genetic risk loci associated with cancers of the digestive system, such as CRC and PAC (Table S15). Furthermore, both REG1A and REG1B were associated with prevalent STC in the AGES study (Table S11).

Lung cancer is primarily influenced by environmental factors, especially smoking, which accounts for approximately 80-90% of cases. Genetic factors contribute to a smaller extent, with heritability estimates ranging from 8% to 18%[154]. These estimates vary across different lung cancer subtypes; for instance, small cell lung carcinoma has a higher heritability (~10.5%) compared to squamous cell carcinoma (~5.2%)[154]. Notably, a subset of lung cancer cases occurs in individuals who have never smoked, and some of these cases exhibit familial clustering, suggesting a genetic component[155]. In a recent cross-ancestry and pan-cancer GWAS studies, 30 genetic risk loci for LUC were identified[134,156]. In the AGES study, 21 of these risk loci affected the levels of 170 serum proteins (Table S15). One of the strongest *cis*-acting pQTL signals was found for junctional adhesion molecule-like protein (JAML) (Table S15), a recently implicated target for agonist-induced cancer immunotherapy that enhances both CD8 and γδ T-cell immunity[157]. Additional strong *cis*-acting genetic risk factors for LUC included von Willebrand factor A domain containing 1 (VWA1), EFNA1, and cathepsin H (CTSH) (Table S15). Lastly, SCGB3A1 serum levels were influenced in *trans* by a genetic risk locus for LUC (Table S15), and this protein was also linked to incident LUC in the AGES study (Table S10).

A large-scale GWAS of BRC comprising 122,977 cases of European ancestry and 14,068 cases of East Asian ancestry revealed 115 common risk loci, accounting for approximately 15% of BRC heritability[158]. Here, 42 of the BRC risk loci were linked to varying levels of 74 proteins in serum in the AGES cohort (Table S15). These included for instance, tumor proteins 53 (TP53) and 63 (TP63), tumor protein p53 inducible protein 3 (TP53I3) and members of the butyrophilin subfamilies. A GWAS of 12,906 endometrial cancer cases of European decent identified 17 independent genetic risk loci of CUC[159]. Nine of these variants influenced the levels of 74 proteins in serum, including strong *cis*-regulated proteins such as gliomedin (GLDN) and sorting nexin 11 (SNX11) (Table S15). Several common genetic variants have been associated with OVC through GWAS in individuals of European ancestry[160,161], resulting in identification of 30 distinct risk loci linked to the disease. Eight of these genetic variants influenced the levels of 27 serum proteins within the AGES cohort, primarily acting in *trans* (Table S15). This includes, for example, C-X-C motif chemokine ligand 16 (CXCL16), which has been identified as a soluble biomarker associated with poor prognosis in ovarian cancer[162].

A large meta-analysis study of 36,760 cases of CMC of European ancestry found 68 independent genetic variants associated with the disease[163]. Of these, 12 SNPs affected serum levels of 16 proteins including agouti-signaling protein (ASIP), melanocyte protein PMEL (PMEL) and lactase-phlorizin hydrolase (LCT) (Table S15). Proteins influenced by genetic risk factors for CMC include, for example, the agouti-signaling protein (ASIP) and pre-melanosome protein (PMEL) (Table S15). In a previous study[142], we demonstrated through a forward two-sample MR analysis that ASIP is causally linked to malignant melanoma ($P = 1\times10^{-17}$). Notably, the melanoma risk allele for the lead GWAS SNP rs910873, was associated with higher serum levels of ASIP, which is located 314 kb downstream of rs910873 (Table S15). ASIP acts as a competitive inhibitor of MC1R[164], and as such, is strongly implicated in the biological mechanisms underlying melanoma risk. We note that ASIP was not associated with either prevalent or incident CMC in our observational analysis.

**Reference tables and figures**:

- Tables S15 (excel) – S16 (excel): Associations with genetic risk factors for each individual cancer type and for the pan-cancer analysis, respectively.
- Fig. 6A in the main text presents a volcano plot depicting the cis-regulatory effects of germline cancer risk loci on serum proteins, with results color-coded by cancer type.
- Fig. 6B-C in the main text shows the overlap between proteins identified in the observational analyses (incident and prevalent cancers) and those linked to genetic cancer risk through proteogenomic analysis, as well as the enrichment of these proteins among top-ranked cancer genes based on literature citation data.

### Supplementary Note 10

#### *Integrated network analysis of cancer-associated proteins*

The central dogma of biology describes how information flows from DNA to proteins, which then exert their effects on biological processes and phenotypes, including diseases, through interconnected biological networks. We identified the first circulating co-regulatory networks in humans derived from 4,786 serum proteins[124], linking them to the genome in an unbiased manner and elaborated on those findings to highlight their connections to a broad range of both current (prevalent) and future (incident) diseases, as well as their ability to predict overall and disease-specific survival[124]. More recently, we reconstructed the circulating causal protein network (CPN) through causal inference analysis of 7,523 serum proteins, identifying 185 subnetworks with at least 10 protein members, each collectively interacting with 5,611 target proteins[165]. Overall, there was a significant overlap between the circulating CPN and the co-regulatory networks, despite fundamental differences in the methodologies used for their reconstruction[165]. However, CPNs offer further insights into causal relationships between proteins that are not apparent in co-regulatory networks, as they can effectively distinguish causation from mere correlation. Importantly, the subnetworks from each network type consist of protein nodes synthesized in various tissues throughout the body, suggesting that their connections within these networks represent relationships that occur both within individual tissues and across tissue boundaries.

We first evaluated the enrichment of all cancer-associated proteins, those linked to incident or prevalent cancers across the 13 specific cancer types and ATC, together with proteogenomic findings, within previously published serum protein co-regulatory networks[124]. Since the co-regulatory network was generated from the 5K aptamer-based platform, only aptamers and corresponding serum proteins shared between the 5K and the 7K platform used in this study could be considered. In some cases, this led to a limited number of proteins likely influencing somewhat the outcome of the enrichment analysis. Table S17 highlights the enrichment of cancer-associated serum proteins in co-regulatory networks, showing a significant enrichment of many of these protein signatures in specific protein modules. Although individual proteins associated with incident or prevalent cancer show relatively little overlap, proteins from these distinct conditions share certain co-regulatory network modules (Table S17). More to the point, the protein modules most frequently enriched for various cancer-associated proteins were PM26 and PM27 (Table S17), both from the same supercluster of proteins and each comprising over 370 proteins[124]. These serum protein co-regulatory modules have been linked to cardiovascular and metabolic diseases, as well as overall and disease-specific survival[124]. Due to the shared protein signatures across individual cancer types, we grouped all proteins associated with new-onset cancer together and those linked to prevalent cancer separately. We then analyzed their enrichment within the co-regulatory network, revealing a significant enrichment in modules PM26 and PM27 (Fig. S11A, B). Further, these overlaps were also evident in the group with a history of any type of cancer, where proteins associated with incident or prevalent ATC were significantly over-represented in the protein modules PM26 and PM27 (Fig. S11C, D). The clustering of serum proteins linked to different types of cancer within the same co-regulatory modules suggests both functional commonalities and a shared, coordinated regulation of cancer-related protein expression.

Next, we analyzed the enrichment of cancer-associated proteins identified in this study within the recently characterized 185 CPN subnetworks[165], which comprise network regulators and their corresponding network targets ($n \geq 10$). Importantly, both studies utilized the 7K platform, enabling direct comparison. We analyzed the enrichment of cancer-associated proteins within specific CPN subnetworks. Many cancer-associated proteins were highly enriched in the CPN networks, with a particularly notable enrichment for those proteins linked to genetic cancer risk (Table S18). Overall, 18 CPN subnetworks are enriched for proteins associated with future cancers (mostly from LUC and ATC), 4 for prevalent cancers, and 42 for proteins linked to the genetic risk of cancer (Table S18). A substantial overlap of CPNs exists among these different groups of cancer-associated proteins (Table S18).

Considering the predictive power of biological networks[124], we highlight the CPN networks enriched for various types of incident cancers along with their interconnectedness (Fig. S12A, B). Moreover, within solid tissues, a subset of these networks adopts a cascade-like configuration (Fig. S12C). For example, several network regulators converge on PTPN11 (aka SHP2) (Fig. S12C), a protein encoded by a well-established canonical oncogene[128,129], which regulates 83 serum proteins including the oncogenic protein AKT1[165], and is associated with prevalent BRC in this study (Table S11). Although these network regulators linked to PTPN11 have not been directly implicated in this study, they represent potential candidates in cancer biology. We analyzed the enrichment of the corresponding network regulators (Table S18), in functional and physical protein-protein interactions (PPI) within solid tissues using the STRING database[166]. This analysis revealed a highly significant enrichment in interaction levels (6 expected edges vs. 29 observed edges; $P = 8 \times 10^{-11}$), forming seven distinct PPI clusters (Fig. S12D). These clusters included proteins involved in complement activation, protein folding, proteasome core complex, platelet development, NK-cell lectin-like receptor binding, allograft rejection, and vault protein inter-alpha-trypsin domain. Consistent with this, analysis using the STRING database[166] revealed that the network regulators of the combined CPN subnetworks, enriched in PPI interactions, were also enriched in pathways such as the complement and coagulation cascades ($FDR = 4\times10^{-14}$) and natural killer cell lectin-like receptor binding (FDR =0.007), as well as in tissues including liver ($FDR = 3\times10^{-9}$), bone marrow cell ($FDR = 3\times10^{-6}$), digestive gland ($FDR = 3\times10^{-8}$), viscus (FDR = 0.002), skeletal system (FDR = 0.0003), and endocrine gland (FDR = 0.0009). The CPNs enriched in the physical PPI networks primarily consist of network regulators enriched with proteins linked to incident cancers and/or genetic risk factors for cancer (Table S18). In summary, these findings suggest that these network regulators interact more frequently than expected by chance and that their interactions in serum mirror similar processes occurring in solid tissues.

While regression analyses highlighted an overlap between proteogenomic findings across various cancer types and to many of the observational findings (see above), the significant shared enrichment of these cancer-associated signatures across multiple CPN networks provides additional depth and context to these findings. These networks include regulators of many proteins and are strongly reflected in functional and physical protein-protein interactions in solid tissues. These findings provide new insights into modeling the complex etiology of different cancers and can help support the development of concrete hypotheses for follow-up research.

**Reference tables and figures**:

- Tables S17 (excel) - S18 (excel): Enrichment of cancer-associated proteins from the observational analysis within co-regulatory networks and circulating causal protein networks, respectively.
- Fig. S11: Overlap between serum protein co-regulatory networks and cancer-associated proteins.
- Fig. S12: Network visualization of the circulating causal protein networks (CPNs) enriched for various cancer-associated serum protein signatures.

### Supplementary Note 11

#### *Causal inference analysis across different types of cancer*

*Overview of the Mendelian randomization strategy*: In this section, we evaluated potential causal relationships between the serum proteome and cancer by conducting a proteome-wide forward two-sample MR analysis[167], using *cis*-acting pQTL instruments identified in the AGES cohort. We leveraged summary statistics from large cancer GWAS consortia (Table S19) and comprehensive *cis*-acting pQTL data[167], to test whether genetically predicted circulating protein levels influence cancer risk (protein → cancer). The causal tests were performed for 2,062 *cis*-acting pQTL instruments across multiple cancers. Results for all cancers are summarized in Table S20 (FDR < 0.10), while below we present a selection of key findings grouped by organ system.

##### I. Digestive system cancers

<u>*Colorectal cancer*</u>

**<u>GREM1 and CHRDL2</u>** - Many of the 22 proteins with potential causal relationships to CRC at FDR <0.05 (Table S20, Fig. S13), either protective or risk-enhancing, identified through the MR analysis play diverse roles in cancer biology. For example, Gremlin-1 (GREM1), which shows a positive and potentially causal association with CRC (Fig. S13), functions as a potent oncogene across multiple cancer types, most notably in CRC, where it promotes motility and invasion through EMT[168]. As a BMP antagonist, GREM1 drives tumor progression via multiple mechanisms, including activation of the PI3K/AKT/mTOR pathway and inhibition of BMP2 signaling[168]. Elevated GREM1 expression is associated with poor prognosis and advanced metastatic disease, and it also influences other cancers such as bladder, lung, esophageal, and glioma[169]. Another BMP antagonist chordin-like 2 (CHRDL2) is positively and potentially causally linked to CRC (Fig. S13), previously found to enhance cancer stem-cell characteristics in CRC[170]. CHRDL2 enhances chemotherapy and radiation resistance by activating DNA damage response pathways[170]. In organoid models, it increases stem-cell marker expression and suppresses differentiation through the WNT pathway[170]. Its overexpression is associated with poor clinical outcomes and treatment resistance[170]. Thus, prior evidence implicating these two BMP antagonists in CRC development is consistent with our findings, which indicate a positive and potentially causal association with the disease.

**DARS1, GSS and GSR** - The protein aspartyl-tRNA synthetase (DARS1) showed a positive and potentially causal association with CRC in our study (Fig. S13). DARS1 is overexpressed in multiple cancer types and has been implicated in tumor development[171,172]. It is notably upregulated in myeloproliferative neoplasms, renal cell carcinoma, glioblastoma, and gastrointestinal cancers[171]. Mechanistically, DARS1 promotes tumor growth through interactions with the Hippo signaling pathway and modulates $CD4^+$ and $CD8^+$ T cells, thereby fostering tumor-associated inflammation and progression within the immune TME[171]. The glutathione metabolism enzymes GSS and GSR exhibit positive and negative causal relationships with CRC, respectively (Fig. S13). Both proteins influence cancer progression by modulating the cellular antioxidant system[173]. Elevated GSS levels enhance chemotherapy resistance and protect cancer cells from oxidative stress, whereas GSR expression with CRC stage is low in 95% of stage I patients but high in 89% of stage III patients[174]. Increased glutathione pathway activity enables cancer cells to withstand treatment-induced oxidative damage[174]. These findings are consistent with our GSS results (Fig. S13), while the role of GSR appears more complex and likely context dependent.

**LGALS4 and HHIP** - Our MR analysis identified galectin-4 (LGALS4) as positively associated with CRC risk (Fig. S13). However, experimental evidence indicates that LGALS4 acts as a potent tumor suppressor in CRC[175]. It inhibits glycolysis, promotes apoptosis, and suppresses β-catenin signaling[175]. Overexpression of LGALS4 reduces cell proliferation by approximately 50%, doubles apoptosis, and increases sensitivity to 5-fluorouracil treatment[175]. In addition, LGALS4 modulates the Wnt/β-catenin and IL-6/NF-κB/STAT3 pathways to inhibit tumor progression[176]. Another protein showing a positive causal relationship with CRC in our analysis (Fig. S13), is reported to act as a tumor suppressor across multiple cancer types is hedgehog-interacting protein (HHIP)[177], a negative regulator of the Hedgehog signaling pathway. Overexpression of HHIP markedly inhibits cell proliferation and invasion while reducing metastatic potential. High HHIP expression has also been associated with longer relapse-free survival and decreased metastasis[178]. Thus, our MR findings appear inconsistent with prior experimental results for both LGALS4 and HIPP.

**NRG4** - Finally, neuregulin 4 (NRG4), which appears to be potentially causally protective for CRC (Fig. S13), has been reported to exert protective effects in cancer biology[179]. NRG4 acts as a survival factor for colonic epithelial cells by blocking TNF-induced apoptosis, suppresses metastasis through inhibition of the ERBB4–YAP1 pathway[180,181], and reduces inflammatory responses while promoting cellular homeostasis[180]. Loss of NRG4 expression is frequently observed in advanced disease stages, whereas higher expression is associated with improved survival[179]. These observations are consistent with our finding that NRG4 may be causally protective against CRC development.

*Stomach cancer*

**HSPA1A and RET** - Our MR analysis identified 19 protein associations with STC at FDR < 0.05, and 41 at FDR < 0.10 (Table S20), several of which have been previously linked to gastric cancer. Three aptamers targeting heat shock protein family A member 1A (HSPA1A) showed negative associations with STC or PRC in our MR analysis (Table S20, Fig. S14). HSPA1A is significantly upregulated in STC and plays a key role in treatment resistance[182]. The HSPA1A protein is strongly stained in gastric cancer and correlates with treatment outcomes[183]. Research indicates that conducting second rounds of HIPEC or chemotherapy at least 24 hours after initial treatment is optimal to minimize tumor cell resistance mediated by heat shock proteins[182]. The RET proto-oncogene (RET)[128,129], was positively and causally linked to STC (Fig. S14), with significant implications in gastric cancer[184]. Activation of RET may represent one of the molecular pathogenic mechanisms in gastric inflammatory and tumoral diseases. The protein is detected in gastric cancer specimens and is associated with phosphorylated epidermal growth factor receptor[185]. Germline mutations in RET cause multiple endocrine neoplasia syndromes and contribute to various cancer types including gastric cancer while RET rearrangements have been detected in multiple human cancers including gastric carcinomas[185]. While our findings for HSPA1A may appear contradictory, our results for RET are consistent with previous reports identifying it as a tumor activator.

**KLK10 and IL6R** - Kallikrein-related peptidase 10 (KLK10) was causally associated with an increased risk of STC (Table 20, Fig. S14). KLK10 has been identified as an independent biomarker of poor prognosis in gastric cancer patients[186]. Its expression is significantly elevated in approximately 70% of gastric cancer samples compared with normal gastric tissue, with high KLK10 levels correlating with lymph node metastasis, greater depth of invasion, and poor histological differentiation[186]. Patients with positive KLK10 expression demonstrate increased risk for relapse, metastasis, and death and urinary KLK10 levels can distinguish operable from inoperable gastric cancer cases[187]. The interleukin-6 receptor (IL6R), which mediates signaling of IL-6, was positively associated with STC in our MR analysis (Table S20, Fig. S14). IL6R plays a key role in inflammation-driven gastric carcinogenesis[188]. IL-6 signaling through both membrane-bound IL6R and its soluble form (sIL6R) promotes tumor development[188], with *trans*-signaling via sIL6R being particularly important in inflammation-associated cancers, including STC[189]. Elevated IL-6 levels correlate with tumor aggressiveness and metastatic potential, while the IL-6/IL6R pathway modulates the TME to support cancer cell survival, proliferation, and invasion[189,190]. Our findings are consistent with these reported roles of KLK10 and IL6R in STC.

**TLR3 and IL17RB** - Two inflammation-related proteins, toll-like receptor 3 (TLR3) and interleukin 17 receptor B (IL17RB), were found to be potentially causally related to STC (Table S20, Fig. S14). TLR3 demonstrates a dual role in gastric cancer, capable of both promoting tumor regression and facilitating cancer progression[191]. In gastric cancer, TLR3 overexpression is associated with poor prognosis[192]. The receptor can activate both antitumor immune responses and pro-tumor pathways depending on cellular context[191,193]. TLR3 signaling can induce microRNA expression that targets DNA methyltransferases, leading to reactivation of tumor suppressor genes[194]. However, it can also promote tumor recurrence, metastasis, and therapy resistance through production of pro-tumor cytokines[193]. IL17RB shows strong associations with gastric cancer progression and cancer cell stemness[195]. Its expression is significantly elevated in gastric cancer tissues and correlates with poor prognosis[195]. IL-17B/IL-17RB signaling promotes gastric cancer stemness through activation of the AKT/β-catenin pathway[195,196], thereby enhancing tumor cell growth, migration, and EMT[195,196]. Notably, IL-17B is upregulated in patient serum rather than in tumor tissue, suggesting a systemic inflammatory response[196]. This signaling axis therefore represents a potential therapeutic target in gastric cancer. Our MR findings are consistent with these reports, as IL17RB showed a positive association with STC, whereas TLR3 exhibited a negative association, potentially reflecting the dual role of TLR3 in tumor biology (Table S20, Fig. S14).

#### II. Genitourinary system cancers

##### _Prostate cancer_

**MSMB** - The MR analysis identified 37 putatively causal protein associations with PRC at FDR < 0.05 and 50 at FDR < 0.10 (Table S20). Selected examples of significant findings at FDR < 0.05 are highlighted below. Notable findings include microseminoprotein-beta (MSMB), which has a well-established tumor suppressor role in PRC[197,198]. This protein is normally abundant in prostatic secretions but becomes significantly downregulated in prostate tumors in correlation with tumor grade. Patients with tumors expressing high MSMB levels show significantly reduced risk for biochemical recurrence after radical prostatectomy[197,198]. Remarkably, aggressive prostate tumors suppress MSMB synthesis not only in tumor tissue but also in adjacent benign prostate tissue, explaining decreased serum MSMB levels in PRC patients[197,198]. Moreover, genetic variants within the *MSMB* locus are well-established determinants of PRC susceptibility, supporting a causally protective role of MSMB in disease development[53]. Our results align with previous evidence suggesting a causal protective role of MSMB in prostate cancer (Table S20, Fig. 7C main text).

**CDH1** - The protein E-cadherin (CDH1) is positively and potentially causally linked to PRC and consistent with our observational findings for incident ATC (Table S20, Fig. 7C main text). However, CDH1 functions as a critical tumor suppressor in PRC through multiple mechanisms[199]. Loss of E-cadherin expression promotes prostate cancer progression and metastasis by dysregulating β-catenin signaling. During prostate development, CDH1 promoter methylation naturally occurs to facilitate prostatic bud outgrowth[200], whereas inappropriate hypermethylation in cancer creates a permissive environment for tumor progression. The CDH1 -160 C/A polymorphism is associated with increased prostate cancer risk, with A allele carriers showing elevated risk compared to CC homozygotes[201]. Although CDH1 is classically considered a tumor suppressor within epithelial tissues, our MR and observational findings indicate a positive causal association between circulating CDH1 and prostate cancer risk. This apparent discrepancy between tissue and circulating biology may reflect increased shedding of CDH1 from epithelial cells, consistent with EMT and early tumorigenic remodeling.

**PDLIM5** - PDZ and LIM domain 5 (PDLIM5) was the second most strongly putatively causal protein associated with PRC in our study (Table S20, Fig. 7C main text) and has been implicated as a potential oncogene in PRC[202,203]. This cytoskeleton-associated protein is overexpressed in prostate cancer tissues compared to normal prostate tissue, with expression correlating strongly with Gleason score, tumor metastasis, and biochemical recurrence[202]. PDLIM5 promotes cancer cell proliferation, migration, and invasion by regulating the AMPK pathway and EMT[202]. Our findings are in line with prior reports linking PDLIM5 to an increased risk of PRC.

**HDGF** - We find that both aptamers targeting the hepatoma-derived growth factor (HDGF), a growth factor with mitogenic and angiogenic activity, are causally related to PRC (Table S20, Fig. 7C main text). HDGF functions as a survival-related protein by up-regulating anti-apoptotic proteins cyclin E and BCL-2 while simultaneously downregulating pro-apoptotic protein BAX[204], thereby promoting cell viability and proliferation. Additionally, HDGF enhances migration and invasion through activation of phospho-AKT and NF-κB signaling pathways, which have been implicated in cancer cell metastasis and progression. Thus, our finding that HDGF is putatively causally related to prostate cancer is consistent with its established roles in promoting cell proliferation, invasion, and tumor progression.

**FGF7** - Fibroblast growth factor 7 (FGF7) was negatively associated with PRC risk in our MR analysis (Table S20, Fig. 7C main text) and is known to function as a paracrine growth factor in this disease[205-207]. It is produced by prostate stromal cells and specifically activates FGFR2/IIIb receptors on epithelial cells[205]. FGF7 expression is particularly associated with hormone-insensitive prostate tumors[207]. Although FGF7 is considered oncogenic in certain contexts, our MR finding that genetically higher circulating FGF7 is protective for prostate cancer may reflect context-dependent biology, differences between circulating versus tissue-level effects, or isoform-specific functions. This observation is biologically plausible and underscores the complexity of growth factor signaling.

**PRSS3** - Mesotrypsin (PRSS3) showed a negative and potentially causal association with PRC risk in our MR analysis (FDR < 0.05), but a positive association with CRC (FDR < 0.10) (Table S20, Fig. 7C in main text). PRSS3, which is primarily expressed in the pancreas, promotes PAC cell proliferation, invasion, and metastasis by upregulating VEGF through PAR1-mediated ERK signaling[208]. In PRC, elevated PRSS3 expression is linked to metastasis, recurrence, and poor prognosis[209]. Mechanistically, PRSS3 facilitates tumor progression *via* activation of a broader protease network, including kallikrein-5[210]. Although genetically predicted higher circulating

PRSS3 levels appear causally protective for PRC risk, experimental data indicate that PRSS3 expression within tumor tissue promotes invasion and metastasis. This apparent discrepancy likely reflects context-specific effects: circulating PRSS3 may contribute to systemic protease homeostasis and protect against tumor initiation, whereas tumor-localized PRSS3 enhances extracellular-matrix degradation and oncogenic signaling.

*Bladder cancer*

**GSTM1, GSTM3, GSTM4 and LRIG1** - Our MR analysis identified the glutathione S-transferases (GSTM): GSTM1, GSTM3, and GSTM4 as inversely associated with bladder cancer risk at FDR < 0.05 (Table S20), suggesting a protective role. GSTM1 and GSTM3 were also causally associated with BRC in a directionally consistent manner at FDR < 0.10 (Table S20). These proteins belong to the mu class of GSTM, a gene cluster on chromosome 1p involved in detoxification of electrophilic and carcinogenic compounds (including that from tobacco use). Prior epidemiological studies have consistently linked the GSTM1-null genotype with increased bladder cancer susceptibility[211], particularly among smokers, and a polymorphism in intron 6 of GSTM3 has also been associated with elevated bladder cancer risk[212]. In fact, tobacco smoking is the single most frequent risk factor for BLC[55]. Our findings therefore provide genetic evidence that higher circulating levels of these GSTM proteins may protect against bladder carcinogenesis, reinforcing the biological plausibility of detoxification pathways as central in modulating bladder cancer risk. In addition to the GSTM family, we also observed an inverse MR association for leucine rich repeats and immunoglobulin like domains 1 (LRIG1), suggesting it may play a protective role in bladder carcinogenesis. LRIG1 encodes a membrane protein that negatively regulates the ErbB receptor tyrosine kinase axis (notably EGFR / HER family) by enhancing receptor ubiquitylation and degradation and thus may act as a tumor suppressor. In bladder cancer, LRIG1 is frequently downregulated, and forced overexpression suppresses cell proliferation, migration, invasion, and in vivo tumorigenesis[213]. While no direct mechanistic connections between LRIG1 and glutathione S-transferases (GSTM1, GSTM3, GSTM4) have been demonstrated, the convergence of detoxification pathways and growth factor signaling is attractive as a hypothesis: for example, reduced mitogenic signaling may lower oxidative stress or demand for detoxification, or conversely, GST-mediated redox balance may modulate signaling thresholds. Our MR findings thus implicate LRIG1 as a potentially independent but complementary tumor suppressor axis in bladder cancer, meriting deeper integrative and mechanistic exploration.

#### III. Female reproductive system cancers

##### _Breast cancer_

**SNUPN and ANXA4** - Our MR analysis identified 17 proteins associated with BRC at FDR < 0.05 and 27 proteins at FDR < 0.10 (Table S20). Many proteins with potential causal links to BRC identified in this study have previously been implicated in BRC or other malignancies, acting as biomarkers, oncogenes, tumor suppressors, or regulators of metastasis. For instance, snurportin 1 (SNUPN) has been identified as a negatively causal, protective protein in BRC[214], consistent with our findings (Table S20, Fig. S15). Another study has reported potential clinical applications of SNUPN in acute lymphoblastic leukemia[215]. Annexin A4 (ANXA4) is positively associated with BRC in our MR analysis (Table S20, Fig. S15). It has been reported to be upregulated in breast tumors, where it activates JAK/STAT3 signaling, correlates with poor survival, and promotes tumor growth, invasion, and chemoresistance[216,217]. Moreover, knockdown of ANXA4 has been shown to reduce cell migration in both ovarian and breast cancer models[217]. These functional findings on ANXA4 are consistent with the potentially positive causal association of the protein with BRC observed in our study.

**TAGLN and PARK7** - Two aptamers target transgelin (TAGLN), which showed a potentially causal protective effect against BRC in our study (Table S20, Fig. S15). Consistently, previous studies have identified TAGLN as a metastasis suppressor, with reduced expression associated with increased metastatic potential in BRC[218]. Our MR analysis identified Parkinsonism-associated deglycase (PARK7) as potentially causally protective against BRC (Table S20, Fig. S15), in line with findings from a previous MR study in an independent cohort[214]. It acts as a negative regulator of PTEN and PKB/Akt phosphorylation, thereby modulating cell survival and apoptosis[219]. PARK7, a key regulator of oxidative stress, influences tumor progression, cell survival, migration, and metastasis in BRC[220]. Higher PARK7 levels are generally associated with lower BRC risk and improved therapeutic response to therapy[221].

**UBA7 and CTSF** - The ubiquitin-like modifier-activating enzyme 7 (UBA7) was negatively associated with BRC in our MR analysis (Table S20, Fig. S15). This aligns with previous reports showing that UBA7 is downregulated in BRC, where low expression correlates with poor prognosis and more aggressive tumor characteristics[222]. UBA7 has also been proposed as a potential diagnostic and prognostic marker in other malignancies[223]. Finally, cathepsin F (CTSF) showed a potential positive causal association with BRC in our MR analysis (Table S20, Fig. S15), consistent in direction with recent evidence causally linking CTSF to BRC in another study[214]. CTSF plays a central role in the lysosomal pathway and has been suggested as a diagnostic marker for various cancer types[224-226]. Additionally, an mRNAsi-based screening study predicted CTSF as a poor prognostic factor for basal-like BRC[227].

#### IV. Skin cancer

*Cutaneous melanoma*

**ASIP, ERBB3 and ARG1** - In the current study, 10 circulating proteins were causally linked to CMC at FDR $< 0.05$ and 13 proteins at FDR $< 0.10$ (Table S20). According to prior findings, ASIP, oncogene erb-b2 receptor tyrosine kinase 3 (ERBB3), and arginase 1 (ARG1) exhibit the strongest associations with melanoma (see below). The ASIP protein exhibited the strongest causal association with CMC in this stdy (Table S20, Fig. S16). Genetic variants in ASIP are strongly linked to melanoma risk and the etiology of other skin cancers[228,229], with certain haplotypes substantially increasing melanoma susceptibility, particularly in individuals with lighter pigmentation traits[230]. ASIP is a secreted protein that antagonizes MC1R signaling and, to a lesser extent, other melanocortin receptors[231]. By inhibiting MC1R, ASIP reduces eumelanin production[232], which may largely explain its positive association with melanoma and other skin cancers. The oncogene ERBB3 was positively and causally associated with CMC in this study (Table S20, Fig. S16). ERBB3 plays a critical role in melanoma metastasis and is frequently hyperactivated in human melanomas, where its activity correlates with disease progression and poor survival[233,234]. Inhibition of ERBB3 has been shown to suppress metastatic potential[233]. Overall, these observations support a potential positive causal role of circulating ERBB3, reflecting tissue biology in CMC development. The serum protein ARG1 showed a positive and potentially causal association with CMC (Table S20, Fig. S16). ARG1 is highly expressed by tumor-associated macrophages in melanoma, where it promotes local immunosuppression and facilitates tumor progression[235,236]. Specifically, it inhibits T-cell infiltration[237], and genetic deletion of Arg1 in mouse myeloid lineages remodels the TME[237,238]. These observations are consistent with our findings.

**RNASET2 and IRF3** - Ribonuclease T2 (RNASET2) showed a positive association with CMC in the MR analysis (Table S20, Fig. S16). RNASET2 encodes a secreted ribonuclease known as a broad tumor suppressor, particularly in ovarian cancer[239]. Its expression is frequently reduced in ovarian tumors[239,240], whereas overexpression can inhibit tumor growth and metastasis[239,240]. Mechanistically, RNASET2 suppresses tumor progression and angiogenesis in part through non-cell-autonomous effects on the TME, including the recruitment and activation of M1-like (pro-inflammatory) macrophages that limit tumor growth[241]. Despite its established tumor-suppressive role in experimental models, our MR analysis revealed a positive causal association between circulating RNASET2 and melanoma risk. This apparent inconsistency may reflect context-dependent effects, whereby intracellular or locally active RNASET2 restrains tumor growth and supports antitumor immunity, while elevated serum RNASET2 represents a systemic response to stress or inflammation that precedes melanoma development. Interferon regulatory factor 3 (IRF3) showed a negative causal association with CMC in the present study (Table S20, Fig. S16). Higher IRF3 expression has been linked to improved survival in colorectal, liver, and lung cancers, likely through negative regulation of Wnt signaling[242,243]. Although IRF3 supports antitumor immunity, it has not been specifically implicated in melanoma. Our findings are therefore consistent with a potentially protective, causal role of IRF3 in CMC.

*STRING-defined clustering of proteins with putative causal associations*: STRING-based analysis[244] of all proteins with causal associations to cancers at FDR < 0.05 revealed significant enrichment of both functional and physical protein–protein interactions (115 observed vs. 53 expected edges; $P = 9 \times 10^{-14}$; Fig. S17). Our STRING + k-means clustering revealed pathway enrichments that map onto biologically coherent cancer processes including (Fig. S17): 1) The Laminins, nidogen (G2) domains and fibulins. These proteins are components of the extracellular matrix (ECM) and basement membrane that regulate cell adhesion, migration, invasion and the TME; remodeling of these molecules drives metastasis and alters stromal/immune interactions[245]. 2) Constitutive PI3K signaling and VEGF ligand–receptor interactions. Chronic activation of PI3K/AKT promotes proliferation, survival and HIF-driven VEGF expression; VEGF receptor signaling in endothelial and tumor cells drives angiogenesis and cooperates with PI3K to support tumor growth[246,247]. 3) Cytokine receptor activity. Cytokine receptors mediate paracrine/autocrine immune and inflammatory signaling in the TME and can promote tumor progression, immune suppression or therapeutic resistance depending on context[248]. 4) Ligand–receptor interactions (intercellular signaling). Cell-cell communication via ligand-receptor pairs (growth factors, chemokines, ECM receptors) structures tumor–stroma crosstalk and coordinates programs such as invasion, immune recruitment and angiogenesis[249]. 5) Glutathione metabolism. Glutathione metabolism buffers oxidative stress, contributes to redox control and drug detoxification, and is frequently upregulated in tumors to support proliferation and chemoresistance[250]. 6) RNA secondary-structure unwinding (RNA helicases). DEAD/H-box RNA helicases and related unwinders regulate mRNA translation, ribosome assembly and stress-responsive gene expression; dysregulation can promote oncogenic translation programs and affect immune sensing[251]. These pathways may interact as follows (integrative interpretation: For example, the ECM-PI3K/VEGF axis: ECM components (laminins, fibulins, nidogen) modulate integrin and growth factor receptor signaling, which feeds into PI3K/AKT activation. In turn, PI3K activation in tumor cells elevates VEGF expression, promoting angiogenesis and microenvironment remodeling, potentially creating a feed-forward loop connecting the two clusters[252]. In fact, these clusters exhibit interactions as revealed by the STRING-based analysis (Fig. S17). The cytokines and ligand-receptor signaling shape immune/stromal responses: Cytokine receptors and other ligand-receptor pairs coordinate recruitment/activation of immune and stromal cells that both respond to and remodel the ECM, affecting invasion and therapy response[248]. Redox and proteostasis support PI3K and other signaling networks: Elevated glutathione protects tumor cells from oxidative damage produced by rapid growth and immune attack, stabilizing signaling proteins and enabling sustained PI3K/VEGF and cytokine signaling. Meanwhile, RNA helicases enable translation of stress- and growth-promoting mRNAs, reinforcing oncogenic programs[253]. This integrative view highlights specific cross-pathway nodes such as ECM-regulated PI3K/VEGF signaling and stress-adaptation hubs that can serve as strategic entry points for follow-up studies aimed at experimentally dissecting causal mechanisms and identifying actionable biomarkers or intervention targets.

**Reference tables and figures**:

- Table S19 (excel): The GWAS studies providing summary statistics data for the MR analyses across various cancer types
- Table S20 (excel): MR results and integration with observational and proteogenomic findings for circulating proteins across cancer types
- Fig. 7 (in the main text): Forward, proteome-wide two-sample MR analysis linking the circulating proteome to cancer.
- Fig. S13: Forest plot of MR results for colorectal (CRC) at FDR < 0.05.
- Fig. S14: MR analysis results for associations between proteins and stomach cancer (STC) at FDR < 0.05.
- Fig. S15: Forest plot depicting MR results for breast cancer (BRC) at FDR < 0.05.
- Fig. S16: Forest plot of MR results for cutaneous melanoma cancer (CMC) at FDR < 0.05.
- Fig. S17: Protein–protein interaction networks among serum proteins with evidence of a causal relationship to cancer at FDR < 0.05.

**Table S1**. Number of cases for the 13 different types of cancer, including current (prevalent) and future (incident) diagnoses, with measurements of 7,523 serum proteins.

| Shared body system for cancer types | Cancer type | Variable* | Number of cancer cases (females), and follow-up | Number of those free of specific cancer type, and follow-up |
|---|---|---|---|---|
| ***Cancers of the digestive system*** | **Esophagus** | Prevalent | 3 (2) | 5320 |
| | | Incident | 16 (3) | 5307 |
| | | Follow-up period (years) | 6.0 [2.8, 8.0] | 10.0 [5.3, 11.4] |
| | **Stomach** | Prevalent | 25 (12) | 5298 |
| | | Incident | 26 (11) | 5297 |
| | | Follow-up period (years) | 5.7 [2.0, 8.9 | 10.6 [6.9, 11.7] |
| | **Colon** | Prevalent | 55 (28) | 5268 |
| | | Incident | 128 (75) | 5195 |
| | | Follow-up period (years) | 5.0 [2.7, 8.1] | 10.6 [7.2, 11.8] |
| | **Rectum** | Prevalent | 11 (3) | 5312 |
| | | Incident | 20 (8) | 5303 |
| | | Follow-up period (years) | 4.3 [1.5, 9.5] | 10.0 [5.3, 11.4] |
| | **Pancreas** | Prevalent | 1 (0) | 5322 |
| | | Incident | 38 (25) | 5285 |
| | | Follow-up period (years) | 6.0 [4.1, 9.5] | 10.6 [7.2, 11.6] |
| ***Cancers of the genitourinary system*** | **Kidney** | Prevalent | 32 (11) | 5291 |
| | | Incident | 49 (18) | 5274 |
| | | Follow-up period (years) | 5.4 [2.2, 7.4] | 10.6 [7.1, 11.8] |
| | **Prostate** | Prevalent | 107 (0) | 5216 |
| | | Incident | 192 (0) | 5131 |
| | | Follow-up period (years) | 4.9 [2.0, 7.0] | 10.6 [7.1, 11.6] |
| | **Bladder** | Prevalent | 71 (21) | 5252 |
| | | Incident | 77 (17) | 5246 |
| | | Follow-up period (years) | 4.9 [1.8, 8.0] | 10.6 [7.1, 11.7] |
| ***Cancers of the respiratory system*** | **Lung and bronchus** | Prevalent | 20 (11) | 5303 |
| | | Incident | 156 (86) | 5167 |
| | | Follow-up period (years) | 5.2 [2.6, 7.8] | 10.5 [6.9, 11.7] |
| ***Cancers of the female reproductive system*** | **Breast** | Prevalent | 124 (122) | 5199 |
| | | Incident | 107 (103) | 5216 |
| | | Follow-up period (years) | 5.1 [2.9, 7.8] | 10.6 [7.1, 11.7] |
| | **Corpus uteri** | Prevalent | 29 (0) | 5294 |
| | | Incident | 14 (0) | 5309 |
| | | Follow-up period (years) | 3.8 [2.6, 4.8] | 10.0 [5.3, 11.4] |
| | **Ovary** | Prevalent | 6 (0) | 5317 |
| | | Incident | 16 (0) | 5307 |
| | | Follow-up period (years) | 3.5 [1.5, 5.5] | 10.0 [5.3, 11.4] |
| ***Cancers of the skin*** | **Melanoma** | Prevalent | 14 (8) | 5309 |
| | | Incident | 79 (42) | 5244 |
| | | Follow-up period (years) | 7.0 [4.6, 9.2] | 10.1 [5.3, 11.5] |

*The follow-up period is the median [IQR; interquartile range] time until the date of specific cancer diagnosis, death, or end of follow-up for all cancer types, except for cancers of the esophagus, rectum, melanoma, ovary and corpus uteri, where follow-up time for any cancer diagnosis was used.

**Table S2**. Baseline characteristics of the AGES study participants, stratified by all 13 combined cancer types. All participants were measured for 7,523 proteins in serum.

| Characteristic | Variable* | Free of cancer | All 13 cancers combined | P-value | Total |
|---|---|---|---|---|---|
| ***Demographics*** | | | | | |
| | Numbers | 4088 (76.8) | 1235 (23.2) | N/A | 5323 |
| | AGE (years) | 76.6 (5.6) | 76.7 (5.5) | 0.354 | 76.6 (5.6) |
| | Sex (females) | 2445 (59.8) | 597 (48.3) | <0.001 | 3042 (57.1) |
| ***Anthropometry*** | | | | | |
| | BMI (kg/m$^2$) | 27.1 (4.4) | 26.9 (4.4) | 0.228 | 27.0 (4.4) |
| | BMI category | | | 0.390 | |
| | BMI<25 kg/m$^2$ | 1346 (33.0) | 431 (35.0) | | 1777 (33.4) |
| | BMI =20-30 kg/m$^2$ | 1823 (44.6) | 539 (43.8) | | 2362 (44.4) |
| | BMI ≥ 30 kg/m$^2$ | 915 (21.3) | 262 (21.3) | | 1177 (22.2) |
| ***Lifestyle*** | | | | | |
| | Smoking status | | | <0.001 | |
| | Never | 1770 (44.5) | 430 (35.8) | | 2200 (41.3) |
| | Former | 1752 (44.0) | 596 (49.6) | | 2348 (44.1) |
| | Current | 459 (11.5) | 176 (14.6) | | 635 (11.9) |
| | Alcohol use | 14.3 (31.0) | 16.1 (33.7) | 0.088 | 14.7 (32.0) |
| ***Physiological*** | | | | | |
| | eGFR (ml/min/1.73m$^2$) | 64.2 (15.4) | 63.6 (15.5) | 0.275 | 64.0 (15.4) |
| | Systolic blood pressure (mmHg) | 142.5 (20.5) | 142.9 (21.1) | 0.567 | 142.6 (20.7) |
| | Diastolic blood pressure (mmHg) | 73.9 (9.7) | 73.9 (9.9) | 0.989 | 73.9 (9.7) |
| ***Other*** | | | | | |
| | Follow-up period (any cancer) | 10.6 [7.0, 11.7] | 5.3 [2.3, 8.5] | <0.001 | 10.0 [5.3, 11.4] |

*Numbers are mean (SD) for continuous-, N (%) for categorical- and median [IQR] for skewed variables. The reported P-values are two-sided. Abbreviations: BMI, body mass index; eGFR, estimated glomerular filtration rate. N/A, not applicable.
Regular ANOVA was applied for continuous variables, while Chi-square test was used for categorical variables. For continuous variables with non-normal distributions, Kruskal-Wallis test was employed.

**Table S3**. Baseline characteristics of the AGES study participants as stratified by all digestive system-related cancers combined. All participants were measured for 7,523 proteins in serum.

| Characteristic | Variable* | Free of digestive system cancers | Cancers of the digestive system | P-value | Total |
|---|---|---|---|---|---|
| ***Demographics*** | | | | | |
| | Numbers | 5005 (94.0) | 318 (6.0) | N/A | 5323 |
| | AGE (years) | 76.6 (5.6) | 77.3 (5.2) | 0.035 | 76.6 (5.6) |
| | Sex (females) | 2876 (57.5) | 166 (52.2) | 0.075 | 3042 (57.1) |
| ***Anthropometry*** | | | | | |
| | BMI ($kg/m^2$) | 27.1 (4.4) | 26.2 (4.3) | 0.001 | 27.0 (4.4) |
| | BMI category | | | 0.022 | |
| | BMI<25 $kg/m^2$ | 1647 (32.9) | 130 (41.0) | | 1777 (33.4) |
| | BMI =20-30 $kg/m^2$ | 2222 (44.4) | 140 (44.2) | | 2362 (44.4) |
| | BMI ≥ 30 $kg/m^2$ | 1130 (22.6) | 47 (14.8) | | 1177 (22.2) |
| ***Lifestyle*** | | | | | |
| | Smoking status | | | 0.047 | |
| | Never | 2086 (42.8) | 114 (37.1) | | 2200 (41.3) |
| | Former | 2188 (44.9) | 160 (52.1) | | 2348 (44.1) |
| | Current | 602 (12.3) | 33 (10.7) | | 635 (11.9) |
| | Alcohol use | 14.7 (31.7) | 14.9 (31.4) | 0.887 | 14.7 (32.0) |
| ***Physiological*** | | | | | |
| | eGFR ($ml/min/1.73m^2$) | 64.0 (15.5) | 64.4 (15.2) | 0.629 | 64.0 (15.4) |
| | Systolic blood pressure (mmHg) | 142.6 (20.6) | 143.1 (21.1) | 0.671 | 142.6 (20.7) |
| | Diastolic blood pressure (mmHg) | 73.9 (9.7) | 73.8 (9.8) | 0.769 | 73.9 (9.7) |
| ***Other*** | | | | | |
| | Follow-up period (any cancer) | 10.2 [5.6, 11.6] | 5.4 [2.4, 9.2] | <0.001 | 10.0 [5.3, 11.4] |

*Numbers are mean (SD) for continuous-, N (%) for categorical- and median [IQR] for skewed variables. The reported P-values are two-sided. Abbreviations: BMI, body mass index; eGFR, estimated glomerular filtration rate. N/A, not applicable.
Regular ANOVA was applied for continuous variables, while Chi-square test was used for categorical variables. For continuous variables with non-normal distributions, Kruskal-Wallis test was employed.

**Table S4**. Baseline characteristics of the AGES study participants stratified for all combined genitourinary system-related cancers. All participants were measured for 7,523 proteins in serum.

| Characteristic | Variable* | Free of genitourinary cancer | Cancers of the genitourinary system | P-value | Total |
|---|---|---|---|---|---|
| ***Demographics*** | | | | | |
| | Numbers | 4838 (90.9) | 485 (9.1) | N/A | 5323 |
| | AGE (years) | 76.5 (5.6) | 77.3 (5.2) | 0.002 | 76.6 (5.6) |
| | Sex (females) | 2977 (61.5) | 65 (13.4) | <0.001 | 3042 (57.1) |
| ***Anthropometry*** | | | | | |
| | BMI (kg/m$^2$) | 27.0 (4.5) | 27.1 (3.8) | 0.975 | 27.0 (4.4) |
| | BMI category | | | 0.115 | |
| | BMI<25 kg/m$^2$ | 1631 (33.7) | 146 (30.2) | | 1777 (33.4) |
| | BMI =20-30 kg/m$^2$ | 2126 (44.0) | 236 (48.9) | | 2362 (44.4) |
| | BMI ≥ 30 kg/m$^2$ | 1076 (22.3) | 101 (20.9) | | 1177 (22.2) |
| ***Lifestyle*** | | | | | |
| | Smoking status | | | <0.001 | |
| | Never | 2059 (43.7) | 141 (29.8) | | 2200 (41.3) |
| | Former | 2070 (43.9) | 278 (58.8) | | 2348 (44.1) |
| | Current | 581 (12.3) | 64 (11.4) | | 635 (11.9) |
| | Alcohol use | 14.2 (30.9) | 19.5 (38.4) | <0.001 | 14.7 (32.0) |
| ***Physiological*** | | | | | |
| | eGFR (ml/min/1.73m$^2$) | 64.1 (15.4) | 62.9 (15.8) | 0.087 | 64.0 (15.4) |
| | Systolic blood pressure (mmHg) | 142.5 (20.7) | 143.0 (20.5) | 0.611 | 142.6 (20.7) |
| | Diastolic blood pressure (mmHg) | 73.8 (9.7) | 75.3 (9.8) | 0.002 | 73.9 (9.7) |
| ***Other*** | | | | | |
| | Follow-up (any cancer) | 10.3 [5.6, 11.6] | 4.6 [1.7, 7.4] | <0.001 | 10.0 [5.3, 11.4] |

*Numbers are mean (SD) for continuous-, N (%) for categorical- and median [IQR] for skewed variables. The reported P-values are two-sided. Abbreviations: BMI, body mass index; eGFR, estimated glomerular filtration rate. N/A, not applicable.
Regular ANOVA was applied for continuous variables, while Chi-square test was used for categorical variables. For continuous variables with non-normal distributions, Kruskal-Wallis test was employed.

**Table S5**. Baseline characteristics of the AGES study participants stratified for cancer of the lung and bronchus. All participants were measured for 7,523 proteins in serum.

| Characteristic | Variable* | Free of respiratory system cancers | Cancers of the respiratory system | P-value | Total |
|---|---|---|---|---|---|
| ***Demographics*** | | | | | |
| | Numbers | 5147 (96.7) | 176 (3.3) | N/A | 5323 |
| | AGE (years) | 76.7 (5.6) | 74.6 (4.7) | <0.001 | 76.6 (5.6) |
| | Sex (females) | 2945 (57.2) | 97 (55.1) | 0.663 | 3042 (57.1) |
| ***Anthropometry*** | | | | | |
| | BMI (kg/m$^2$) | 27.1 (4.4) | 26.3 (4.6) | 0.033 | 27.0 (4.4) |
| | BMI category | | | 0.146 | |
| | BMI<25 kg/m$^2$ | 1707 (33.2) | 70 (39.8) | | 1777 (33.4) |
| | BMI =20-30 kg/m$^2$ | 2295 (44.6) | 67 (38.1) | | 2362 (44.4) |
| | BMI ≥ 30 kg/m$^2$ | 1138 (22.1) | 39 (22.2) | | 1177 (22.2) |
| ***Lifestyle*** | | | | | |
| | Smoking status | | | <0.001 | |
| | Never | 2187 (43.6) | 13 (7.6) | | 2200 (41.3) |
| | Former | 2257 (45.0) | 91 (53.5) | | 2348 (44.1) |
| | Current | 569 (11.4) | 66 (38.8) | | 635 (11.9) |
| | Alcohol use | 14.5 (31.4) | 19.3 (38.1) | 0.052 | 14.7 (32.0) |
| ***Physiological*** | | | | | |
| | eGFR (ml/min/1.73m$^2$) | 63.9 (15.5) | 66.8 (14.6) | 0.015 | 64.0 (15.4) |
| | Systolic blood pressure (mmHg) | 142.7 (20.6) | 139.9 (22.0) | 0.077 | 142.6 (20.7) |
| | Diastolic blood pressure (mmHg) | 73.9 (9.7) | 73.2 (9.9) | 0.318 | 73.9 (9.7) |
| ***Other*** | | | | | |
| | Follow-up (any cancer) | 10.1 [5.5, 11.5] | 4.5 [1.3, 7.1] | <0.001 | 10.0 [5.3, 11.4] |

*Numbers are mean (SD) for continuous-, N (%) for categorical- and median [IQR] for skewed variables. The reported P-values are two-sided. Abbreviations: BMI, body mass index; eGFR, estimated glomerular filtration rate. N/A, not applicable.

Regular ANOVA was applied for continuous variables, while Chi-square test was used for categorical variables. For continuous variables with non-normal distributions, Kruskal-Wallis test was employed.

**Table S6**. Baseline characteristics of the AGES study participants stratified for all cancers of the female reproductive system. All participants were measured for 7,523 proteins in serum.

| **Characteristic** | **Variable*** | **Free of female reproductive system cancers** | **Cancers of the female reproductive system** | **P-value** | **Total** |
|---|---|---|---|---|---|
| ***Demographics*** | | | | | |
| | Numbers | 5036 (94.6) | 287 (5.4) | N/A | 5323 |
| | AGE (years) | 76.7 (5.6) | 75.9 (5.6) | 0.030 | 76.6 (5.6) |
| | Sex (females) | 2761 (54.8) | 281 (97.9) | <0.001 | 3042 (57.1) |
| ***Anthropometry*** | | | | | |
| | BMI (kg/m$^2$) | 27.0 (4.4) | 28.1 (5.1) | <0.001 | 27.0 (4.4) |
| | BMI category | | | 0.001 | |
| | BMI<25 kg/m$^2$ | 1693 (33.7) | 84 (29.3) | | 1777 (33.4) |
| | BMI =20-30 kg/m$^2$ | 2249 (44.7) | 113 (39.4) | | 2362 (44.4) |
| | BMI ≥ 30 kg/m$^2$ | 1087 (21.6) | 90 (31.4) | | 1177 (22.2) |
| ***Lifestyle*** | | | | | |
| | Smoking status | | | <0.001 | |
| | Never | 2047 (41.8) | 153 (54.4) | | 2200 (41.3) |
| | Former | 2255 (46.0) | 93 (33.1) | | 2348 (44.1) |
| | Current | 600 (12.2) | 35 (12.5) | | 635 (11.9) |
| | Alcohol use | 14.9 (31.9) | 11.4 (26.4) | 0.072 | 14.7 (32.0) |
| ***Physiological*** | | | | | |
| | eGFR (ml/min/1.73m$^2$) | 64.1 (15.4) | 62.3 (15.9) | 0.062 | 64.0 (15.4) |
| | Systolic blood pressure (mmHg) | 142.5 (20.6) | 144.2 (22.4) | 0.168 | 142.6 (20.7) |
| | Diastolic blood pressure (mmHg) | 74.0 (9.7) | 73.2 (9.9) | 0.181 | 73.9 (9.7) |
| ***Other*** | | | | | |
| | Follow-up (any cancer) | 10.1 [5.5, 11.5] | 6.0 [3.0, 10.1] | <0.001 | 10.0 [5.3, 11.4] |

*Numbers are mean (SD) for continuous-, N (%) for categorical- and median [IQR] for skewed variables. The reported P-values are two-sided. Abbreviations: BMI, body mass index; eGFR, estimated glomerular filtration rate. N/A, not applicable.

Regular ANOVA was applied for continuous variables, while Chi-square test was used for categorical variables. For continuous variables with non-normal distributions, Kruskal-Wallis test was employed.

**Table S7**. Baseline characteristics of the AGES study participants stratified for all skin cancers (melanoma). All participants were measured for 7,523 proteins in serum.

| Characteristic | Variable* | Free of melanoma | Melanoma | P-value | Total |
|---|---|---|---|---|---|
| ***Demographics*** | | | | | |
| | Numbers | 5230 (98.3) | 93 (1.7) | N/A | 5323 |
| | AGE (years) | 76.6 (5.6) | 77.5 (5.5) | 0.137 | 76.6 (5.6) |
| | Sex (females) | 2992 (57.2) | 50 (53.8) | 0.576 | 3042 (57.1) |
| ***Anthropometry*** | | | | | |
| | BMI ($kg/m^2$) | 27.1 (4.4) | 26.6 (3.4) | 0.308 | 27.0 (4.4) |
| | BMI category | | | 0.565 | |
| | BMI<25 $kg/m^2$ | 1742 (33.4) | 35 (37.6) | | 1777 (33.4) |
| | BMI =20-30 $kg/m^2$ | 2321 (44.4) | 41 (44.1) | | 2362 (44.4) |
| | BMI ≥ 30 $kg/m^2$ | 1160 (21.2) | 17 (18.3) | | 1177 (22.2) |
| ***Lifestyle*** | | | | | |
| | Smoking status | | | 0.569 | |
| | Never | 2166 (42.5) | 34 (37.4) | | 2200 (41.3) |
| | Former | 2302 (45.2) | 46 (50.5) | | 2348 (44.1) |
| | Current | 624 (12.3) | 11 (12.1) | | 635 (11.9) |
| | Alcohol use | 14.7 (31.7) | 14.9 (32.5) | 0.961 | 14.7 (32.0) |
| ***Physiological*** | | | | | |
| | eGFR ($ml/min/1.73m^2$) | 63.9 (15.4) | 64.9 (15.5) | 0.100 | 64.0 (15.4) |
| | Systolic blood pressure (mmHg) | 142.6 (20.7) | 144.0 (21.0) | 0.514 | 142.6 (20.7) |
| | Diastolic blood pressure (mmHg) | 73.9 (9.7) | 72.1 (10.5) | 0.064 | 73.9 (9.7) |
| ***Other*** | | | | | |
| | Follow-up (any cancer) | 10.1 [5.3, 11.5] | 7.0 [4.4, 9.2] | <0.001 | 10.0 [5.3, 11.4] |

*Numbers are mean (SD) for continuous-, N (%) for categorical- and median [IQR] for skewed variables. The reported P-values are two-sided. Abbreviations: BMI, body mass index; eGFR, estimated glomerular filtration rate. N/A, not applicable.

Regular ANOVA was applied for continuous variables, while Chi-square test was used for categorical variables. For continuous variables with non-normal distributions, Kruskal-Wallis test was employed.

**Table S9**. Baseline characteristics of AGES study participants, stratified by an additional 684 patients in the "any cancer" group. All participants were measured for 7,523 proteins in serum.

| Characteristic | Variable* | Without cancers diagnosed in the 684-patient group | The 684-patient group | P-value | Total |
|---|---|---|---|---|---|
| ***Demographics*** | | | | | |
| | Numbers | 4639 (87.2) | 684 (12.8) | N/A | 5323 |
| | AGE (years) | 76.6 (5.6) | 76.4 (5.4) | 0.262 | 76.6 (5.6) |
| | Sex (females) | 2604 (56.1) | 438 (64.0) | <0.001 | 3042 (57.1) |
| ***Anthropometry*** | | | | | |
| | BMI (kg/m$^2$) | 27.0 (4.4) | 27.0 (4.5) | 0.983 | 27.0 (4.4) |
| | BMI category | | | 0.935 | |
| | BMI<25 kg/m$^2$ | 1551 (33.5) | 226 (33.0) | | 1777 (33.4) |
| | BMI =20-30 kg/m$^2$ | 2059 (44.5) | 303 (44.3) | | 2362 (44.4) |
| | BMI ≥ 30 kg/m$^2$ | 1022 (22.1) | 155 (22.7) | | 1177 (22.1) |
| ***Lifestyle*** | | | | | |
| | Smoking status | | | 0.521 | |
| | Never | 1926 (42.6) | 274 (41.1) | | 2200 (41.3) |
| | Former | 2045 (45.3) | 303 (45.4) | | 2348 (44.1) |
| | Current | 545 (12.1) | 90 (13.5) | | 635 (11.9) |
| | Alcohol use | 14.9 (31.9) | 13.4 (30.4) | 0.251 | 14.7 (32.0) |
| ***Physiological*** | | | | | |
| | eGFR (ml/min/1.73m$^2$) | 64.0 (15.5) | 61.8 (14.5) | 0.175 | 64.0 (15.4) |
| | Systolic blood pressure (mmHg) | 142.5 (20.5) | 143.0 (21.5) | 0.578 | 142.6 (20.7) |
| | Diastolic blood pressure (mmHg) | 73.9 (9.7) | 74.1 (10.3) | 0.598 | 73.9 (9.7) |
| ***Other*** | | | | | |
| | Follow-up (any cancer) | 10.2 [5.6, 11.6] | 7.1 [3.4, 10.3] | <0.001 | 10.0 [5.3, 11.4] |

*Numbers are mean (SD) for continuous-, N (%) for categorical- and median [IQR] for skewed variables. The reported P-values are two-sided. Abbreviations: BMI, body mass index; eGFR, estimated glomerular filtration rate. N/A, not applicable.
Regular ANOVA was applied for continuous variables, while Chi-square test was used for categorical variables. For continuous variables with non-normal distributions, Kruskal-Wallis test was employed.

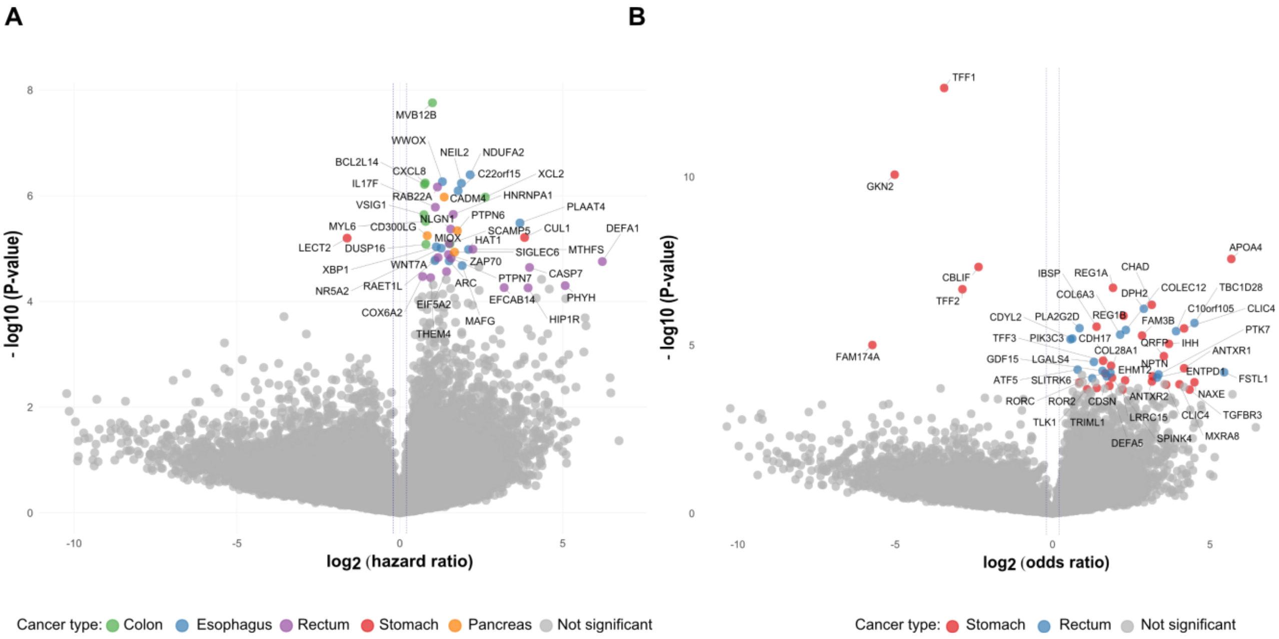


**Fig. S1**. A multi-cancer volcano plot illustrating serum proteins significantly associated with various digestive system cancers: (**A**) incident cancers of the colon (green circles), esophagus (blue circles), rectum (purple circles), stomach (red circles), and pancreas (orange circles); (**B**) prevalent cancers of the stomach (red circles) and rectum (blue circles). A selected subset of proteins is highlighted. The regression analysis incorporated standard covariate adjustments. Grey data points represent non-significant associations at FDR < 0.05. For further details on proteins linked to incident or prevalent cancers, see Table S10-10.

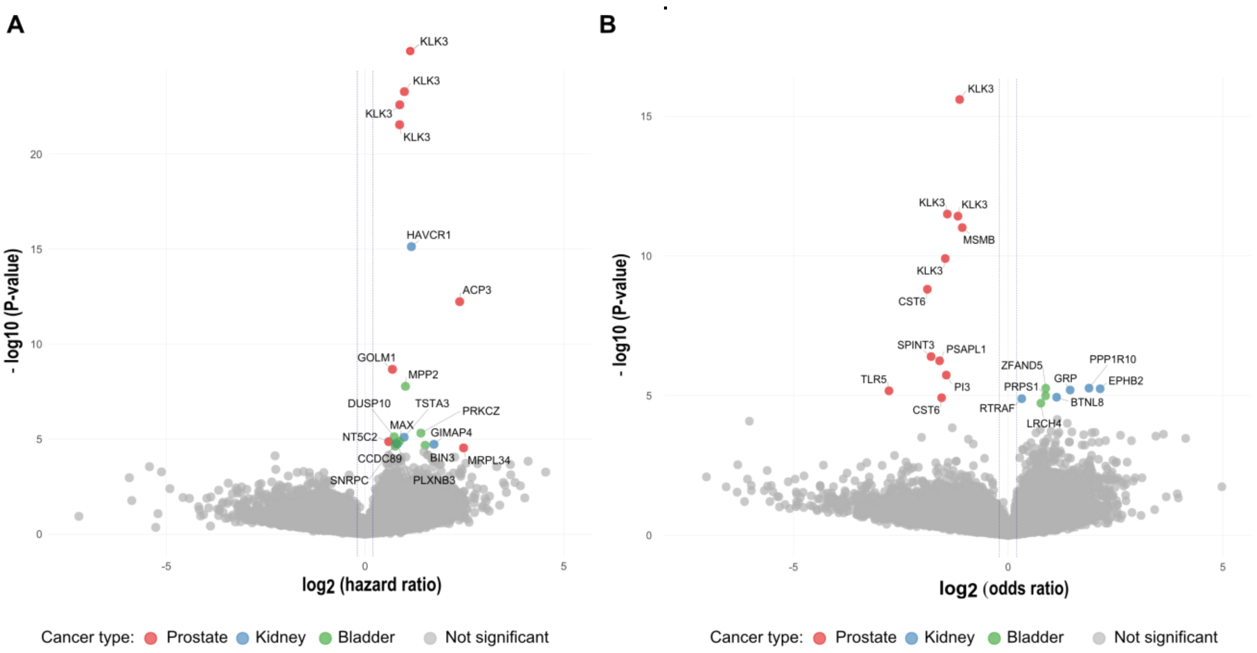


**Fig. S2**. A multicancer volcano plot shows serum proteins associated with cancers of the genitourinary system: (**A**) incident cancers of the prostate (red circles), kidney (blue circles), and bladder (green circles); (**B**) prevalent cancers of the same type as in (**A**). A selected set of proteins is highlighted. The regression analysis applied the standard covariate adjustment. Data points in grey indicate non-significant associations at FDR < 0.05. For additional information on proteins associated with incident or prevalent cancers, refer to Tables S10–S11.

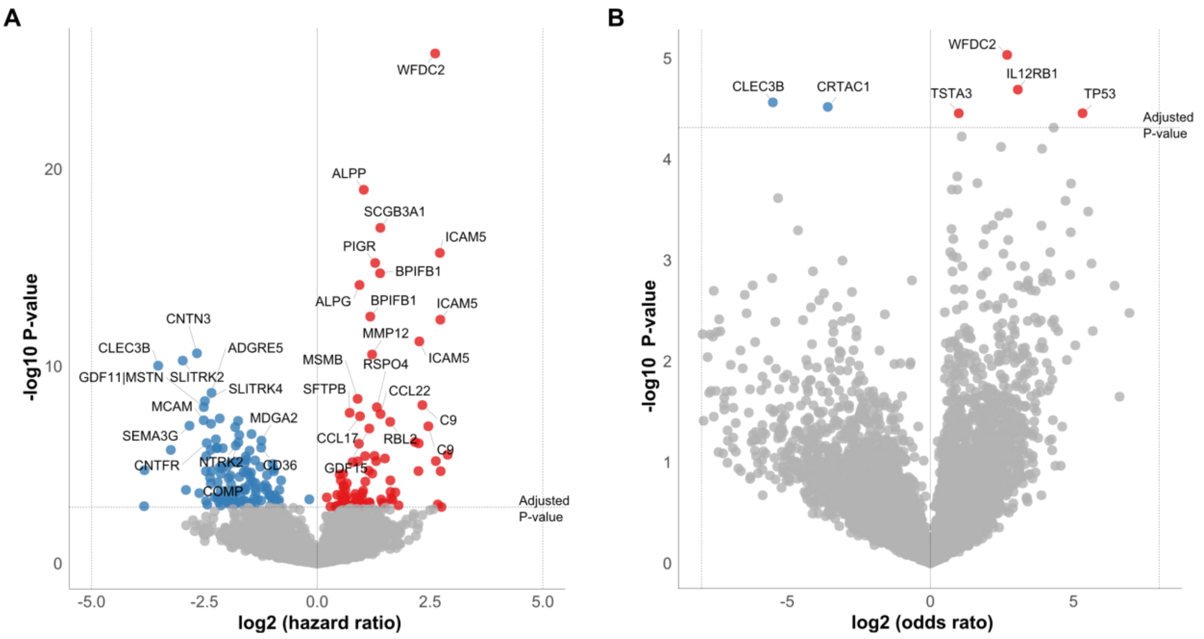


**Fig. S3**. The figure shows a volcano plot of proteins associated with (**A**) incident lung cancer (LUC) and (**B**) prevalent LUC. Proteins positively associated with ATC are shown in red, while those negatively associated are shown in blue. The plot highlights a selected set of proteins. The regression analysis incorporated the standard covariate adjustment. For additional information on proteins associated with incident or prevalent cancers, refer to Tables S10–S11.

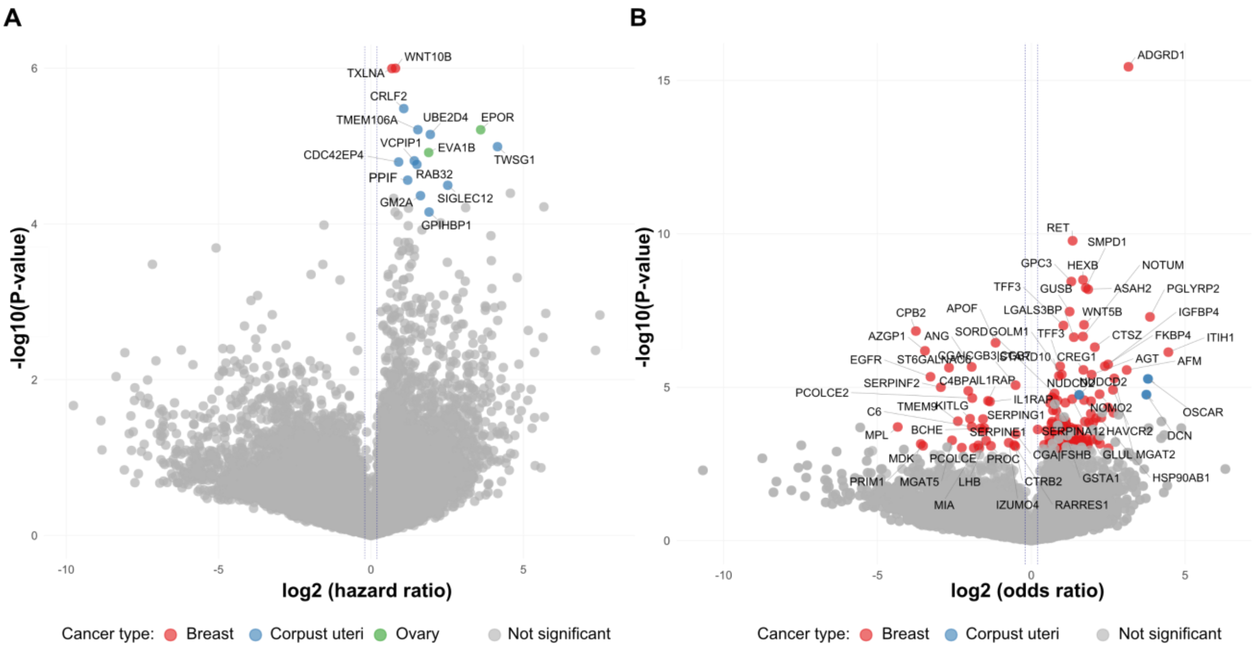


**Fig. S4**. A multicancer volcano plot displaying serum proteins significantly linked to various cancers of the female reproductive system: (**A**) incident cancers of the breast (red circles), corpus uteri (blue circles), and ovaries (green circles); (**B**) prevalent cancers of same type as in (**A**). A selected set of proteins is highlighted. The regression analysis applied the standard covariate adjustment. Data points in grey indicate non-significant associations at FDR < 0.05. For additional information on proteins associated with incident or prevalent cancers, refer to Tables S10–S11.

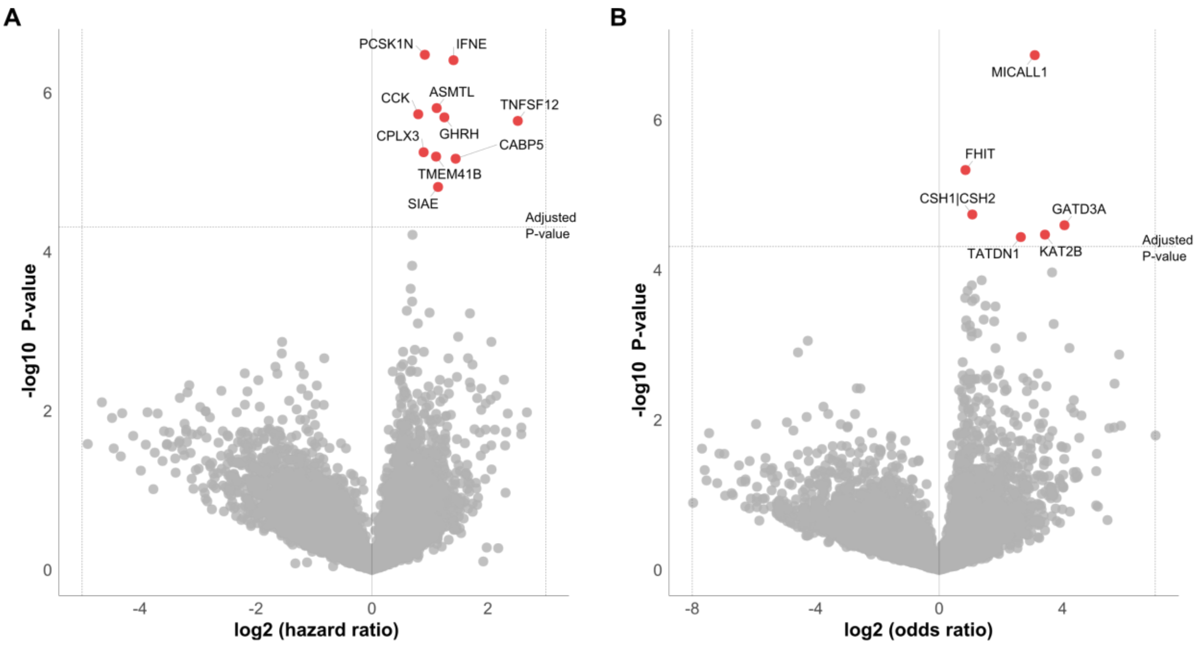


**Fig. S5**. The figure displays a volcano plot of proteins associated with (**A**) incident cutaneous melanoma (CMC) and (**B**) prevalent CMC. Proteins positively associated with ATC are shown in red, while those negatively associated are shown in blue. The plot highlights a selected set of proteins. The regression analysis incorporated the standard covariate adjustment. For additional information on proteins associated with incident or prevalent cancers, refer to Tables S10–S11.

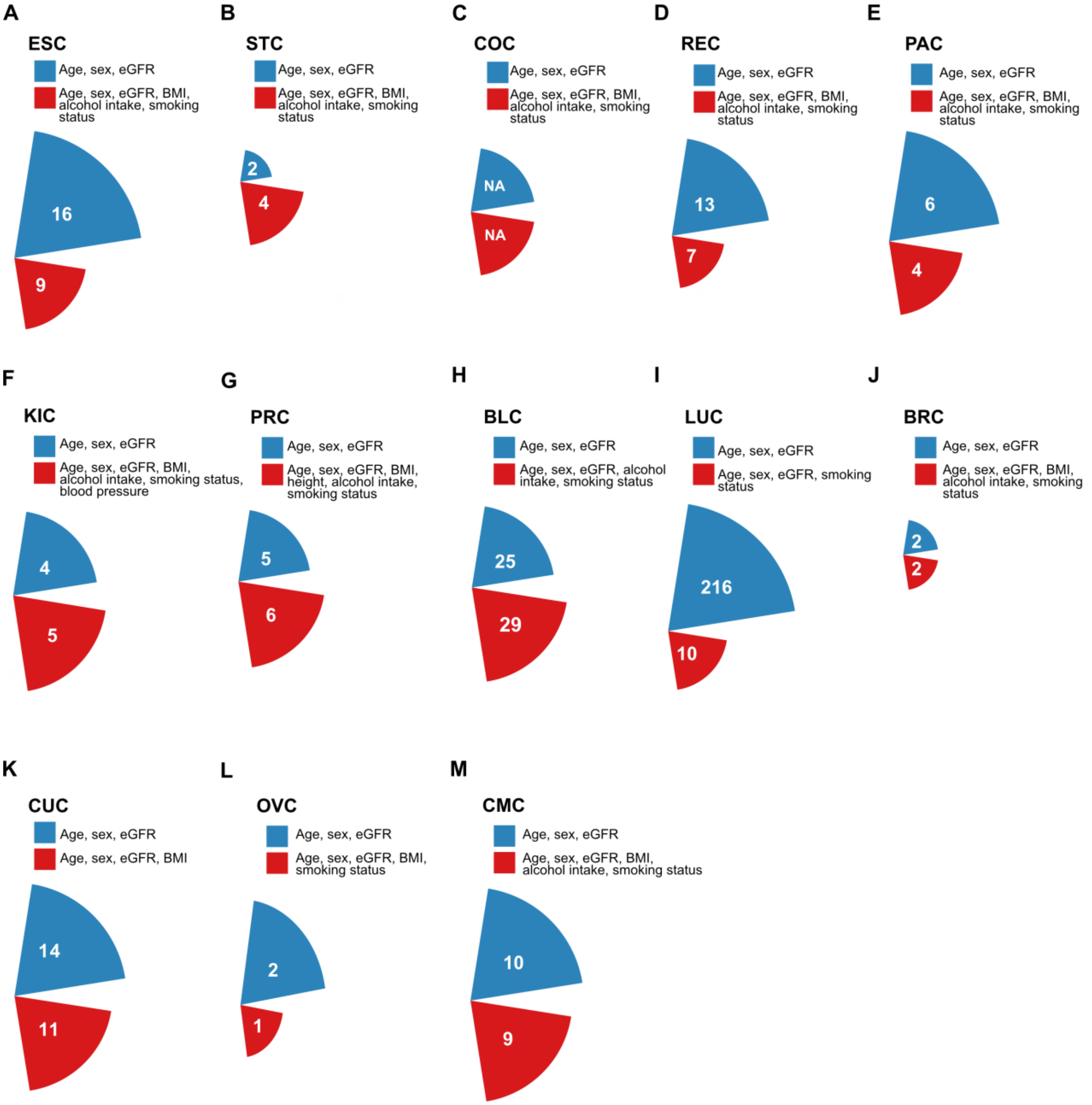


**Fig. S6A-M**. The number of serum proteins associated with incident cancers across various types before (adjusted for standard covariates; blue boxes and corresponding sectors) and after full adjustment (additionally adjusted for cancer type–specific epidemiological risk factors; red boxes and corresponding sectors). See Fig. 1 for cancer type abbreviations and the Supplementary Text for adjustment details. “NA” indicates no significant associations (incident COC).

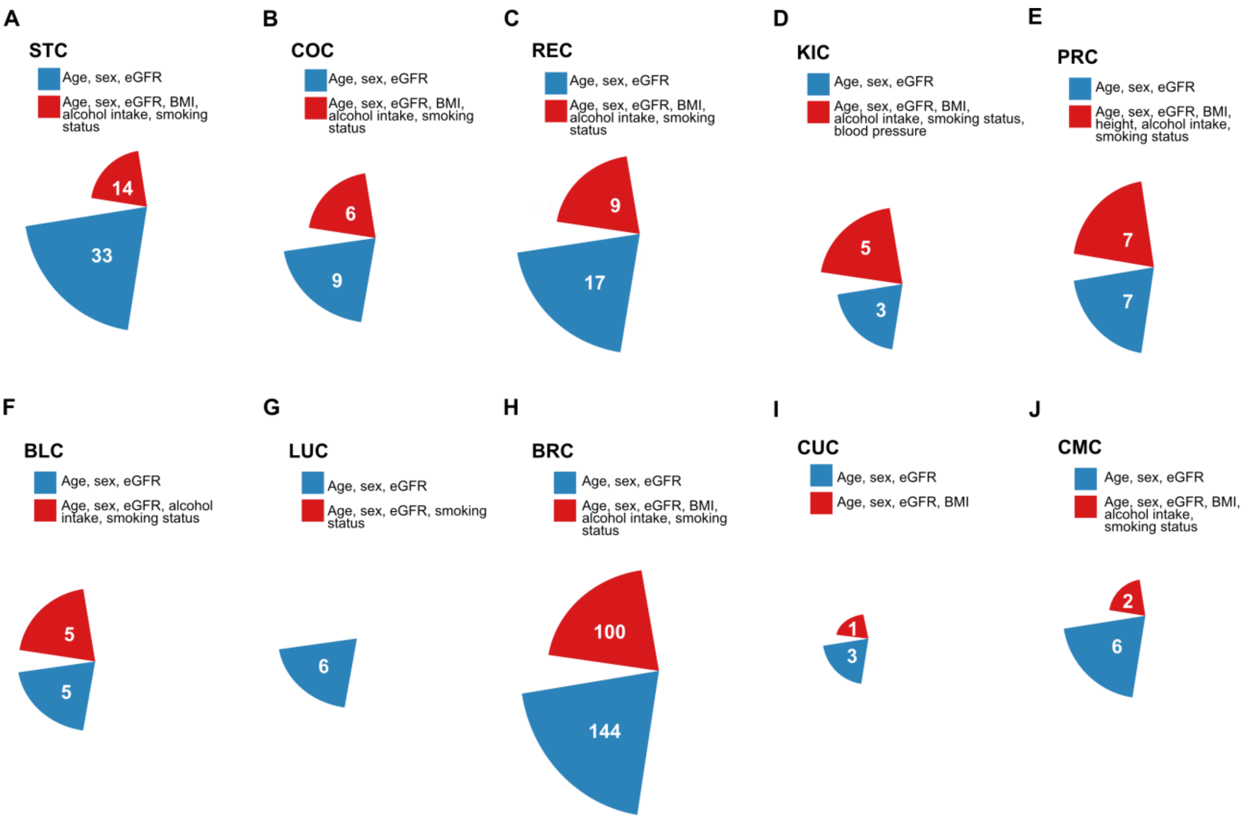


**Fig. S7A-M**. The number of serum proteins associated with different types of prevalent cancers before (adjusted for standard covariates; blue boxes and corresponding sectors) and after full adjustment (additionally adjusted for cancer type–specific epidemiological risk factors; red boxes and corresponding sectors). See Fig. 1 for cancer type abbreviations and the Supplementary Text for adjustment details.

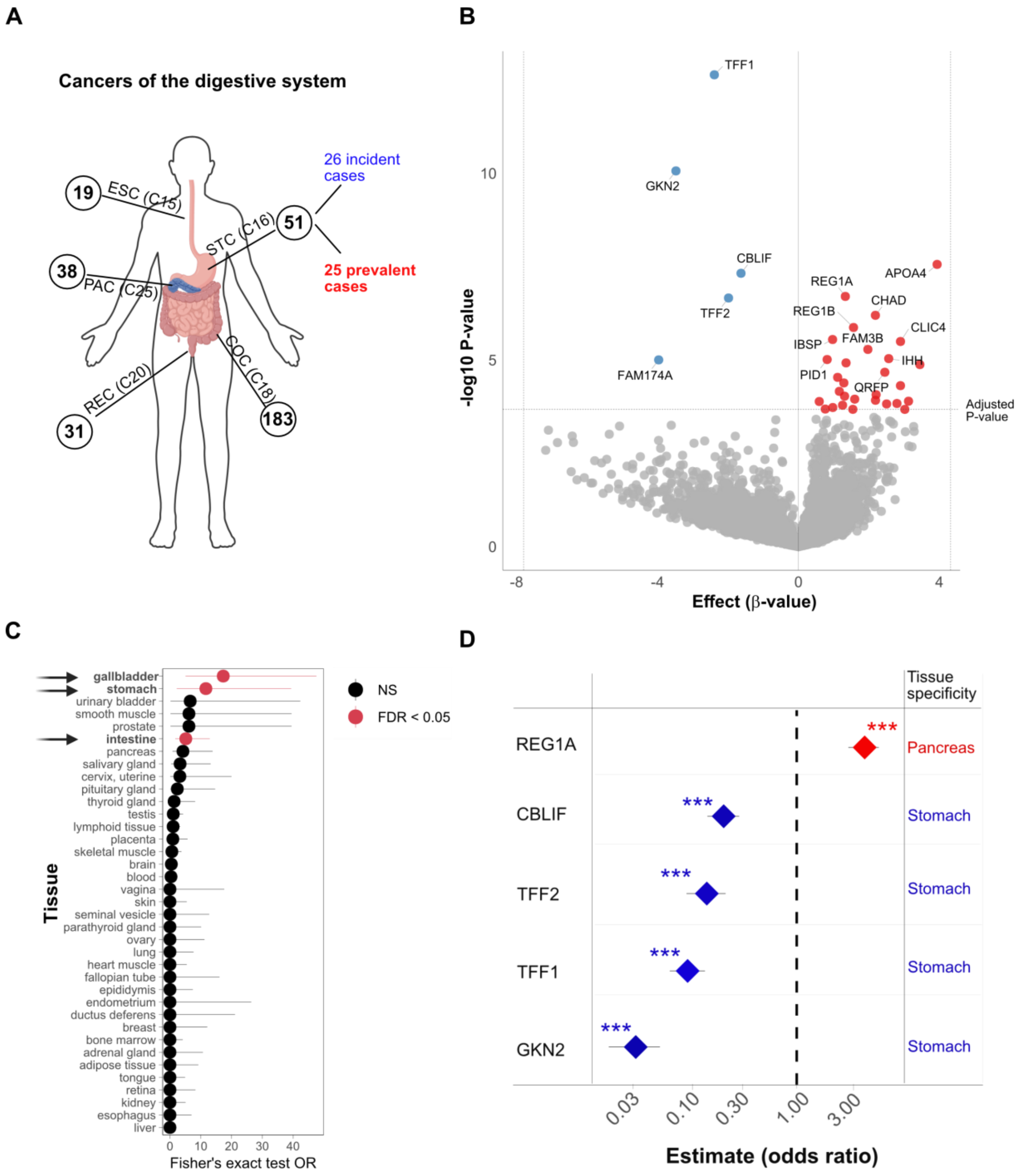


**Fig. S8**. Tissue specificity of serum proteins associated with prevalent stomach cancer: (**A**) Digestive system cancers examined in the present study. (**B**) Volcano plot depicting proteins significantly associated with prevalent stomach cancer (STC). (**C**) Tissue distribution of these proteins according to data from the Human Protein Atlas[11]. (**D**) Forest plot showing log2-transformed protein levels (odds ratios) with 95% confidence intervals (CIs) represented as horizontal lines, highlighting the tissue specificity of the selected subset of associated proteins. Statistical significance was determined using Benjamini-Hochberg correction with a false discovery rate (FDR) threshold of < 0.05, ***. Red data points in (**B**) and (**D**) represent significantly upregulated proteins, while blue data points indicate downregulated proteins.

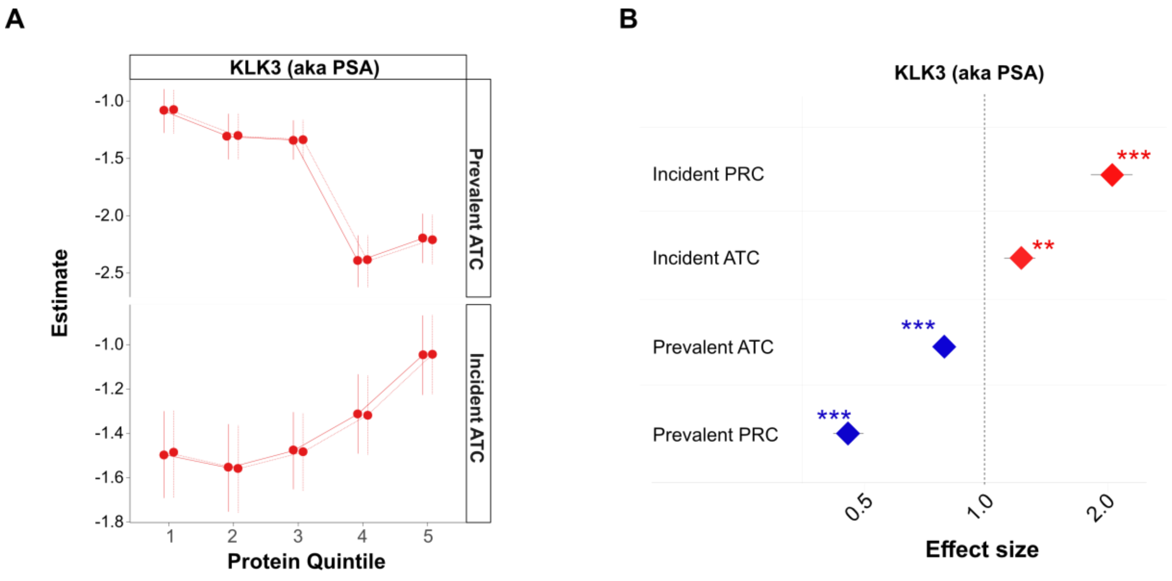


**Fig. S9**. (**A**) Quintile plot of KLK3 serum levels in relation to prevalent and incident cases of any cancer (ATC). The solid line represents adjustment for age and sex, while the dotted line additionally includes eGFR. (**B**) A forest plot depicting the association of KLK3 with ATC and prostate cancer (PRC), emphasizing differences in effect sizes between ATC and the individual analysis of PRC.

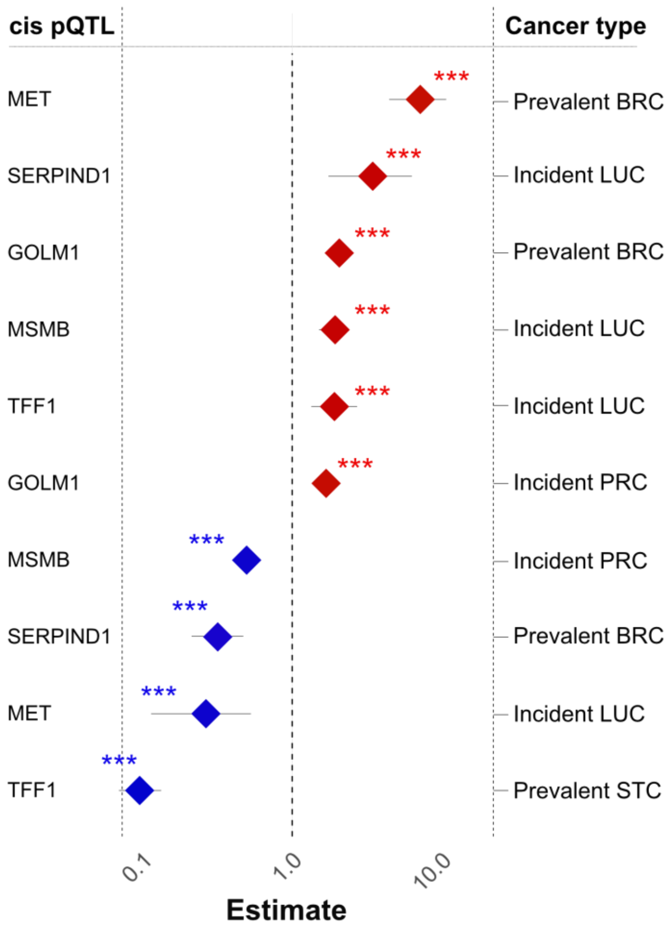


**Fig. S10**. The figure highlights the overlap of certain proteins among those linked to genetic cancer risk and incident and/or prevalent cancers in the observational analysis, specifically those that also have a *cis*-acting protein quantitative trait locus (pQTL) in the AGES study (see main text).

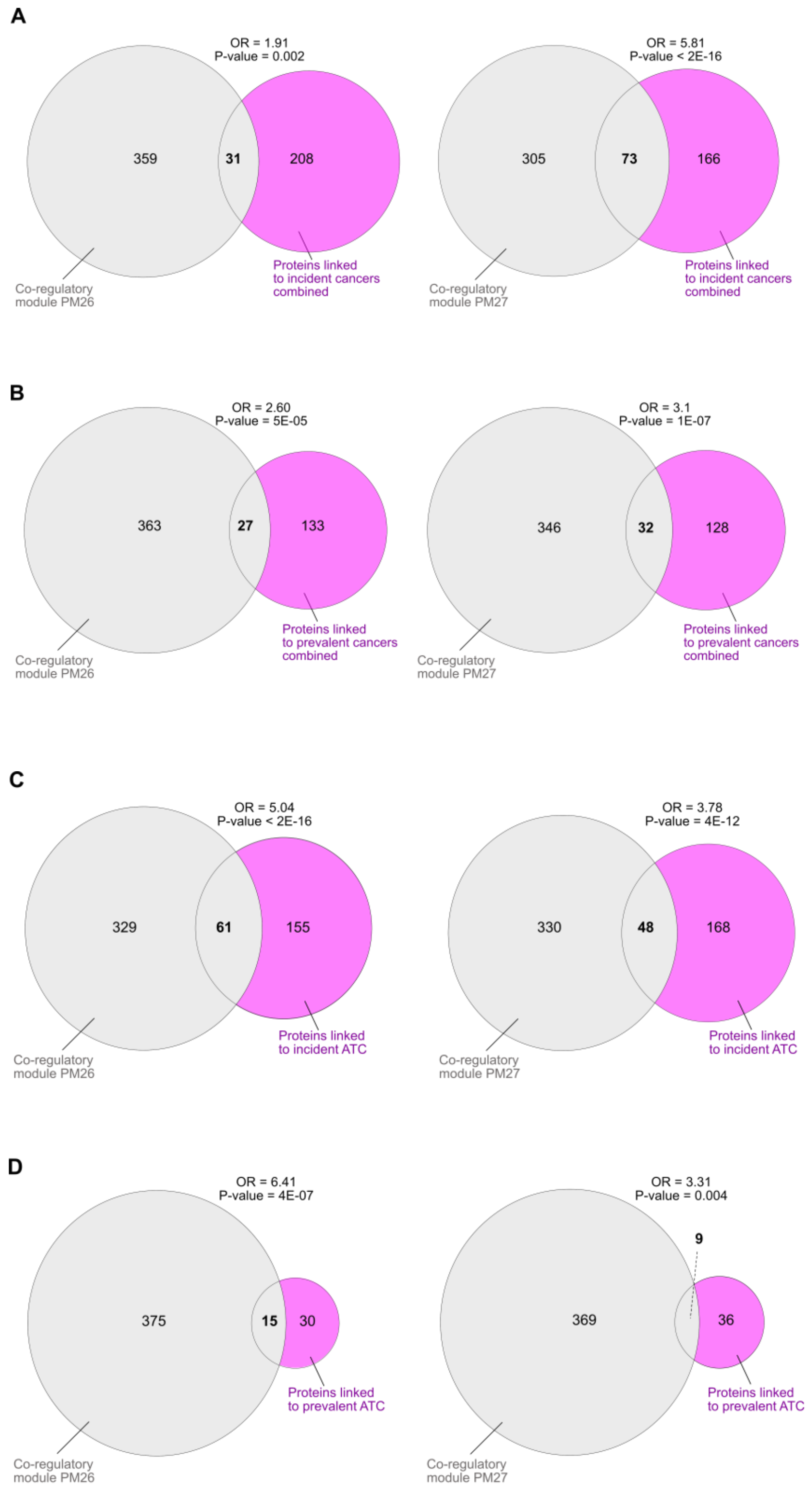


**Fig. S11**. Overlap between serum protein co-regulatory networks and proteins associated with (**A**) all incident cancers combined, (**B**) all prevalent cancers combined, (**C**) incident ATC, and (**D**) prevalent ATC.

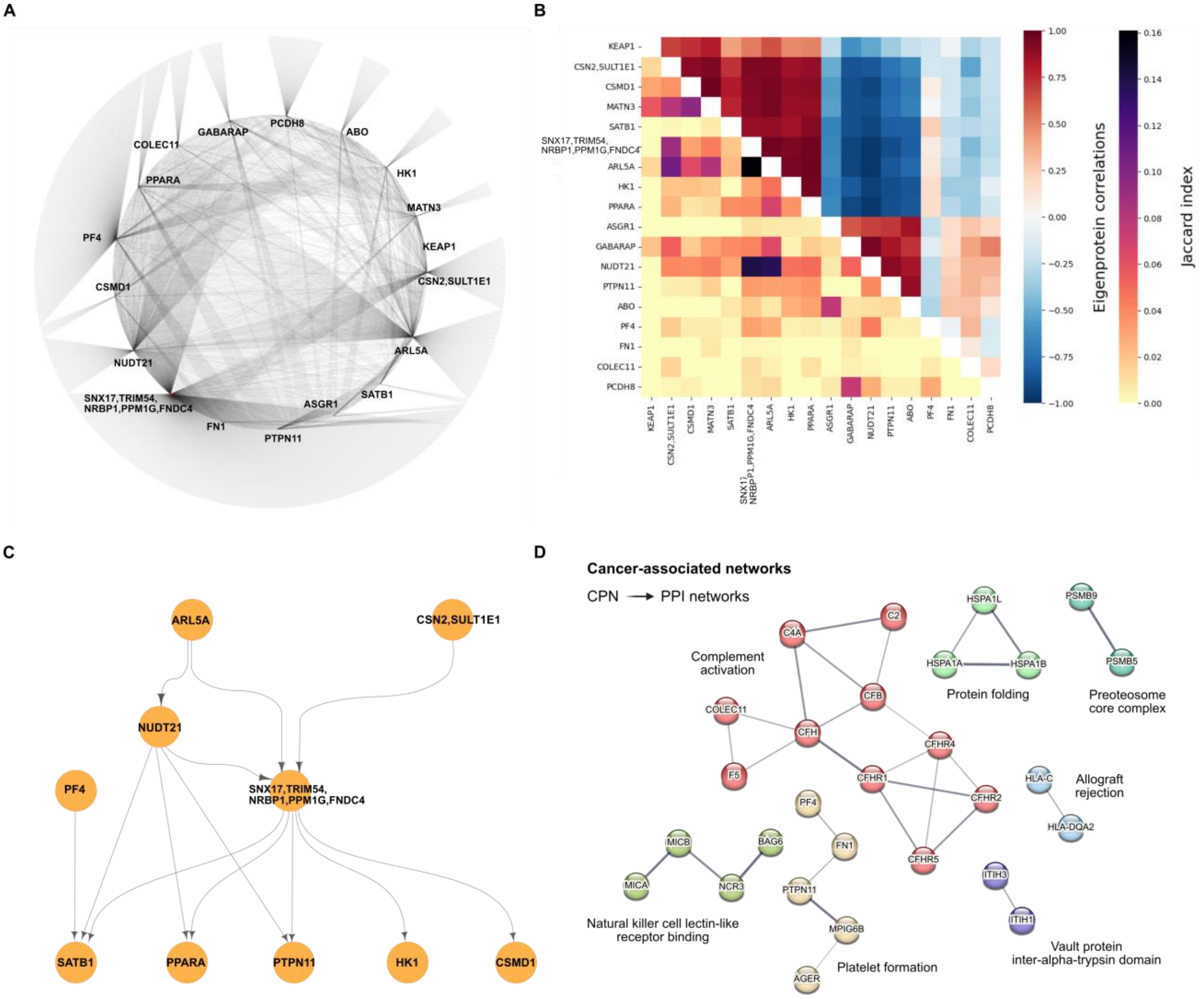


**Fig. S12**. (**A**) Network visualization showing the interconnections among circulating causal protein networks (CPNs) enriched for various incident cancer–associated protein signatures. (**B**) The heatmap illustrates the similarity between CPN subnetworks enriched with cancer-associated protein signatures. The upper right triangle presents hierarchical clustering based on pairwise Pearson correlations between network eigenproteins. The clustering leaf order is applied to the lower left triangle, which displays pairwise Jaccard Index scores between the same networks. (**C**) Hierarchical representation of interacting network regulators for CPNs enriched with cancer-associated proteins. (**D**) Functional and physical protein-protein interactions networks among regulators of the CPN subnetworks enriched for cancer-associated proteins across multiple cancer types, as identified by the STRING database. The edges represent functional and physical interactions, and unconnected network regulators are excluded from visualization.

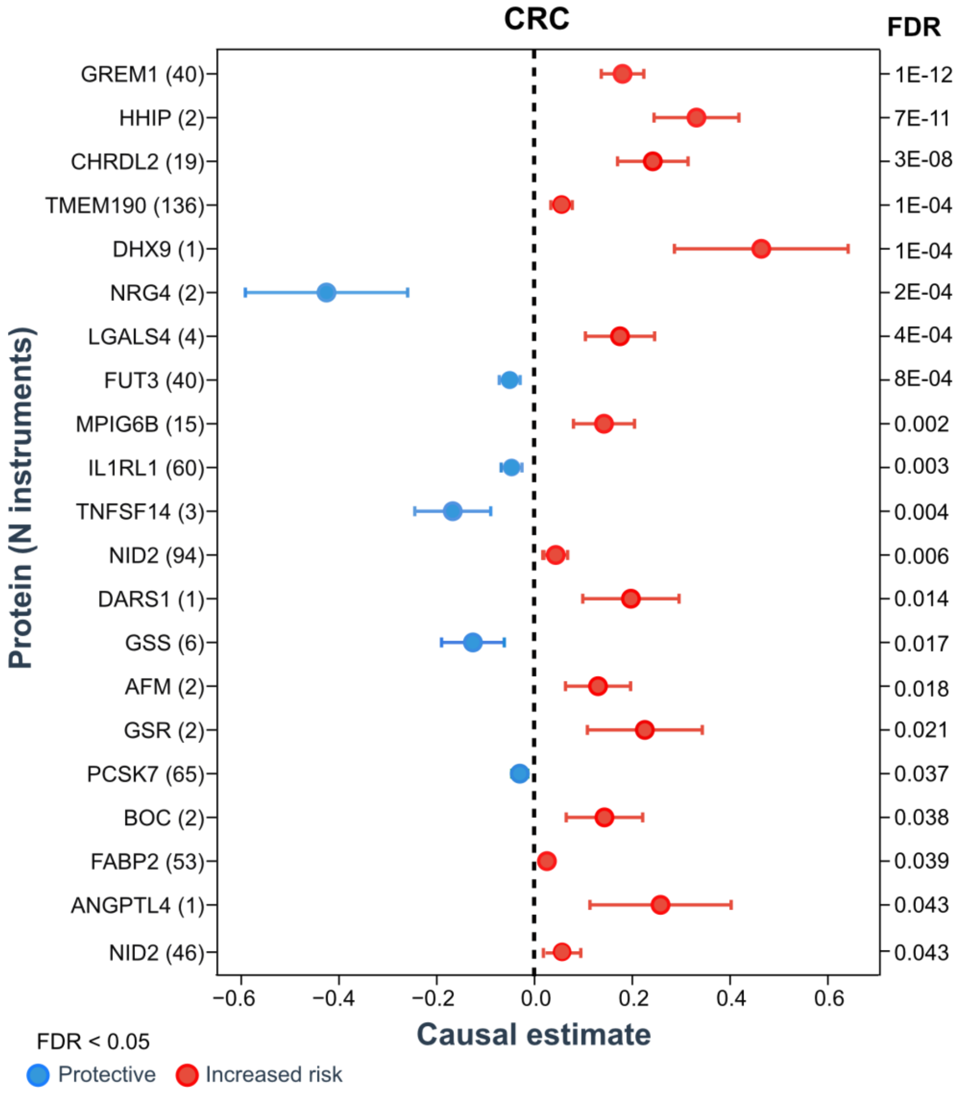


**Fig. S13**. Forest plot of MR results for colorectal (CRC) at FDR < 0.05 where proteins with the strongest associations appear toward the top. Red dots indicate positive causal associations (increasing risk), while blue dots denote negative associations (potentially protective) with the disease. The y-axis lists protein annotations with the number of instruments in parentheses. Repeated protein names indicate multiple aptamers with potential causal associations with CRC. The corresponding false discovery rate (FDR) for each test is shown to the right of the plot.

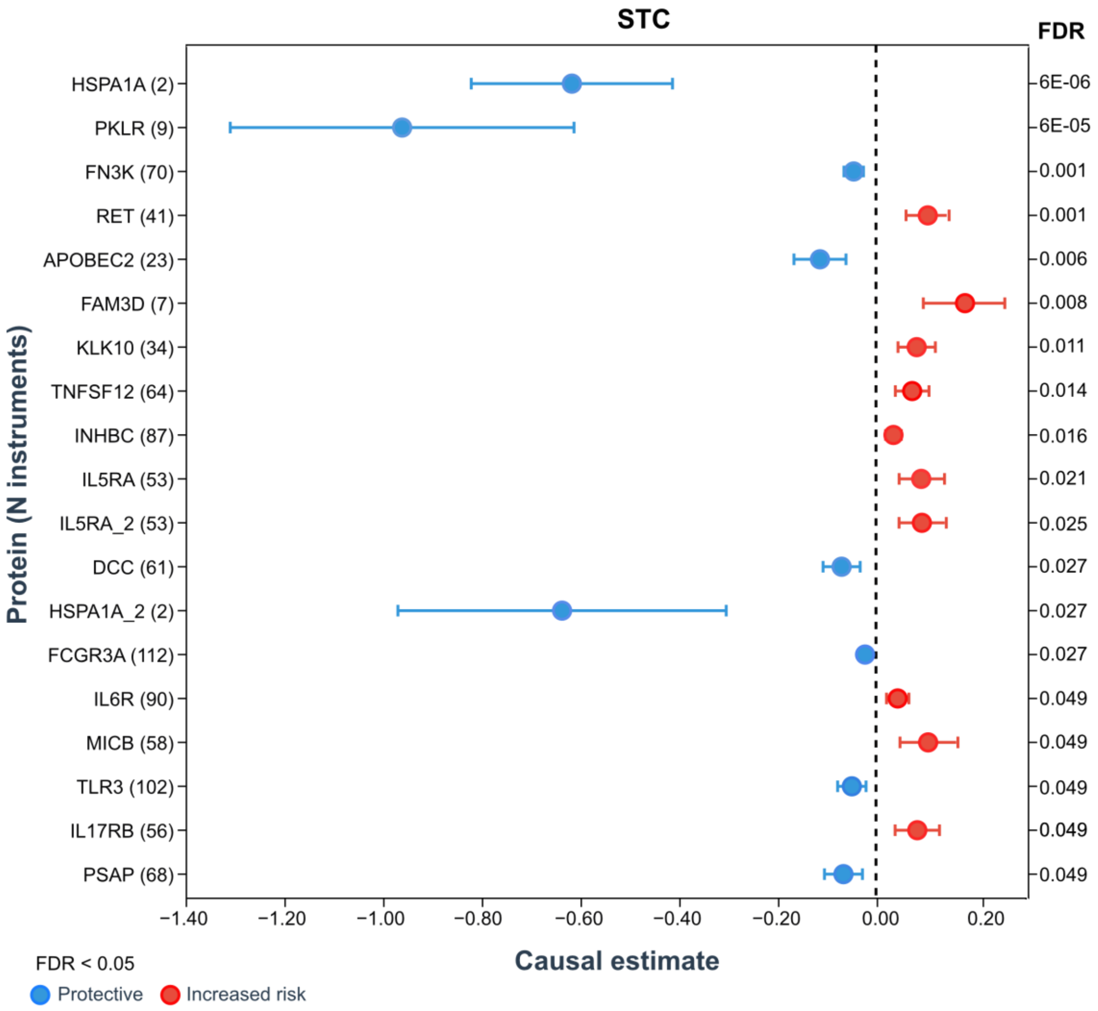


**Fig. S14.** Mendelian randomization analysis of protein vs. stomach cancer (STC) associations meeting the FDR < 0.05 significance threshold, ranked by effect significance (FDR). Color coding denotes directionality: red indicates risk-increasing causal relationships, while blue represents potentially protective associations. The y-axis shows protein identifiers with genetic instrument counts in parentheses. Duplicate protein entries correspond to distinct aptamer probes exhibiting independent causal effects. Corresponding FDR values are presented on the right side of the plot..

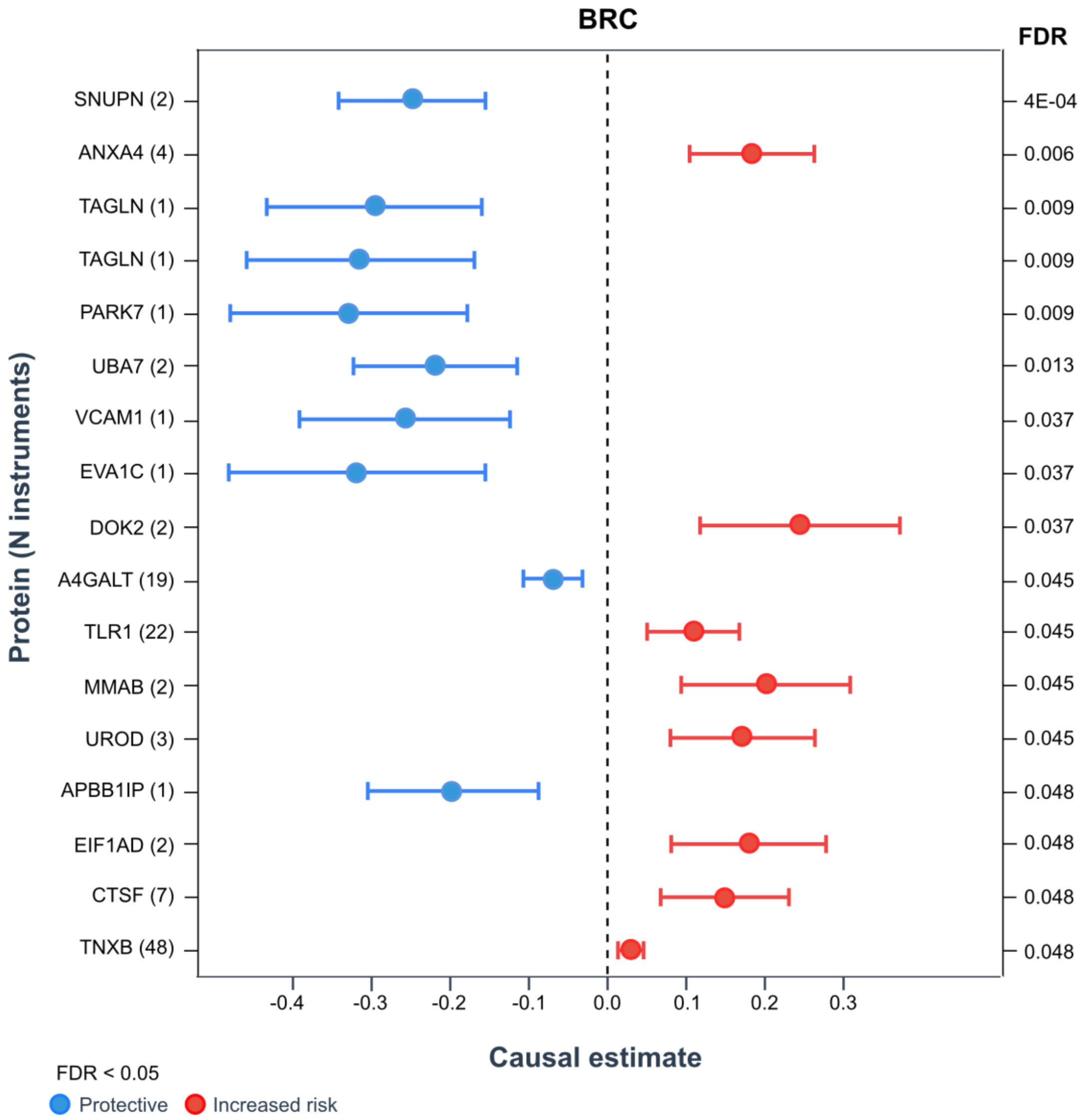


**Fig. S15**. Forest plot depicting MR results for breast cancer (BRC) at FDR < 0.05, ranked by association significance. Red dots represent proteins with positive causal effects (increased risk), while blue dots indicate negative causal effects (potentially protective). Protein names are shown on the y-axis with the number of genetic instruments in parentheses. Multiple entries for the same protein reflect distinct aptamers showing significant causal associations. FDR values are displayed on the right.

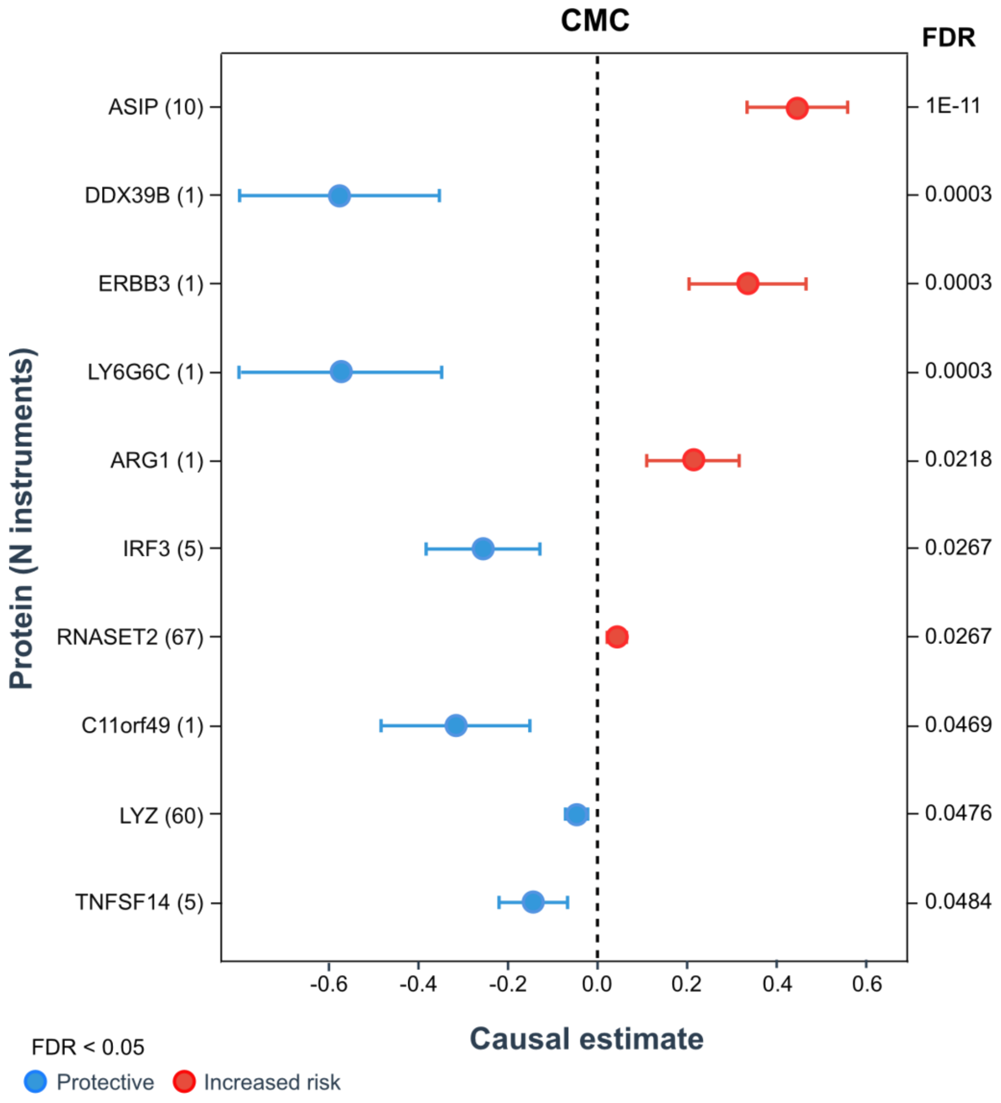


**Fig. S16**. Forest plot of MR results for cutaneous melanoma cancer (CMC) at FDR < 0.05. Proteins with the strongest associations are shown toward the top. Red dots represent positive causal associations (increased disease risk), while blue dots indicate negative associations (potentially protective effects). The y-axis lists protein annotations with the number of instruments in parentheses. Repeated protein names denote multiple aptamers showing potential causal associations with CMC. The corresponding false discovery rate (FDR) for each test is displayed to the right of the plot.

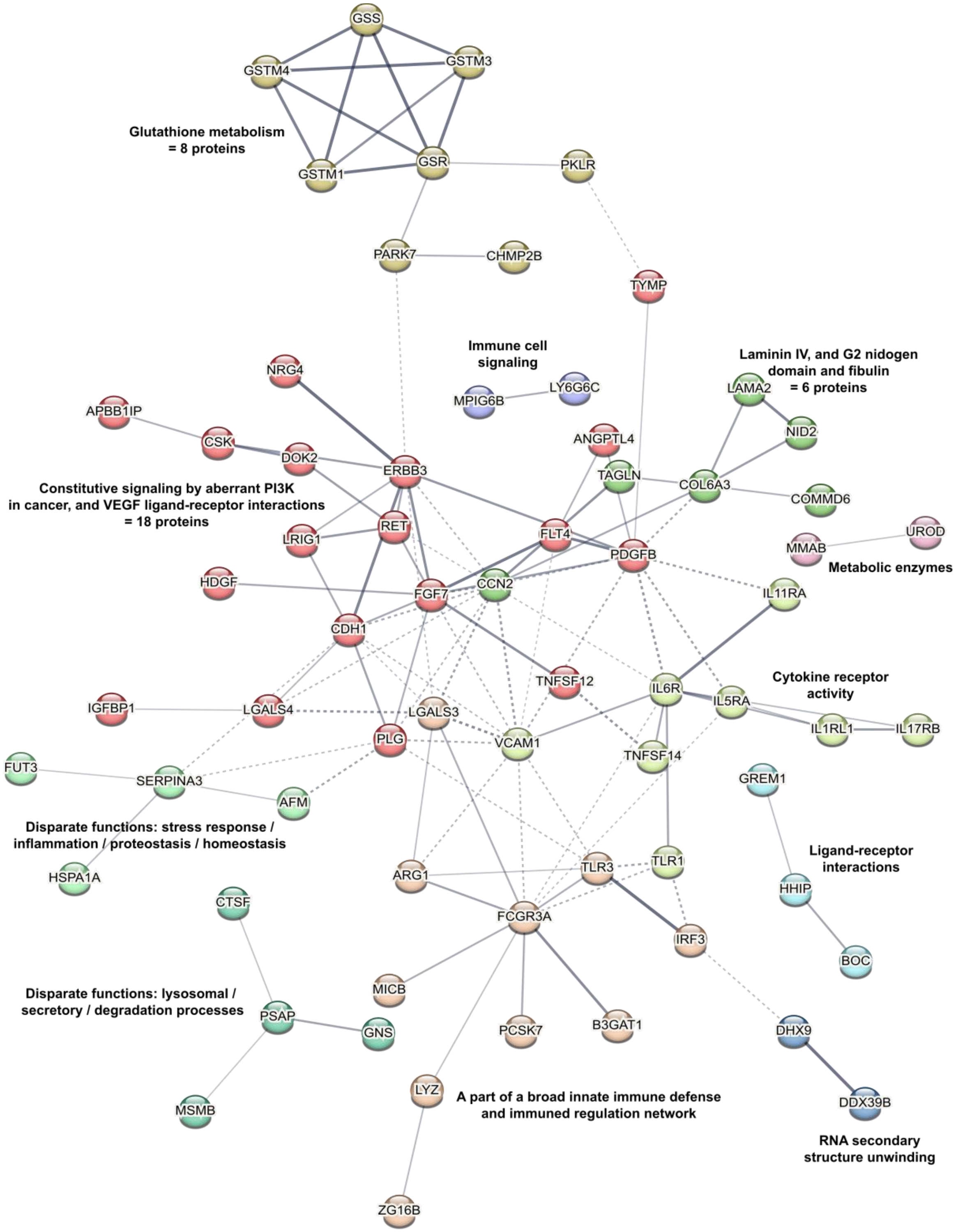


**Fig. S17**. Protein–protein interaction networks among serum proteins with evidence of a causal relationship to cancer (FDR < 0.05), as identified using the STRING database. Solid edges represent functional or physical interactions within the 11 clusters, while unconnected network regulators were omitted from the visualization. Dotted lines indicate interactions between clusters. The PPI enrichment P-value was $9 \times 10^{-14}$ (115 edges observed versus 53 expected).

#### References


1. Harris, T.B., *et al.* Age, Gene/Environment Susceptibility-Reykjavik Study: multidisciplinary applied phenomics. *Am J Epidemiol* **165**, 1076-1087 (2007).
2. Cox, D.R. Regression Models and Life-Tables. **34**, 187-202 (1972).
3. Pernar, C.H., Ebot, E.M., Wilson, K.M. & Mucci, L.A. The Epidemiology of Prostate Cancer. *Cold Spring Harbor perspectives in medicine* **8**(2018).
4. Chow, W.H., Dong, L.M. & Devesa, S.S. Epidemiology and risk factors for kidney cancer. *Nature reviews. Urology* **7**, 245-257 (2010).
5. Robinson, L.D. & Jewell, N.P. Some Surprising Results about Covariate Adjustment in Logistic Regression Models. *International Statistical Review / Revue Internationale de Statistique* **59**, 227-240 (1991).
6. Benjamini, Y. & Hochberg, Y. Controlling the False Discovery Rate: A Practical and Powerful Approach to Multiple Testing. *Journal of the Royal Statistical Society* **Vol.57**, No. 1: 289-300 (1995).
7. Tang, Z., *et al.* GEPIA: a web server for cancer and normal gene expression profiling and interactive analyses. *Nucleic Acids Res* **45**, W98-w102 (2017).
8. Consortium, G. The GTEx Consortium atlas of genetic regulatory effects across human tissues. *Science* **369**, 1318-1330 (2020).
9. Buniello, A., *et al.* The NHGRI-EBI GWAS Catalog of published genome-wide association studies, targeted arrays and summary statistics 2019. *Nucleic Acids Res* **47**, D1005-d1012 (2019).
10. Kurki, M.I., *et al.* FinnGen provides genetic insights from a well-phenotyped isolated population. *Nature* **613**, 508-518 (2023).
11. Schwenk, J.M., *et al.* The Human Plasma Proteome Draft of 2017: Building on the Human Plasma PeptideAtlas from Mass Spectrometry and Complementary Assays. *J Proteome Res* **16**, 4299-4310 (2017).
12. Arnold, M., *et al.* Global Burden of 5 Major Types of Gastrointestinal Cancer. *Gastroenterology* **159**, 335-349.e315 (2020).
13. Pennathur, A., Gibson, M.K., Jobe, B.A. & Luketich, J.D. Oesophageal carcinoma. *Lancet (London, England)* **381**, 400-412 (2013).
14. Stabellini, N., *et al.* Sex differences in esophageal cancer overall and by histological subtype. *Sci Rep* **12**, 5248 (2022).
15. Futreal, P.A., *et al.* A census of human cancer genes. *Nat Rev Cancer* **4**, 177-183 (2004).
16. Ferlay, J., *et al.* Cancer incidence and mortality worldwide: sources, methods and major patterns in GLOBOCAN 2012. *International journal of cancer* **136**, E359-386 (2015).
17. de Martel, C., Forman, D. & Plummer, M. Gastric cancer: epidemiology and risk factors. *Gastroenterology clinics of North America* **42**, 219-240 (2013).
18. Liu, F. & Wu, H. Prognostic Value of Gastrokine-2 (GKN2) and Its Correlation with Tumor-Infiltrating Immune Cells in Lung Cancer and Gastric Cancers. *Journal of inflammation research* **13**, 933-944 (2020).
19. Chen, Z., Downing, S. & Tzanakakis, E.S. Four Decades After the Discovery of Regenerating Islet-Derived (Reg) Proteins: Current Understanding and Challenges. *Frontiers in cell and developmental biology* **7**, 235 (2019).
20. Chung Nien Chin, S., *et al.* Coordinate expression loss of GKN1 and GKN2 in gastric cancer via impairment of a glucocorticoid-responsive enhancer. *American journal of physiology. Gastrointestinal and liver physiology* **319**, G175-g188 (2020).
21. Hoffmann, W. Trefoil Factor Family (TFF) Peptides and Their Links to Inflammation: A Re-evaluation and New Medical Perspectives. *International journal of molecular sciences* **22**(2021).
22. Kirikoshi, H. & Katoh, M. Expression of TFF1, TFF2 and TFF3 in gastric cancer. *International journal of oncology* **21**, 655-659 (2002).
23. Westley, B.R., Griffin, S.M. & May, F.E. Interaction between TFF1, a gastric tumor suppressor trefoil protein, and TFIZ1, a brichos domain-containing protein with homology to SP-C. *Biochemistry* **44**, 7967-7975 (2005).

24. Moss, S.F., *et al.* Decreased expression of gastrokine 1 and the trefoil factor interacting protein TFIZ1/GKN2 in gastric cancer: influence of tumor histology and relationship to prognosis. *Clin Cancer Res* **14**, 4161-4167 (2008).
25. Kim, O., *et al.* Heterodimeric interaction between GKN2 and TFF1 entails synergistic antiproliferative and pro-apoptotic effects on gastric cancer cells. *Gastric cancer : official journal of the International Gastric Cancer Association and the Japanese Gastric Cancer Association* **20**, 772-783 (2017).
26. Li, Z., *et al.* Immune checkpoint reprogramming via sequential nucleic acid delivery strategy optimizes systemic immune responses for gastrointestinal cancer immunotherapy. *Cancer letters* **599**, 217152 (2024).
27. Dekker, E., Tanis, P.J., Vleugels, J.L.A., Kasi, P.M. & Wallace, M.B. Colorectal cancer. *Lancet* **394**, 1467-1480 (2019).
28. Paschke, S., *et al.* Are Colon and Rectal Cancer Two Different Tumor Entities? A Proposal to Abandon the Term Colorectal Cancer. *International journal of molecular sciences* **19**(2018).
29. Sung, H., *et al.* Global Cancer Statistics 2020: GLOBOCAN Estimates of Incidence and Mortality Worldwide for 36 Cancers in 185 Countries. *CA Cancer J Clin* **71**, 209-249 (2021).
30. Law, P.J., *et al.* Association analyses identify 31 new risk loci for colorectal cancer susceptibility. *Nat Commun* **10**, 2154 (2019).
31. Yu, J., Feng, Q., Kim, J.H. & Zhu, Y. Combined Effect of Healthy Lifestyle Factors and Risks of Colorectal Adenoma, Colorectal Cancer, and Colorectal Cancer Mortality: Systematic Review and Meta-Analysis. *Frontiers in oncology* **12**, 827019 (2022).
32. Signs, S.A., *et al.* Stromal miR-20a controls paracrine CXCL8 secretion in colitis and colon cancer. *Oncotarget* **9**, 13048-13059 (2018).
33. Vierthaler, M., *et al.* ADCK2 Knockdown Affects the Migration of Melanoma Cells via MYL6. *Cancers* **14**(2022).
34. Low, H.B., *et al.* DUSP16 promotes cancer chemoresistance through regulation of mitochondria-mediated cell death. *Nat Commun* **12**, 2284 (2021).
35. Pennel, K.A., *et al.* CXCL8 expression is associated with advanced stage, right sidedness, and distinct histological features of colorectal cancer. *The journal of pathology. Clinical research* **8**, 509-520 (2022).
36. Shimizu, M. & Tanaka, N. IL-8-induced O-GlcNAc modification via GLUT3 and GFAT regulates cancer stem cell-like properties in colon and lung cancer cells. *Oncogene* **38**, 1520-1533 (2019).
37. Jia, S.N., Han, Y.B., Yang, R. & Yang, Z.C. Chemokines in colon cancer progression. *Seminars in cancer biology* **86**, 400-407 (2022).
38. Siculella, L., *et al.* A comprehensive understanding of hnRNP A1 role in cancer: new perspectives on binding with noncoding RNA. *Cancer gene therapy* **30**, 394-403 (2023).
39. Vangilbergen, M., *et al.* The role of interleukin-17 and interleukin-23 inhibitors in the development, progression, and recurrence of cancer: A systematic review. *JAAD international* **17**, 71-79 (2024).
40. Jang, S.M., *et al.* Loss of Wnt7a expression correlates with tumor progression and poor prognosis in colorectal carcinoma. *International journal of clinical and experimental pathology* **11**, 4967-4976 (2018).
41. Rahib, L., *et al.* Projecting cancer incidence and deaths to 2030: the unexpected burden of thyroid, liver, and pancreas cancers in the United States. *Cancer Res* **74**, 2913-2921 (2014).
42. Zou, W., *et al.* Up-regulation of S100P predicts the poor long-term survival and construction of prognostic signature for survival and immunotherapy in patients with pancreatic cancer. *Bioengineered* **12**, 9006-9020 (2021).
43. Marciel, M.P., Haldar, B., Hwang, J., Bhalerao, N. & Bellis, S.L. Role of tumor cell sialylation in pancreatic cancer progression. *Advances in cancer research* **157**, 123-155 (2023).
44. Siegel, R.L., Miller, K.D. & Jemal, A. Cancer statistics, 2020. *CA Cancer J Clin* **70**, 7-30 (2020).
45. Bray, F., *et al.* Global cancer statistics 2018: GLOBOCAN estimates of incidence and mortality worldwide for 36 cancers in 185 countries. *CA Cancer J Clin* **68**, 394-424 (2018).
46. Mancini, M., Righetto, M. & Baggio, G. Gender-Related Approach to Kidney Cancer Management: Moving Forward. *International journal of molecular sciences* **21**(2020).
47. Vilà, M.R., *et al.* Hepatitis A virus receptor blocks cell differentiation and is overexpressed in clear cell renal cell carcinoma. *Kidney Int* **65**, 1761-1773 (2004).
48. Uhlen, M., *et al.* Tissue-based map of the human proteome. *Science* **347**(2015).

49. Huang, Z., *et al.* Key role for EphB2 receptor in kidney fibrosis. *Clin Sci (Lond)* **135**, 2127-2142 (2021).
50. Fitzmaurice, C., *et al.* Global, Regional, and National Cancer Incidence, Mortality, Years of Life Lost, Years Lived With Disability, and Disability-Adjusted Life-years for 32 Cancer Groups, 1990 to 2015: A Systematic Analysis for the Global Burden of Disease Study. *JAMA oncology* **3**, 524-548 (2017).
51. Lilja, H., Ulmert, D. & Vickers, A.J. Prostate-specific antigen and prostate cancer: prediction, detection and monitoring. *Nat Rev Cancer* **8**, 268-278 (2008).
52. Guy, M., *et al.* Identification of new genetic risk factors for prostate cancer. *Asian journal of andrology* **11**, 49-55 (2009).
53. Smith Byrne, K., *et al.* The role of plasma microseminoprotein-beta in prostate cancer: an observational nested case-control and Mendelian randomization study in the European prospective investigation into cancer and nutrition. *Annals of oncology : official journal of the European Society for Medical Oncology* **30**, 983-989 (2019).
54. Antoni, S., *et al.* Bladder Cancer Incidence and Mortality: A Global Overview and Recent Trends. *European urology* **71**, 96-108 (2017).
55. Burger, M., *et al.* Epidemiology and risk factors of urothelial bladder cancer. *European urology* **63**, 234-241 (2013).
56. Nagata, Y., *et al.* Mineralocorticoid receptor signaling inhibits bladder cancer progression. *American journal of cancer research* **14**, 696-708 (2024).
57. Li, J. & Xu, Z. NR3C2 suppresses the proliferation, migration, invasion and angiogenesis of colon cancer cells by inhibiting the AKT/ERK signaling pathway. *Mol Med Rep* **25**(2022).
58. Liu, Y., *et al.* IL1R2 promotes tumor progression via JAK2/STAT3 pathway in human clear cell renal cell carcinoma. *Pathology, research and practice* **238**, 154069 (2022).
59. Lang, Y., *et al.* IL-1R2 promotes tumorigenesis and modulates the tumor immune microenvironment in colorectal cancer. *Cancer immunology, immunotherapy : CII* **74**, 284 (2025).
60. Global, regional, and national burden of respiratory tract cancers and associated risk factors from 1990 to 2019: a systematic analysis for the Global Burden of Disease Study 2019. *Lancet Respir Med* **9**, 1030-1049 (2021).
61. Houghton, S.C. & Hankinson, S.E. Cancer Progress and Priorities: Breast Cancer. *Cancer epidemiology, biomarkers & prevention : a publication of the American Association for Cancer Research, cosponsored by the American Society of Preventive Oncology* **30**, 822-844 (2021).
62. Zhang, T., Chu, L., Tan, W., Ye, C. & Dong, H. Human epididymis protein 4, a novel potential biomarker for diagnostic and prognosis monitoring of lung cancer. *The clinical respiratory journal* **18**, e13774 (2024).
63. Min, B. & Wang, Y. WFDC2 is a potential prognostic and immunotherapy biomarker in lung adenocarcinoma. *The Journal of international medical research* **52**, 3000605241258893 (2024).
64. Tomita, T. & Kimura, S. Regulation of mouse Scgb3a1 gene expression by NF-Y and association of CpG methylation with its tissue-specific expression. *BMC molecular biology* **9**, 5 (2008).
65. Zhang, X., *et al.* The prognostic significance and Immunomodulatory role of SCGB3A1 expression in stage I lung adenocarcinoma. *BMC medical genomics* **18**, 119 (2025).
66. Sun, J., *et al.* CLEC3B as a potential diagnostic and prognostic biomarker in lung cancer and association with the immune microenvironment. *Cancer cell international* **20**, 106 (2020).
67. Xie, X.W., Jiang, S.S. & Li, X. CLEC3B as a Potential Prognostic Biomarker in Hepatocellular Carcinoma. *Frontiers in molecular biosciences* **7**, 614034 (2020).
68. Cohen, S.Y., Stoll, C.R., Anandarajah, A., Doering, M. & Colditz, G.A. Modifiable risk factors in women at high risk of breast cancer: a systematic review. *Breast cancer research : BCR* **25**, 45 (2023).
69. Petrova, D., Cruz, M. & Sánchez, M.J. BRCA1/2 testing for genetic susceptibility to cancer after 25 years: A scoping review and a primer on ethical implications. *Breast (Edinburgh, Scotland)* **61**, 66-76 (2022).
70. El Ayachi, I., *et al.* The WNT10B Network Is Associated with Survival and Metastases in Chemoresistant Triple-Negative Breast Cancer. *Cancer Res* **79**, 982-993 (2019).
71. Veeck, J., *et al.* Wnt signalling in human breast cancer: expression of the putative Wnt inhibitor Dickkopf-3 (DKK3) is frequently suppressed by promoter hypermethylation in mammary tumours. *Breast Cancer Research* **10**, R82 (2008).

72. Kim, S.W., Choi, J.W., Lee, D.S. & Yun, J.W. Sex hormones regulate hepatic fetuin expression in male and female rats. *Cellular physiology and biochemistry : international journal of experimental cellular physiology, biochemistry, and pharmacology* **34**, 554-564 (2014).
73. Siegel, R.L., Miller, K.D., Wagle, N.S. & Jemal, A. Cancer statistics, 2023. *CA Cancer J Clin* **73**, 17-48 (2023).
74. Allen, N.E.*, et al.* Endogenous sex hormones and endometrial cancer risk in women in the European Prospective Investigation into Cancer and Nutrition (EPIC). *Endocrine-related cancer* **15**, 485-497 (2008).
75. Kyrgiou, M.*, et al.* Adiposity and cancer at major anatomical sites: umbrella review of the literature. *Bmj* **356**, j477 (2017).
76. Takeshita, T.*, et al.* PTIP associated protein 1, PA1, is an independent prognostic factor for lymphnode negative breast cancer. *PLoS One* **8**, e80552 (2013).
77. Zhang, M.*, et al.* UBE2S promotes the development of ovarian cancer by promoting PI3K/AKT/mTOR signaling pathway to regulate cell cycle and apoptosis. *Molecular medicine (Cambridge, Mass.)* **28**, 62 (2022).
78. Zhang, M.*, et al.* Diverse roles of UBE2S in cancer and therapy resistance: Biological functions and mechanisms. *Heliyon* **10**, e24465 (2024).
79. Liao, X.*, et al.* OSCAR facilitates malignancy with enhanced metastasis correlating to inhibitory immune microenvironment in multiple cancer types. *Journal of Cancer* **12**, 3769-3780 (2021).
80. Lelièvre, P., Sancey, L., Coll, J.L., Deniaud, A. & Busser, B. The Multifaceted Roles of Copper in Cancer: A Trace Metal Element with Dysregulated Metabolism, but Also a Target or a Bullet for Therapy. *Cancers* **12**(2020).
81. Coburn, S.B., Bray, F., Sherman, M.E. & Trabert, B. International patterns and trends in ovarian cancer incidence, overall and by histologic subtype. *International journal of cancer* **140**, 2451-2460 (2017).
82. Jones, M.R., Kamara, D., Karlan, B.Y., Pharoah, P.D.P. & Gayther, S.A. Genetic epidemiology of ovarian cancer and prospects for polygenic risk prediction. *Gynecologic oncology* **147**, 705-713 (2017).
83. Miki, Y.*, et al.* A strong candidate for the breast and ovarian cancer susceptibility gene BRCA1. *Science* **266**, 66-71 (1994).
84. Wooster, R.*, et al.* Identification of the breast cancer susceptibility gene BRCA2. *Nature* **378**, 789-792 (1995).
85. Kriška, J.*, et al.* Human erythropoietin increases the pro-angiogenic potential of A2780 ovarian adenocarcinoma cells under hypoxic conditions. *Oncology reports* **30**, 1455-1462 (2013).
86. Paragh, G.*, et al.* RNA interference-mediated inhibition of erythropoietin receptor expression suppresses tumor growth and invasiveness in A2780 human ovarian carcinoma cells. *The American journal of pathology* **174**, 1504-1514 (2009).
87. Hu, J.*, et al.* TMEM166/EVA1A interacts with ATG16L1 and induces autophagosome formation and cell death. *Cell death & disease* **7**, e2323 (2016).
88. Xu, Q.*, et al.* Down-regulation of EVA1A by miR-103a-3p promotes hepatocellular carcinoma cells proliferation and migration. *Cellular & molecular biology letters* **27**, 93 (2022).
89. Siegel, R.L., Miller, K.D. & Jemal, A. Cancer statistics, 2016. *CA Cancer J Clin* **66**, 7-30 (2016).
90. Slominski, R.M.*, et al.* Malignant Melanoma: An Overview, New Perspectives, and Vitamin D Signaling. *Cancers* **16**(2024).
91. Karimkhani, C.*, et al.* The global burden of melanoma: results from the Global Burden of Disease Study 2015. *The British journal of dermatology* **177**, 134-140 (2017).
92. Bolick, N.L. & Geller, A.C. Epidemiology and Screening for Melanoma. *Hematology/oncology clinics of North America* (2024).
93. Batta, N.*, et al.* Global melanoma correlations with obesity, smoking, and alcohol consumption. *JMIR dermatology* **4**, e31275 (2021).
94. Paduano, F., Gaudio, E. & Trapasso, F. The Tumour Suppressor Fhit Protein Activates C-Raf Ubiquitination and Degradation in Human Melanoma Cells by Interacting with Hsp90. *Biomedicines* **10**(2022).
95. Grabenstein, S.*, et al.* Deacetylated sialic acids modulates immune mediated cytotoxicity via the sialic acid-Siglec pathway. *Glycobiology* **31**, 1279-1294 (2021).

96. Moretti, F., *et al.* TMEM41B is a novel regulator of autophagy and lipid mobilization. *EMBO Rep* **19**(2018).
97. Villarreal-García, V., *et al.* Inhibition of microRNA-660-5p decreases breast cancer progression through direct targeting of TMEM41B. *Hereditas* **161**, 53 (2024).
98. Brown, K.F., *et al.* The fraction of cancer attributable to modifiable risk factors in England, Wales, Scotland, Northern Ireland, and the United Kingdom in 2015. *British journal of cancer* **118**, 1130-1141 (2018).
99. Gapstur, S.M., *et al.* Alcohol and Cancer: Existing Knowledge and Evidence Gaps across the Cancer Continuum. *Cancer epidemiology, biomarkers & prevention : a publication of the American Association for Cancer Research, cosponsored by the American Society of Preventive Oncology* **31**, 5-10 (2022).
100. Ali, M.W., *et al.* A risk variant for Barrett's esophagus and esophageal adenocarcinoma at chr8p23.1 affects enhancer activity and implicates multiple gene targets. *Hum Mol Genet* **31**, 3975-3986 (2022).
101. Siegel, R.L., Miller, K.D., Fuchs, H.E. & Jemal, A. Cancer statistics, 2022. *CA Cancer J Clin* **72**, 7-33 (2022).
102. Mazumdar, J., *et al.* HIF-2alpha deletion promotes Kras-driven lung tumor development. *Proc Natl Acad Sci U S A* **107**, 14182-14187 (2010).
103. Krop, I., *et al.* HIN-1, an inhibitor of cell growth, invasion, and AKT activation. *Cancer Res* **65**, 9659-9669 (2005).
104. Reynolds, S.D., Reynolds, P.R., Pryhuber, G.S., Finder, J.D. & Stripp, B.R. Secretoglobins SCGB3A1 and SCGB3A2 define secretory cell subsets in mouse and human airways. *American journal of respiratory and critical care medicine* **166**, 1498-1509 (2002).
105. Goldfarbmuren, K.C., *et al.* Dissecting the cellular specificity of smoking effects and reconstructing lineages in the human airway epithelium. *Nat Commun* **11**, 2485 (2020).
106. Lu, X., Shen, J., Huang, S., Wang, H. & Liu, D. Down-regulation of CLEC3B facilitates epithelial-mesenchymal transition, migration and invasion of lung adenocarcinoma cells. *Tissue & cell* **76**, 101802 (2022).
107. Ungaro, F., *et al.* Lymphatic endothelium contributes to colorectal cancer growth via the soluble matrisome component GDF11. *International journal of cancer* **145**, 1913-1920 (2019).
108. Ojima, C., *et al.* Peptide-2 from mouse myostatin precursor protein alleviates muscle wasting in cancer-associated cachexia. *Cancer science* **111**, 2954-2964 (2020).
109. Xu, T., *et al.* ApoM suppresses kidney renal clear cell carcinoma growth and metastasis via the Hippo-YAP signaling pathway. *Archives of biochemistry and biophysics* **743**, 109642 (2023).
110. Thapa, B., *et al.* OX40/OX40 ligand and its role in precision immune oncology. *Cancer metastasis reviews* **43**, 1001-1013 (2024).
111. Liu, K., Li, L. & Han, G. CHST12: a potential prognostic biomarker related to the immunotherapy response in pancreatic adenocarcinoma. *Frontiers in endocrinology* **14**, 1226547 (2023).
112. Chandrashekar, D.S., *et al.* UALCAN: An update to the integrated cancer data analysis platform. *Neoplasia (New York, N.Y.)* **25**, 18-27 (2022).
113. Nguyen, P.M., *et al.* Loss of Bcl-G, a Bcl-2 family member, augments the development of inflammation-associated colorectal cancer. *Cell death and differentiation* **27**, 742-757 (2020).
114. Bai, J., *et al.* Overexpression of Cullin1 is associated with poor prognosis of patients with gastric cancer. *Human pathology* **42**, 375-383 (2011).
115. Leblond, M.M., Zdimerova, H., Desponds, E. & Verdeil, G. Tumor-Associated Macrophages in Bladder Cancer: Biological Role, Impact on Therapeutic Response and Perspectives for Immunotherapy. *Cancers* **13**(2021).
116. Maldonado, M.D.M., Schlom, J. & Hamilton, D.H. Blockade of tumor-derived colony-stimulating factor 1 (CSF1) promotes an immune-permissive tumor microenvironment. *Cancer immunology, immunotherapy : CII* **72**, 3349-3362 (2023).
117. Tan, D.X. & Hardeland, R. The Reserve/Maximum Capacity of Melatonin's Synthetic Function for the Potential Dimorphism of Melatonin Production and Its Biological Significance in Mammals. *Molecules (Basel, Switzerland)* **26**(2021).
118. Masri, S. & Sassone-Corsi, P. The emerging link between cancer, metabolism, and circadian rhythms. *Nat Med* **24**, 1795-1803 (2018).

119. Ishibashi, Y., *et al.* Serum TFF1 and TFF3 but not TFF2 are higher in women with breast cancer than in women without breast cancer. *Sci Rep* **7**, 4846 (2017).
120. Zhang, Y., Liu, Y., Wang, L. & Song, H. The expression and role of trefoil factors in human tumors. *Translational cancer research* **8**, 1609-1617 (2019).
121. Cohen, J.D., *et al.* Detection and localization of surgically resectable cancers with a multi-analyte blood test. *Science* **359**, 926-930 (2018).
122. Pin, E., Fredolini, C. & Petricoin, E.F., 3rd. The role of proteomics in prostate cancer research: biomarker discovery and validation. *Clinical biochemistry* **46**, 524-538 (2013).
123. Lokshin, A., Bast, R.C. & Rodland, K. Circulating Cancer Biomarkers. *Cancers* **13**(2021).
124. Emilsson, V., *et al.* Co-regulatory networks of human serum proteins link genetics to disease. *Science* **361**, 769-773 (2018).
125. Sun, B.B., *et al.* Genomic atlas of the human plasma proteome. *Nature* **558**, 73-79 (2018).
126. Sun, B.B., *et al.* Plasma proteomic associations with genetics and health in the UK Biobank. *Nature* **622**, 329-338 (2023).
127. Gudjonsson, A., *et al.* A genome-wide association study of serum proteins reveals shared loci with common diseases. *Nature Communications* **13**, 1-13 (2022).
128. Suehnholz, S.P., *et al.* Quantifying the Expanding Landscape of Clinical Actionability for Patients with Cancer. *Cancer Discov* **14**, 49-65 (2024).
129. Chakravarty, D., *et al.* OncoKB: A Precision Oncology Knowledge Base. *JCO Precis Oncol* **17**, 16 (2017).
130. Perez-Garcia, A., *et al.* Genetic loss of SH2B3 in acute lymphoblastic leukemia. *Blood* **122**, 2425-2432 (2013).
131. Wen, R.M., *et al.* AZGP1 deficiency promotes angiogenesis in prostate cancer. *Journal of translational medicine* **22**, 383 (2024).
132. Deng, L., *et al.* AZGP1 activation by lenvatinib suppresses intrahepatic cholangiocarcinoma epithelial-mesenchymal transition through the TGF-β1/Smad3 pathway. *Cell death & disease* **14**, 590 (2023).
133. Lindström, S., *et al.* Genome-wide analyses characterize shared heritability among cancers and identify novel cancer susceptibility regions. *Journal of the National Cancer Institute* **115**, 712-732 (2023).
134. Rashkin, S.R., *et al.* Pan-cancer study detects genetic risk variants and shared genetic basis in two large cohorts. *Nat Commun* **11**, 4423 (2020).
135. Gharahkhani, P., *et al.* Genome-wide association studies in oesophageal adenocarcinoma and Barrett's oesophagus: a large-scale meta-analysis. *The Lancet. Oncology* **17**, 1363-1373 (2016).
136. Helgason, H., *et al.* Loss-of-function variants in ATM confer risk of gastric cancer. *Nat Genet* **47**, 906-910 (2015).
137. Hess, T., *et al.* Dissecting the genetic heterogeneity of gastric cancer. *EBioMedicine* **92**, 104616 (2023).
138. Sakaue, S., *et al.* A cross-population atlas of genetic associations for 220 human phenotypes. *Nat Genet* **53**, 1415-1424 (2021).
139. Menheniott, T.R., *et al.* Loss of gastrokine-2 drives premalignant gastric inflammation and tumor progression. *J Clin Invest* **126**, 1383-1400 (2016).
140. Joshi, N., *et al.* Urinary Proteomics for Discovery of Gastric Cancer Biomarkers to Enable Precision Clinical Oncology. *Omics : a journal of integrative biology* **27**, 361-371 (2023).
141. Fernandez-Rozadilla, C., *et al.* Deciphering colorectal cancer genetics through multi-omic analysis of 100,204 cases and 154,587 controls of European and east Asian ancestries. *Nat Genet* **55**, 89-99 (2023).
142. Emilsson, V., *et al.* Coding and regulatory variants are associated with serum protein levels and disease. *Nature Communications* **13**, 1-11 (2022).
143. Kim, J.W., Rim, D., Ann, C.H. & Lee, S.H. Frameshift mutations of immunomodulatory BTN2A1, BTN2A2, and BTNL3 genes in colon cancers. *Pathology, research and practice* **249**, 154769 (2023).
144. Adib, E., *et al.* CDH1 germline variants are enriched in patients with colorectal cancer, gastric cancer, and breast cancer. *British journal of cancer* **126**, 797-803 (2022).
145. Bartolomé, R.A., *et al.* Loss of cadherin 17 downregulates LGR5 expression, stem cell properties and drug resistance in metastatic colorectal cancer cells. *Cell death & disease* **16**, 475 (2025).

146. Zhu, M., *et al.* Silence of a dependence receptor CSF1R in colorectal cancer cells activates tumor-associated macrophages. *Journal for immunotherapy of cancer* **10**(2022).
147. Klein, A.P., *et al.* Genome-wide meta-analysis identifies five new susceptibility loci for pancreatic cancer. *Nat Commun* **9**, 556 (2018).
148. Gentiluomo, M., *et al.* Germline genetic variability in pancreatic cancer risk and prognosis. *Seminars in cancer biology* **79**, 105-131 (2022).
149. Zhang, G., *et al.* Integration of metabolomics and transcriptomics revealed a fatty acid network exerting growth inhibitory effects in human pancreatic cancer. *Clin Cancer Res* **19**, 4983-4993 (2013).
150. Purdue, M.P., *et al.* Multi-ancestry genome-wide association study of kidney cancer identifies 63 susceptibility regions. *Nat Genet* (2024).
151. Mucci, L.A., *et al.* Familial Risk and Heritability of Cancer Among Twins in Nordic Countries. *Jama* **315**, 68-76 (2016).
152. Koutros, S., *et al.* Genome-wide Association Study of Bladder Cancer Reveals New Biological and Translational Insights. *European urology* **84**, 127-137 (2023).
153. Figueroa, J.D., *et al.* Genome-wide association study identifies multiple loci associated with bladder cancer risk. *Hum Mol Genet* **23**, 1387-1398 (2014).
154. Pettit, R.W., *et al.* The shared genetic architecture between epidemiological and behavioral traits with lung cancer. *Sci Rep* **11**, 17559 (2021).
155. Oxnard, G.R., *et al.* Germline EGFR Mutations and Familial Lung Cancer. *J Clin Oncol* **41**, 5274-5284 (2023).
156. Byun, J., *et al.* Cross-ancestry genome-wide meta-analysis of 61,047 cases and 947,237 controls identifies new susceptibility loci contributing to lung cancer. *Nat Genet* **54**, 1167-1177 (2022).
157. McGraw, J.M., *et al.* JAML promotes CD8 and γδ T cell antitumor immunity and is a novel target for cancer immunotherapy. *The Journal of experimental medicine* **218**(2021).
158. Michailidou, K., *et al.* Association analysis identifies 65 new breast cancer risk loci. *Nature* **551**, 92-94 (2017).
159. O'Mara, T.A., *et al.* Identification of nine new susceptibility loci for endometrial cancer. *Nat Commun* **9**, 3166 (2018).
160. Kuchenbaecker, K.B., *et al.* Identification of six new susceptibility loci for invasive epithelial ovarian cancer. *Nat Genet* **47**, 164-171 (2015).
161. Phelan, C.M., *et al.* Identification of 12 new susceptibility loci for different histotypes of epithelial ovarian cancer. *Nat Genet* **49**, 680-691 (2017).
162. Gooden, M.J., *et al.* Elevated serum CXCL16 is an independent predictor of poor survival in ovarian cancer and may reflect pro-metastatic ADAM protease activity. *British journal of cancer* **110**, 1535-1544 (2014).
163. Landi, M.T., *et al.* Genome-wide association meta-analyses combining multiple risk phenotypes provide insights into the genetic architecture of cutaneous melanoma susceptibility. *Nat Genet* **52**, 494-504 (2020).
164. Blanchard, S.G., *et al.* Agouti antagonism of melanocortin binding and action in the B16F10 murine melanoma cell line. *Biochemistry* **34**, 10406-10411 (1995).
165. Bankier, S., *et al.* Circulating causal protein networks linked to future risk of myocardial infarction. *medRxiv* (2025).
166. Szklarczyk, D., *et al.* STRING v11: protein-protein association networks with increased coverage, supporting functional discovery in genome-wide experimental datasets. *Nucleic Acids Res* **47**, D607-d613 (2019).
167. Jonmundsson, T., *et al.* Systematic comparison of observational and Mendelian Randomization estimates for cardiometabolic proteomic signatures. 2025.2012.2002.25341031 (2025).
168. Li, R., *et al.* Gremlin-1 Promotes Colorectal Cancer Cell Metastasis by Activating ATF6 and Inhibiting ATF4 Pathways. *Cells* **11**(2022).
169. Yu, Q., *et al.* GREM1 may be a biological indicator and potential target of bladder cancer. *Sci Rep* **14**, 23280 (2024).
170. Clarkson, E. & Lewis, A. BMP antagonist CHRDL2 enhances the cancer stem-cell phenotype and increases chemotherapy resistance in colorectal cancer. *Molecular oncology* (2025).
171. Xiong, H., *et al.* DARS expression in BCR/ABL1-negative myeloproliferative neoplasms and its association with the immune microenvironment. *Sci Rep* **14**, 16711 (2024).

172. Yeom, E., *et al.* Asparaginyl-tRNA Synthetase, a Novel Component of Hippo Signaling, Binds to Salvador and Enhances Yorkie-Mediated Tumorigenesis. *Frontiers in cell and developmental biology* **8**, 32 (2020).
173. Zitka, O., *et al.* Redox status expressed as GSH:GSSG ratio as a marker for oxidative stress in paediatric tumour patients. *Oncology letters* **4**, 1247-1253 (2012).
174. Brzozowa-Zasada, M., *et al.* Glutathione Reductase Expression and Its Prognostic Significance in Colon Cancer. *International journal of molecular sciences* **25**(2024).
175. Li, S., *et al.* LGALS4 inhibits glycolysis and promotes apoptosis of colorectal cancer cells via β-catenin signaling. *Oncology letters* **29**, 126 (2025).
176. Cao, Z.Q. & Guo, X.L. The role of galectin-4 in physiology and diseases. *Protein & cell* **7**, 314-324 (2016).
177. Song, Y., *et al.* HHIP Overexpression Suppresses Human Gastric Cancer Progression and Metastasis by Reducing Its CpG Island Methylation. *Frontiers in oncology* **10**, 1667 (2020).
178. Sun, H., *et al.* Hedgehog Interacting Protein 1 is a Prognostic Marker and Suppresses Cell Metastasis in Gastric Cancer. *Journal of Cancer* **9**, 4642-4649 (2018).
179. Bernard, J.K., McCann, S.P., Bhardwaj, V., Washington, M.K. & Frey, M.R. Neuregulin-4 is a survival factor for colon epithelial cells both in culture and in vivo. *J Biol Chem* **287**, 39850-39858 (2012).
180. Miano, C., *et al.* Neuregulin 4 Boosts the Efficacy of Anti-ERBB2 Neutralizing Antibodies. *Frontiers in oncology* **12**, 831105 (2022).
181. Wang, S., *et al.* NRG4 suppresses breast cancer metastasis via ERBB4-YAP1-mediated down-regulation of MMPs. *Genes & Diseases*, 101691 (2025).
182. Tu, Y., Tian, Y., Wu, Y. & Cui, S. Clinical significance of heat shock proteins in gastric cancer following hyperthermia stress: Indications for hyperthermic intraperitoneal chemoperfusion therapy. *Oncology letters* **15**, 9385-9391 (2018).
183. Thul, P.J. & Lindskog, C. The human protein atlas: A spatial map of the human proteome. **27**, 233-244 (2018).
184. Zhang, F., Tang, J.M., Wang, L., Wu, P.P. & Zhang, M. Immunohistochemical detection of RET proto-oncogene product in tumoral and nontumoral mucosae of gastric cancer. *Analytical and quantitative cytopathology and histopathology* **36**, 128-136 (2014).
185. Takahashi, M., Kawai, K. & Asai, N. Roles of the RET Proto-oncogene in Cancer and Development. *JMA journal* **3**, 175-181 (2020).
186. Jiao, X., *et al.* Overexpression of kallikrein gene 10 is a biomarker for predicting poor prognosis in gastric cancer. *World journal of gastroenterology* **19**, 9425-9431 (2013).
187. Shimura, T., *et al.* Urinary kallikrein 10 predicts the incurability of gastric cancer. *Oncotarget* **8**, 29247-29257 (2017).
188. Yu, B., *et al.* IL-6 facilitates cross-talk between epithelial cells and tumor- associated macrophages in Helicobacter pylori-linked gastric carcinogenesis. *Neoplasia (New York, N.Y.)* **50**, 100981 (2024).
189. Chalaris, A., Garbers, C., Rabe, B., Rose-John, S. & Scheller, J. The soluble Interleukin 6 receptor: generation and role in inflammation and cancer. *European journal of cell biology* **90**, 484-494 (2011).
190. Rašková, M., *et al.* The Role of IL-6 in Cancer Cell Invasiveness and Metastasis-Overview and Therapeutic Opportunities. *Cells* **11**(2022).
191. Zheng, X., Li, S. & Yang, H. Roles of Toll-Like Receptor 3 in Human Tumors. *Front Immunol* **12**, 667454 (2021).
192. Muresan, X.M., Bouchal, J., Culig, Z. & Souček, K. Toll-Like Receptor 3 in Solid Cancer and Therapy Resistance. *Cancers* **12**(2020).
193. Hsieh, M.L., Nishizaki, D., Adashek, J.J., Kato, S. & Kurzrock, R. Toll-like receptor 3: a double-edged sword. *Biomarker research* **13**, 32 (2025).
194. Galli, R., *et al.* Toll-like receptor 3 (TLR3) activation induces microRNA-dependent reexpression of functional RARβ and tumor regression. *Proc Natl Acad Sci U S A* **110**, 9812-9817 (2013).
195. Bie, Q., *et al.* Non-tumor tissue derived interleukin-17B activates IL-17RB/AKT/β-catenin pathway to enhance the stemness of gastric cancer. *Sci Rep* **6**, 25447 (2016).
196. Bie, Q., *et al.* IL-17B activated mesenchymal stem cells enhance proliferation and migration of gastric cancer cells. *Oncotarget* **8**, 18914-18923 (2017).

197. Bergström, S.H., Järemo, H., Nilsson, M., Adamo, H.H. & Bergh, A. Prostate tumors downregulate microseminoprotein-beta (MSMB) in the surrounding benign prostate epithelium and this response is associated with tumor aggressiveness. *Prostate* **78**, 257-265 (2018).
198. Dahlman, A.*, et al.* Evaluation of the prognostic significance of MSMB and CRISP3 in prostate cancer using automated image analysis. *Modern pathology : an official journal of the United States and Canadian Academy of Pathology, Inc* **24**, 708-719 (2011).
199. Olson, A.*, et al.* The comprehensive role of E-cadherin in maintaining prostatic epithelial integrity during oncogenic transformation and tumor progression. *PLoS Genet* **15**, e1008451 (2019).
200. Keil, K.P.*, et al.* DNA methylation of E-cadherin is a priming mechanism for prostate development. *Developmental biology* **387**, 142-153 (2014).
201. Qiu, L.X.*, et al.* The E-cadherin (CDH1)--160 C/A polymorphism and prostate cancer risk: a meta-analysis. *European journal of human genetics : EJHG* **17**, 244-249 (2009).
202. Liu, X.*, et al.* High expression of PDLIM5 facilitates cell tumorigenesis and migration by maintaining AMPK activation in prostate cancer. *Oncotarget* **8**, 98117-98134 (2017).
203. Huang, X., Qu, R., Ouyang, J., Zhong, S. & Dai, J. An Overview of the Cytoskeleton-Associated Role of PDLIM5. *Front Physiol* **11**, 975 (2020).
204. Shetty, A.*, et al.* Hepatoma-derived growth factor: A survival-related protein in prostate oncogenesis and a potential target for vitamin K2. *Urologic oncology* **34**, 483.e481-483.e488 (2016).
205. Ropiquet, F.*, et al.* FGF7/KGF triggers cell transformation and invasion on immortalised human prostatic epithelial PNT1A cells. *International journal of cancer* **82**, 237-243 (1999).
206. Story, M.T. Regulation of prostate growth by fibroblast growth factors. *World journal of urology* **13**, 297-305 (1995).
207. Leung, H.Y.*, et al.* Keratinocyte growth factor expression in hormone insensitive prostate cancer. *Oncogene* **15**, 1115-1120 (1997).
208. Jiang, G.*, et al.* PRSS3 promotes tumour growth and metastasis of human pancreatic cancer. *Gut* **59**, 1535-1544 (2010).
209. Hockla, A.*, et al.* PRSS3/mesotrypsin is a therapeutic target for metastatic prostate cancer. *Molecular cancer research : MCR* **10**, 1555-1566 (2012).
210. Ma, H.*, et al.* PRSS3/Mesotrypsin and kallikrein-related peptidase 5 are associated with poor prognosis and contribute to tumor cell invasion and growth in lung adenocarcinoma. *Sci Rep* **9**, 1844 (2019).
211. Zhou, T.*, et al.* Association of Glutathione S-transferase gene polymorphism with bladder Cancer susceptibility. *BMC cancer* **18**, 1088 (2018).
212. Schnakenberg, E., Breuer, R., Werdin, R., Dreikorn, K. & Schloot, W. Susceptibility genes: GSTM1 and GSTM3 as genetic risk factors in bladder cancer. *Cytogenetics and cell genetics* **91**, 234-238 (2000).
213. Chang, L.*, et al.* Restoration of LRIG1 suppresses bladder cancer cell growth by directly targeting EGFR activity. *Journal of experimental & clinical cancer research : CR* **32**, 101 (2013).
214. Wang, Y., Yi, K., Chen, B., Zhang, B. & Jidong, G. Elucidating the susceptibility to breast cancer: an in-depth proteomic and transcriptomic investigation into novel potential plasma protein biomarkers. *Frontiers in molecular biosciences* **10**, 1340917 (2023).
215. Mata-Rocha, M.*, et al.* Identification and Characterization of Novel Fusion Genes with Potential Clinical Applications in Mexican Children with Acute Lymphoblastic Leukemia. *International journal of molecular sciences* **20**(2019).
216. Li, L., Zhang, R., Liu, Y. & Zhang, G. ANXA4 Activates JAK-STAT3 Signaling by Interacting with ANXA1 in Basal-Like Breast Cancer. *DNA and cell biology* **39**, 1649-1656 (2020).
217. Yao, H., Sun, C., Hu, Z. & Wang, W. The role of annexin A4 in cancer. *Frontiers in bioscience (Landmark edition)* **21**, 949-957 (2016).
218. Yang, L.*, et al.* Downregulation of transgelin 2 promotes breast cancer metastasis by activating the reactive oxygen species/nuclear factor-κB signaling pathway. *Mol Med Rep* **20**, 4045-4258 (2019).
219. Kim, R.H.*, et al.* DJ-1, a novel regulator of the tumor suppressor PTEN. *Cancer cell* **7**, 263-273 (2005).
220. Jin, W. Novel Insights into PARK7 (DJ-1), a Potential Anti-Cancer Therapeutic Target, and Implications for Cancer Progression. *Journal of clinical medicine* **9**(2020).

221. Wang, T., *et al.* PARK7 is a Key Regulator of Oxidative Stress - Related Breast Cancer Risk: A Multi-Omics Study. *Journal of Cancer* **16**, 2877-2889 (2025).
222. Lin, M., Li, Y., Qin, S., Jiao, Y. & Hua, F. Ubiquitin-like modifier-activating enzyme 7 as a marker for the diagnosis and prognosis of breast cancer. *Oncology letters* **19**, 2773-2784 (2020).
223. Alatawi, S., *et al.* Identification of UBA7 expression downregulation in myelodysplastic neoplasm with SF3B1 mutations. *Sci Rep* **15**, 10856 (2025).
224. Vazquez-Ortiz, G., *et al.* Overexpression of cathepsin F, matrix metalloproteinases 11 and 12 in cervical cancer. *BMC cancer* **5**, 68 (2005).
225. Ji, C., *et al.* Cathepsin F Knockdown Induces Proliferation and Inhibits Apoptosis in Gastric Cancer Cells. *Oncology research* **26**, 83-93 (2018).
226. Wei, S., *et al.* Cathepsin F and Fibulin-1 as novel diagnostic biomarkers for brain metastasis of non-small cell lung cancer. *British journal of cancer* **126**, 1795-1805 (2022).
227. Huang, K., Wu, Y., Xie, Y., Huang, L. & Liu, H. Analyzing mRNAsi-Related Genes Identifies Novel Prognostic Markers and Potential Drug Combination for Patients with Basal Breast Cancer. *Disease markers* **2021**, 4731349 (2021).
228. Taylor, N.J., *et al.* Inherited variation at MC1R and ASIP and association with melanoma-specific survival. *International journal of cancer* **136**, 2659-2667 (2015).
229. Lin, W., *et al.* ASIP genetic variants and the number of non-melanoma skin cancers. *Cancer causes & control : CCC* **22**, 495-501 (2011).
230. Gudbjartsson, D.F., *et al.* ASIP and TYR pigmentation variants associate with cutaneous melanoma and basal cell carcinoma. *Nat Genet* **40**, 886-891 (2008).
231. Suzuki, I., *et al.* Agouti signaling protein inhibits melanogenesis and the response of human melanocytes to alpha-melanotropin. *The Journal of investigative dermatology* **108**, 838-842 (1997).
232. Li, Y., Li, Q., Cao, Z. & Wu, J. Multicenter proteome-wide Mendelian randomization study identifies causal plasma proteins in melanoma and non-melanoma skin cancers. *Commun Biol* **7**, 857 (2024).
233. Tiwary, S., *et al.* ERBB3 is required for metastasis formation of melanoma cells. *Oncogenesis* **3**, e110 (2014).
234. Buac, K., *et al.* NRG1 / ERBB3 signaling in melanocyte development and melanoma: inhibition of differentiation and promotion of proliferation. *Pigment cell & melanoma research* **22**, 773-784 (2009).
235. Grzywa, T.M., *et al.* Myeloid Cell-Derived Arginase in Cancer Immune Response. *Front Immunol* **11**, 938 (2020).
236. Arlauckas, S.P., *et al.* Arg1 expression defines immunosuppressive subsets of tumor-associated macrophages. *Theranostics* **8**, 5842-5854 (2018).
237. Canè, S., *et al.* Neutralization of NET-associated human ARG1 enhances cancer immunotherapy. *Sci Transl Med* **15**, eabq6221 (2023).
238. Ramji, K., *et al.* Targeting arginase-1 exerts antitumor effects in multiple myeloma and mitigates bortezomib-induced cardiotoxicity. *Sci Rep* **12**, 19660 (2022).
239. Bruno, A., *et al.* Human RNASET2: A Highly Pleiotropic and Evolutionary Conserved Tumor Suppressor Gene Involved in the Control of Ovarian Cancer Pathogenesis. *International journal of molecular sciences* **23**(2022).
240. Wu, L., Xu, Y., Zhao, H. & Li, Y. RNase T2 in Inflammation and Cancer: Immunological and Biological Views. *Front Immunol* **11**, 1554 (2020).
241. Acquati, F., *et al.* Innate Immune Response Regulation by the Human RNASET2 Tumor Suppressor Gene. *Front Immunol* **10**, 2587 (2019).
242. Tian, M., *et al.* IRF3 prevents colorectal tumorigenesis via inhibiting the nuclear translocation of β-catenin. *Nat Commun* **11**, 5762 (2020).
243. Guinn, Z., Brown, D.M. & Petro, T.M. Activation of IRF3 contributes to IFN-γ and ISG54 expression during the immune responses to B16F10 tumor growth. *International immunopharmacology* **50**, 121-129 (2017).
244. Szklarczyk, D., *et al.* The STRING database in 2021: customizable protein-protein networks, and functional characterization of user-uploaded gene/measurement sets. *Nucleic Acids Res* **49**, D605-d612 (2021).

245. Nonnast, E., Mira, E. & Mañes, S. The role of laminins in cancer pathobiology: a comprehensive review. *Journal of translational medicine* **23**, 83 (2025).
246. Sheng, Z., Beck, P., Gabby, M., Habte-Mariam, S. & Mitkos, K. Molecular Basis of Oncogenic PI3K Proteins. *Cancers* **17**(2024).
247. Jiang, B.H. & Liu, L.Z. PI3K/PTEN signaling in angiogenesis and tumorigenesis. *Advances in cancer research* **102**, 19-65 (2009).
248. Yi, M., *et al.* Targeting cytokine and chemokine signaling pathways for cancer therapy. *Signal transduction and targeted therapy* **9**, 176 (2024).
249. Chen, Z., *et al.* Ligand-receptor interaction atlas within and between tumor cells and T cells in lung adenocarcinoma. *International journal of biological sciences* **16**, 2205-2219 (2020).
250. Bansal, A. & Simon, M.C. Glutathione metabolism in cancer progression and treatment resistance. *The Journal of cell biology* **217**, 2291-2298 (2018).
251. Heerma van Voss, M.R., van Diest, P.J. & Raman, V. Targeting RNA helicases in cancer: The translation trap. *Biochimica et biophysica acta. Reviews on cancer* **1868**, 510-520 (2017).
252. Winkler, J., Abisoye-Ogunniyan, A., Metcalf, K.J. & Werb, Z. Concepts of extracellular matrix remodelling in tumour progression and metastasis. *Nat Commun* **11**, 5120 (2020).
253. Kennedy, L., Sandhu, J.K., Harper, M.E. & Cuperlovic-Culf, M. Role of Glutathione in Cancer: From Mechanisms to Therapies. *Biomolecules* **10**(2020).
